# A Phase IV Trial of Proton Therapy in Children: The First Report from the SJPROTON1 Study

**DOI:** 10.1101/2022.11.28.22282836

**Authors:** John T. Lucas, Matthew Marker, Jared Becksfort, Tushar Patni, Austin M. Faught, Christopher L. Tinkle, Valerie J. Groben, Elizabeth Burghen, Haley Ruleman, Chia-Ho Hua, Sue C. Kaste, Sahaja Acharya, Noah D. Sabin, Melissa Hudson, Shengjie Wu, Yimei Li, Matthew J. Krasin, Thomas E. Merchant

## Abstract

**Importance:** Early reports suggest adverse events following proton therapy (PT) for childhood cancer are more prevalent given the increased number of PT centers and use in clinical trials.

**Objective:** To assess treatment-failure and toxicity events following Pencil Beam Scanning (PBS)-PT in children.

**Design, Setting, and Participants:** The single-institution Phase IV surveillance trial screened 856 children, of which 528 were eligible for PBS-PT, and 500 enrolled in SJPROTON1 from 2017–2020. The median follow-up was 2.1 years (range 1.1-4.4). PBS-RT attributable toxicities were systematically identified, and graded at baseline to four years following PBS-PT.

**Interventions:** All patients underwent PBS-PT or combined photon/PBS-PT.

**Main Outcomes and Measures:** The primary objective was the CI of ≥grade 3 non-hematologic, PBS-PT attributable Common Terminology Criteria for Adverse Events (CTCAE) v 4.0. Additional outcomes included PBS-PT attributable hospitalization, toxicity related procedures, and treatment-related mortality. Pre-specified toxicities including necrosis, vasculopathy, neurologic deficits, and fracture/osteoradionecrosis were further characterized (any grade) as a secondary objective. Medical record review augmented in-person visits for failure and adverse event reporting. Competing risk regression was used to evaluate predictors of ≥grade 3 PBS-PT attributable toxicity.

**Results:** At two years, the event-free survival was 73.2% (95% CI, 68.9%-77.8%). Distant and local failure predominated with a 2-year cumulative incidence (CI) of 16.5% (95% CI 13.1-20.4) and 6.8% (95% CI 4.6-9.5) respectively. The CI of ≥ grade 3 toxicity events at four years was 24.5%; including necrosis (3.7%), permanent neurologic deficit (2.9%), and fracture/osteoradionecrosis (0.79%). The rates of hospitalization and procedures due to PBS-PT– attributable toxicity was 3.9% and 7.6%, respectively. Predictors of an increased event-specific hazard for any ≥ grade 3 toxicity included baseline total toxicity burden (TTB) (HR 1.043, 95% CI 1.012-1.07, p=0.007), use of mixed photon/PBS-PT (HR 2.62, 95% CI 1.51-4.54, p=0.006) in the CNS population, and treatment of the pelvic body site (HR 4.25, 95% CI 1.08-16.72, p=0.038), and TTB (HR 1.1, 95% CI 1.04-1.16, p=0.002) in the non-CNS population, respectively.

**Conclusions and Relevance:** Children treated with PBS-PT experience a low incidence of toxicity, but subsets may be at increased risk, and experience toxicity that requires additional procedures or hospitalization.

**Key Points:** *Question:* What is the efficacy and safety profile of Pencil Beam Scanning Proton therapy (PBS-PT) in children?

*Findings:* In this single institution phase IV trial, the rate of local failure and clinically significant ≥grade 3 PBS-RT attributable toxicity were low, but a subset of patients may be at increased risk for hospitalization and procedures related to managing PBS-RT attributable adverse events.

*Meaning:* PBS-RT has an acceptable therapeutic ratio in children but the risk of adverse events requiring hospitalization or subsequent procedures may be increased in select populations.

## INTRODUCTION

The promise of proton therapy (PT) to cause less adverse effects compared to conventional photon therapy has led to its rapid adoption by the pediatric oncology community and preferential referral for treatment.^1^ Disproportionate use of PT and differential access afforded by higher socioeconomic status may have confounded early estimates of its benefit^2^.

Although years of systematic follow-up are required to evaluate children for long-term effects of radiation, early evidence suggests that PT limits late effects across multiple outcomes (e.g., neurologic, endocrine, cognitive)^3-10^, with ongoing efforts to demonstrate similar advantages in quality of life and reduced induction of secondary tumors attributable to radiotherapy.^11-16^

Prospective, in-person, direct assessment using objective toxicity scales is the ideal approach to accurate evaluation of adverse events.^17^ To address the incidence of subsequent cancers, general disease, and/or site-specific outcomes, many previous investigations have identified such information from registries^2^; however, that approach has numerous limitations.^18^. A Phase IV clinical trial may circumvent those limitations by employing a more systematic approach to characterizing toxicity directly in a manner that is not specific to disease, sampling, or systemic therapy.

In 2015, St. Jude Children’s Research Hospital opened the first PT center dedicated solely to the treatment of children and initiated a Phase IV clinical trial SJPROTON1 to estimate treatment-related complications in children receiving PBS-PT.^19^ Here we report our preliminary findings from the first 500 children with cancer treated on this single-institution study.

## METHODS

### Study Design and Population

The SJPROTON1 trial^20^was designed to assess the effects of PBS-PT on pediatric patients with cancer treated at St. Jude. This Phase IV trial is an observational study; all PBS-PT administered was prescribed by disease-specific therapeutic protocols or non-protocol treatment plans. Patients enrolled in SJPROTON1 were assessed at baseline (before any proton exposure) to document baseline patient and disease characteristics. After completion of PBS-PT, the participants undergo standard follow-up evaluations at 4–6 weeks or 6 months and at 1, 2, 3, 5, and 10 years to document PBS-PT attributable acute and late complications, disease control, mortality, and survival (SF1). Quality assurance reviews of the completeness of data are done at 6 months, and years 1, 5, and 10 and are documented in SF2. The study’s primary objective is to estimate the CI of Common Terminology for Adverse Events (CTCAE) v4.0 and SJLIFE-modified CTCAE^21^ ≥ grade 3 toxicity events following PBS-PT. The secondary objectives included estimating the CI of specific toxicities including CNS necrosis, vasculopathy, neurologic deficits, osteoradionecrosis, and fracture. Additional events of interest included estimating the rate of PBS-PT attributable hospitalization, and utilization of procedures to mitigate PBS-PT attributable toxicity.

The SJPROTON1 trial was reviewed and approved by the Institutional Review Board at St. Jude. Informed consent was obtained from the parent, caregiver, or patient, with assent from the patient, as appropriate.

### Toxicity Assessment, Outcomes, and Analysis Methods

All patient information was gained via clinical evaluation or review of electronic medical records. Traits of interest, treatment regimens, disease categories, toxicity categories and grading, patient outcomes, record search, and statistical analyses are examined further in the supplementary methods.

### Statistical Analyses

The MOSAIQ SQL database was queried for the trial period (2017–2020). Categorical variables were summarized as count and frequencies, and continuous variables were summarized using the median, interquartile range (IQR), and range. CI of ≥grade 3 toxicities, hospitalization, and procedure utilization were estimated by accounting disease progression and death as competing events. When calculating the CI of treatment failures, death was used as the competing event. The Fine and Gray method (1999) was used to investigate the association between covariates and the CI of events of interest. Cox proportional hazard models were used to explore the association of risk factors on time to symptom resolution and time to radiographic resolution.

The log-rank test was also used to compare the radiographic resolution between groups with and without intervention. In the case of competing risk regression, models were done for each individual risk factor first. Factors identified with p-value <0.05 and other possible confounders were included in multivariable regression models when the sample size and event rate were sufficient. All the analyses were done in R 3.6.2.

## RESULTS

### Patient Demographics

Between 2017 and 2020 we enrolled 500 children with cancer who were undergoing PBS-PT to their primary and metastatic sites onto the Phase IV SJPROTON1 trial (Fig. 1). The median age was 10.4 years (interquartile range [IQR] 5.65–15.3 years). The cohort included 323 (64.6%) participants treated on a prospective therapeutic trial, and 177 (35.4%) patients treated on non-protocol treatment plans. Participants underwent either mixed photon/PBS-PT (n=91, 18.2%) or PBS-PT (n = 409, 81.8%). The most commonly treated sites included the CNS (n=349, 69.8 %), head and neck (n=51, 10.2%), abdomen (n=28, 5.6%), pelvis (n=28, 5.6%), musculoskeletal (n=22, 4.4%), and chest (n=22, 4.4%) (Table 1). The majority (90.6%) of participants had not received prior radiotherapy, but 30 (6%) received prior photon therapy, and 17 (3.4%) received prior PT. Extended information on demographic, economic, and disease related parameters for CNS and non-CNS patients are shown in ST1, ST2. At the time of this writing, 353 participants were in active protocol-defined follow-up, 10 were removed from follow-up due to the absence of the need for further PBS-PT related toxicity surveillance, 127 experienced disease progression and follow-up was terminated, five withdrew from the study, and 75 died of various causes (Fig. 1).

**Figure 1.**
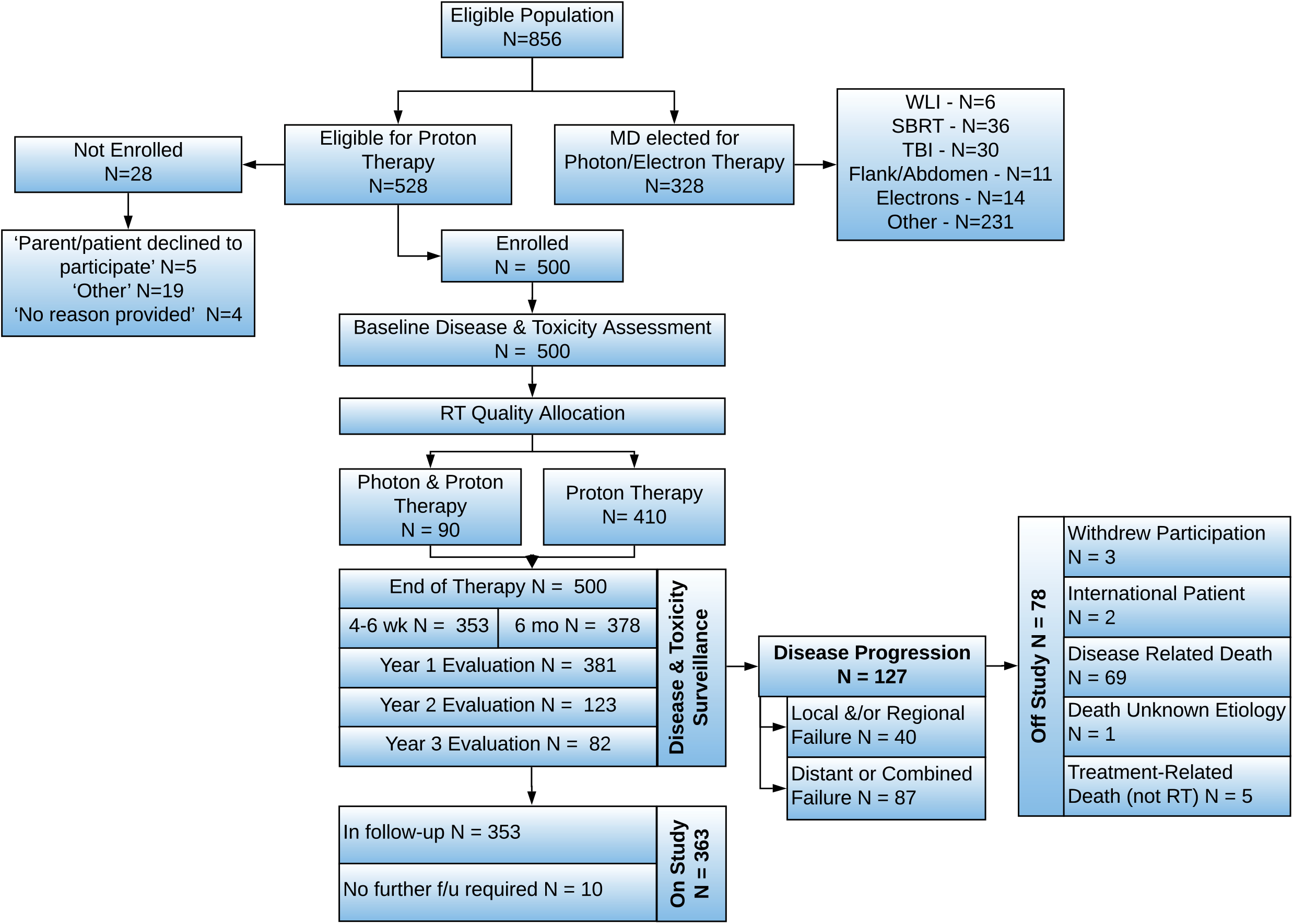
SJPROTON1 Consort Diagram. Abbreviations: f/u, follow-up; N, number; RT, radiotherapy

**Table 1.** Status and Characteristics of SJPROTON1 Participants Grouped by Treated Body Site.

|  | Abdomen<br>(N=28) | Chest<br>(N=21) | CNS<br>(N=349) | H&N<br>(N=51) | MSK<br>(N=22) | Pelvis<br>(N=28) | Overall<br>(N=500) |
| --- | --- | --- | --- | --- | --- | --- | --- |
| **Trial Status** |  |  |  |  |  |  |  |
| Off Trial,<br>Records Retained | 8 (28.6%) | 3 (14.3%) | 45 (12.9%) | 9(17.6%) | 7 (31.8%) | 6(21.4%) | 78 (15.6%) |
| In follow-up | 16 (57.1%) | 15 (71.4%) | 272 (77.9%) | 36 (70.6%) | 11 (50.0%) | 15 (53.6%) | 366 (73.2%) |
| Cancer Progression | 4 (14.3%) | 3 (14.3%) | 32 (9.2%) | 6(11.8%) | 4(18.2%) | 7 (25.0%) | 56 (11.2%) |
| **Prior RT** |  |  |  |  |  |  |  |
| Prior Photon | 3 (10.7%) | 1 (4.8%) | 15(4.3%) | 7 (13.7%) | 2 (9.1%) | 2 (7.1%) | 30 (6.0%) |
| Prior Proton | 0 (0%) | 0 (0%) | 17(4.9%) | 0 (0%) | 0 (0%) | 0 (0%) | 17 (3.4%) |
| No Prior RT | 25 (89.3%) | 20 (95.2%) | 317 (90.8%) | 44 (86.3%) | 20 (90.9%) | 26 (92.9%) | 453 (90.6%) |
| **Treatment Modality** |  |  |  |  |  |  |  |
| Pho/Pro | 6 (21.4%) | 1 (4.8%) | 73 (20.9%) | 6(11.8%) | 3 (13.6%) | 1 (3.6%) | 90 (18.0%) |
| Proton | 21 (75.0%) | 20 (95.2%) | 276 (79.1%) | 44 (86.3%) | 19(86.4%) | 27 (96.4%) | 410 (82%) |
| **Enrollment Category** |  |  |  |  |  |  |  |
| Retrospective | 13 (46.4%) | 7 (33.3%) | 149 (42.7%) | 20 (39.2%) | 11 (50.0%) | 11 (39.3%) | 212 (42.4%) |
| Prospective | 15 (53.6%) | 14 (66.7%) | 200 (57.3%) | 31 (60.8%) | 11 (50.0%) | 17 (60.7%) | 288 (57.6%) |
| N = Number, CNS = Central Nervous System, H&N = Head and Neck, MSK = Musculoskeletal |  |  |  |  |  |  |  |

### Baseline and Radiation Attributable Toxicity Burden

Prior to the initiation of PBS-PT, we detected 246 adverse events attributable to PBS-PT in 159 participants, with a higher baseline TTB in identified in participants with sellar, embryonal, soft-tissue sarcoma, ependymal, or germ cell tumors (SF6, SF7). We also identified 171 ≥ grade 3 toxicities among 101 participants. The CI of toxicity events at Years 1–4 after completion of PBS-PT was 15.5% (95% CI 12.5-18.9), 19.5% (95% CI 16-23.3), 23.5% (95% CI 19.1-28.2), and 25.5% (95% CI 20.5-30.9), respectively (Fig. 2). The CIs at 2 years by treated site were as follows: CNS 20.7% (95% CI 16.4-25.4), head/neck 17.3% (95% CI 7.7-30.1), chest 9.8% (95% CI 1.6-27.3), abdomen 7.1% (95% CI 1.2-20.7), pelvis 25.2% (95% CI 10.9-42.4), and musculoskeletal 22.7% (95% CI 8-41.9). The most common toxicities included hearing loss, other neurologic deficits, CNS necrosis, radiation dermatitis, and endocrinopathy.

**Figure 2.**
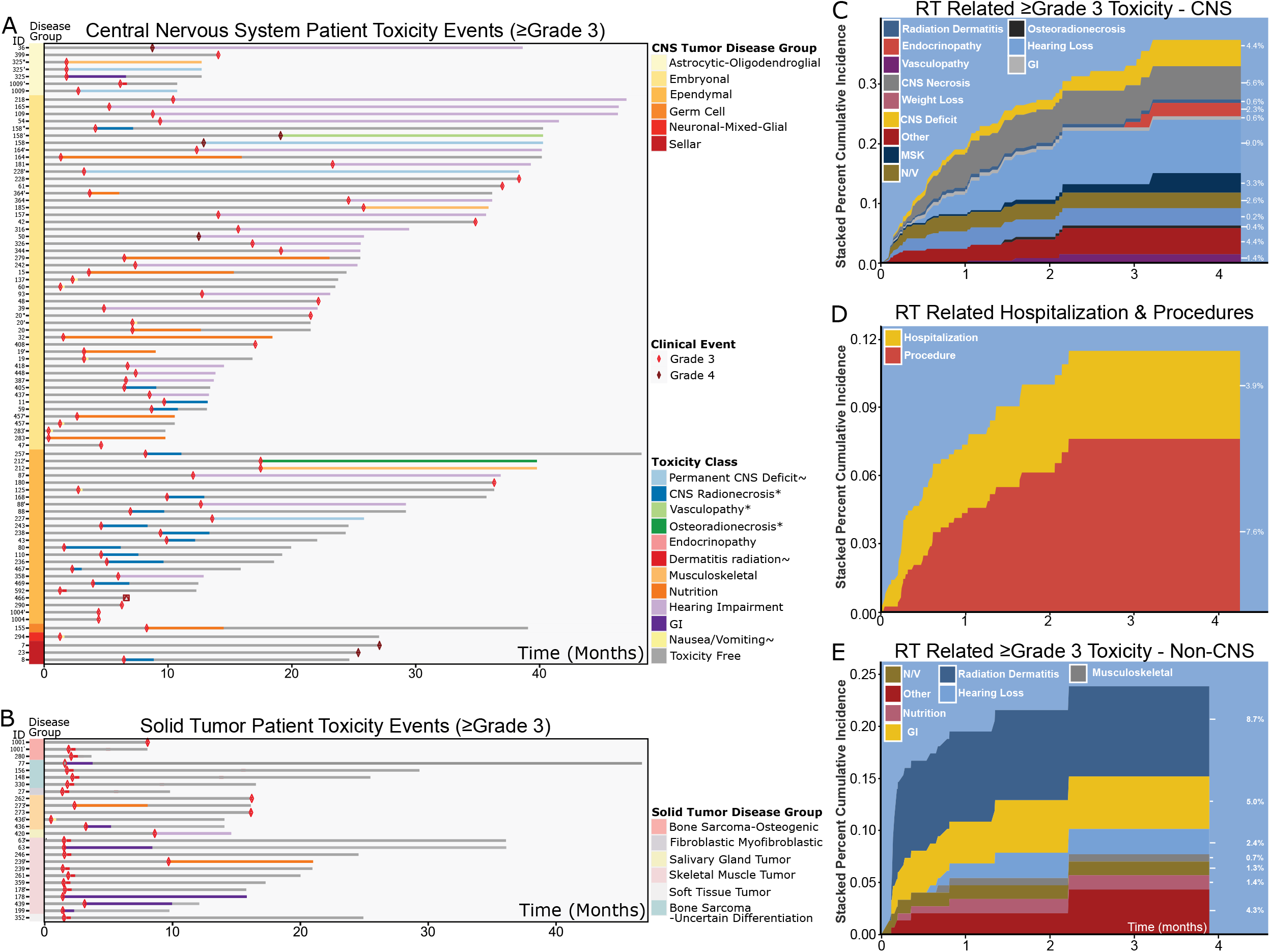
PBS-PT–Attributable Toxicity Events. All PBS-PT–attributable toxicity events in patients with CNS tumors (A) or solid tumors (B). Grey lines indicate the absence or resolution of a toxicity depending on their relationship to the red or dark red diamond representing the adverse event. (C) Stacked CI curves of ≥ grade 3 toxicity in patients with a CNS tumor. (D) Stacked CI curves of PBS-PT–attributable hospitalizations and procedures. (E) Stacked CI of ≥ grade 3 toxicity events in patients with solid tumors. Each CI plot is shown as a multistate curve, and the sum of all toxicities is indicated by the stacked width of the sum of each solid color at each timepoint. The width of each curve reflects the CI of that toxicity. The duration of the ≥ grade 3 event in the swimmer plots is indicated by the length of the colored bar. When multiple ≥ grade 3 toxicity events occurred after PBS-PT, an apostrophe indicates a repeated patient identifier (ID).

### Toxicities Associated with Treating CNS Tumors

Among the 349 participants with CNS tumors, we identified 125 PBS-PT–attributable toxicities in 76 participants (Fig. 2A). The CIs of ≥ grade 3 toxicities in these participants at Years 1–4 after completion of therapy was 15.5% (95% CI 11.9-19.6), 20.7% (95% CI 16.4-25.4), 25.2% (95% CI 19.8-30.9), and 27.6% (95% CI 21.5-34), respectively (Fig. 2C). The most common toxicities in participants with CNS tumors were hearing loss, neurologic deficits, and CNS necrosis (Fig. 2C). These events occurred more frequently in participants with embryonal, ependymal, or astrocytic-oligodendroglial tumors (SF8). The most common toxicities and their maximum grades are shown in Fig. 3 as event counts by disease group and event type.

**Figure 3.**
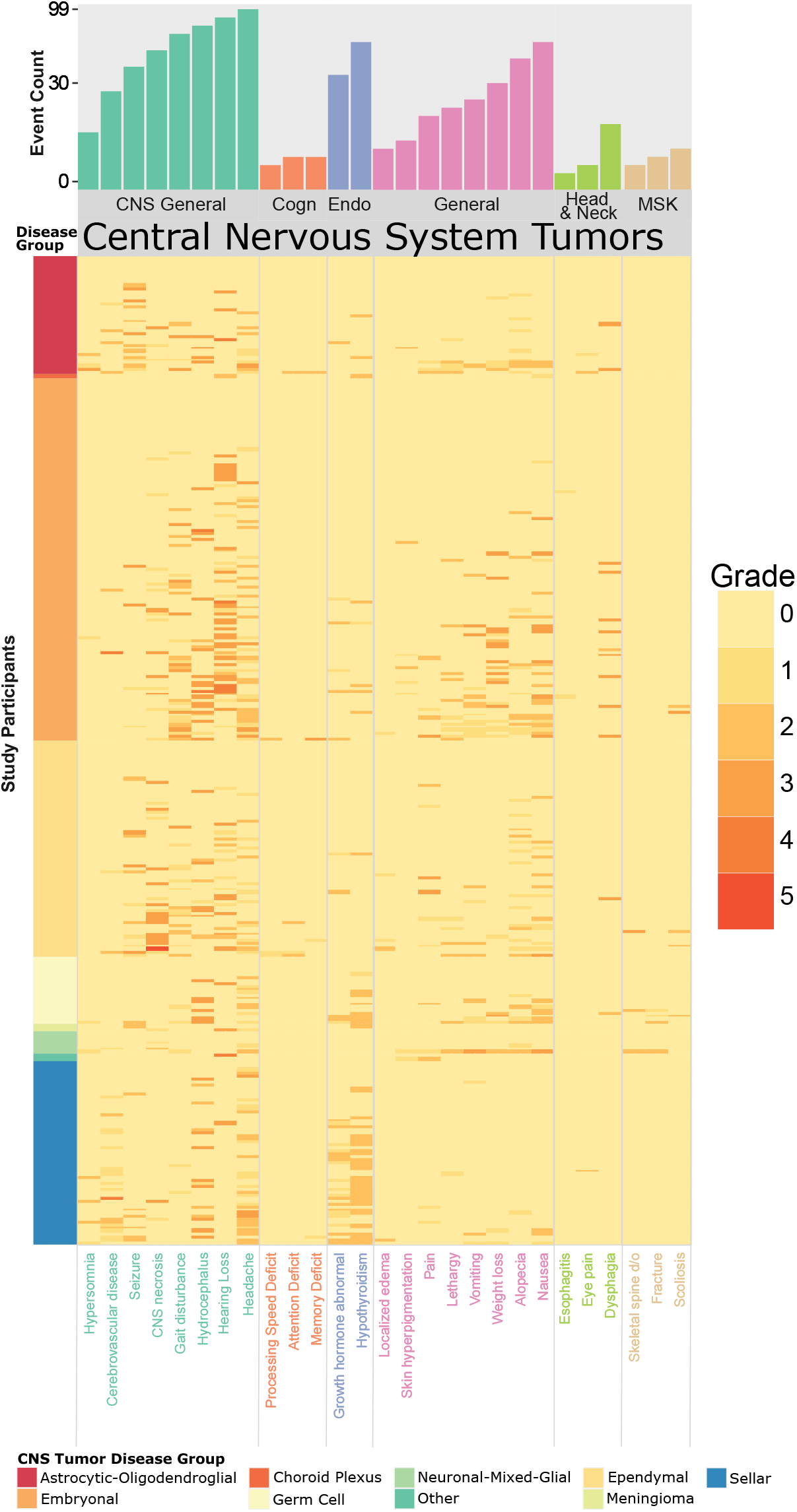
Frequency Heatmap Demonstrating Frequency of All-Grade Toxicity Events Across CNS Tumors by Disease Group and Toxicity Class. Each row in the heatmap represents a single participant’s cumulative frequency of toxicity events and grade across all trial timepoints. The most common toxicity classes are represented in column headers, and the specific toxicity types within that class are shown along the x-axis. Severity of the grades observed for each patient are shown as a colorimetric heatmap in which higher grade toxicities are presented as progressively darker orange. The raw counts and frequencies represented in the heatmap is in Supplementary Data (SD5). The corresponding figures for solid tumor and leukemia/lymphoma cases are shown in Supplementary Figures S10 and S11.

### Toxicities Associated with Treating Solid Tumors or Lymphoma

Among participants with solid tumors, we detected 45 PBS-PT–attributable toxicities in 26 participants (Fig. 2B). The CIs of ≥ grade 3 toxicity events at Years 1–4 after completion of therapy was 15.4% (95% CI 10.2-21.7), 16.5% (95% CI 10.9-23), 18.8% (95% CI 12-26.9) (Fig. 2E). The most common observed toxicities in these participants were radiation dermatitis, gastrointestinal toxicity, and hearing loss, which occurred more frequently in bone sarcomas, sarcoma not otherwise classified, and fibroblastic-myofibroblastic tumors (SF9). Toxicities and their maximum grades are shown in SF10 and SF11.

### Evaluation and Resolution of Prespecified Select Toxicities

We conducted an additional review to better understand the potential for multimodality therapy and preexisting injury to contribute to the PBS-PT–attributable events specified in the study’s secondary objectives.

#### CNS Necrosis

All-grade CNS necrosis was more common in astrocytic-oligodendroglial, embryonal, and ependymal tumors, though the majority (68%) of ≥ grade 3 events occurred in participants with ependymal disease (SF12A). PBS-PT attribution in all but 2 events was likely or plausible. Although the location of CNS necrosis could not be solely attributed to end-of-range effects, 15 of 45 (33.3%) cases occurred in or near the cervicomedullary junction and/or lower medulla. This finding suggested a potential influence of beam arrangement when treating fourth ventricle tumors.

Resolution occurred in 9 of 14 (64.3%) symptomatic toxicity events and in 33 of 35 (91.4%) radiographic events (SF12 C, H, K, SD7). Grade 3 or more severe events were treated with a mix of bevacizumab, dexamethasone, and hyperbaric oxygen therapy, all of which aided symptomatic resolution with comparable efficacy, though bevacizumab increased the hazard for radiographic resolution (HR 4.3, 95% CI 1.6-11) (SF12 I,J). The only factors associated with symptom resolution were eloquence of the region of necrosis (Sawya Grade HR 0.51, 95% CI 0.15-1.78, p=0.29) and chemotherapy exposure (HR 0.14, 95% CI 0.025-0.74, p=0.021) (SF12 L,M).

#### Vasculopathy

Of the 41 PBS-PT-related vasculopathy events identified in 32 participants, all but 2 were low grade (SF13 A1, A2, SD6); 26 of 41 (63.4%) vasculopathy events were asymptomatic stenoses detected by MRA in participants with sellar tumors (SD6). More than half of the events were documented within 16 months of PBS-PT (SF13 B). Medical interventions were recommended for 13 participants, and indirect revascularization was recommended for 2 (SF13 C2, C3). Vasculopathy events which were more likely PBS-PT– attributable were associated with longer times to the event (SF13 D).

#### Hearing and Vision Loss

Sensorineural hearing loss was the most common anatomic neurologic deficit we detected after PBS-PT (N=25 patients, N=37 grade 3 events[ears]) (SF14 A,B, SD8). Ninety percent of grade 3 hearing events occurred in those with embryonal tumors who received post-irradiation chemotherapy and in those who received a combination of photon/PBS-PT (SF14D). The rate of hearing loss attribution being likely attributed to PBS-PT were reduced in the PBS-PT only cohort relative to mixed photon/PBS-PT. The likelihood of attribution was heavily influenced by the mean radiation dose to the cochlea and chemotherapy exposure (i.e., cisplatin). Per self-reports, 100% of participants had their hearing adequately restored by hearing aids or other assistive hearing devices (SF14D).

#### Endocrinopathy

All 28 endocrinopathy events detected were low grade (grades 1-2) and more common in participants with CNS tumors in the sellar region or embryonal tumors that required high dose craniospinal irradiation (CSI) (SD8). Hormone replacement was initiated in all but 1 participant, at the parent’s request following documentation of poor linear growth or declining growth velocity.

#### Neurologic Deficits

Although the 12 non-anatomic CNS deficits identified as potential PBS-PT–attributable varied, they were typically low grade and indirectly related to either the tumor (i.e., 2 participants experienced pseudo-progression) or another toxicity event (i.e., 4 participants also had CNS necrosis) (SF3 A2, B2, SD9). Only 3 (23.1%) cases were classified as likely attributable to PBS-PT (SF3 A3). In 8 (66.7%) participants, the deficits improved with supportive interventions (SF3 C2).

#### Musculoskeletal Events

Seventeen participants experienced either bone injury or changes in bone growth: 8 participants suffered a fracture, 9 experienced osteoradionecrosis, and 1 had tooth development hypoplasia (SF15, SD10). Many musculoskeletal toxicity events were associated with prior steroid use or bone injury (SF15 A1, A2, B1, B6). The most common type of fracture was vertebral compression fracture, which occurred at a median of 9 months from PBS-PT and often required limited intervention. Radiation dose had a strongly relation with osteoradionecrosis (SF15 B4). Musculoskeletal toxicity events that occurred soon after PBS-PT were typically in CSI cases to modest doses (15-36 GyRBE) with prior prolonged steroid exposure (SF15 A2, A6, D).

#### Nutritional and Gastrointestinal Events

Nutritional and gastrointestinal toxicities were common among participants with CNS or solid tumors. Most events occurred during or immediately after PBS-PT, were frequently attributable to combined modality therapy, and resolved with conservative interventions (SF16 A, C, D, SD11). Only 4 participants received PBS-PT doses potentially high enough to result in substantial mucosal doses due to the organ’s approximation to the target: 2 participants experienced esophagitis, and 2 had proctitis (SF16 A, B).

### Management of PBS-PT–Attributable Toxicity Events

At 2 years after completion of therapy, the CI of hospitalization and procedure utilization due to a PBS-PT–attributable toxicity were 3.9%(95% CI 2.4-6) and 7.6%(95% CI 4.1-8.7), respectively (Fig. 2D). The most common reason for hospitalization was nausea/vomiting (nutrition/fluid support). Other toxicity events resulting in hospitalization are detailed in the Supplementary Data (SD3). The most common procedures were hyperbaric oxygen therapy, bevacizumab, and gastrostomy tube placement or total parenteral nutrition initiation.

### Treatment Failure

Treatment failure occurred in 127 (25.4%) participants, with a 2-year CI of 26.6%(95% CI 22.3-31.1) (Fig. 4). Distant failure predominated with a 2-year CI of 16.5%(95% CI 13.1-20.4); that of local failure was 6.8%(95% CI 4.6-9.5) (Fig. 4B).

**Figure 4.**
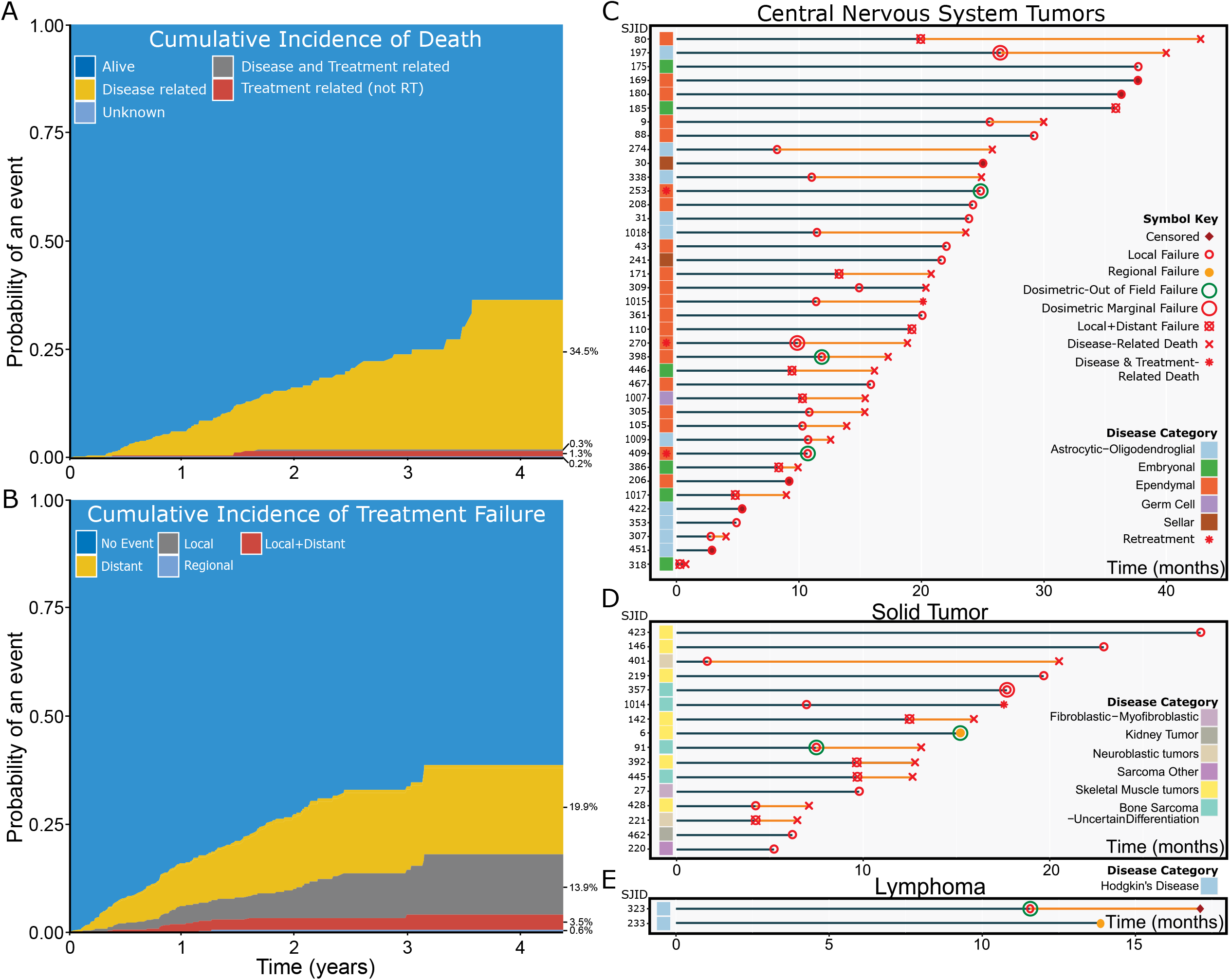
Cancer-Related Events in Patients Treated with Proton Therapy. (A) CI of death. (B) CI of treatment failure events. (C-E) Swimmer plots showing local treatment failure events in individual patients with CNS tumors (C), solid tumors (D), or lymphoma (E). The gray line indicates disease-free survival. Local, regional, and combined treatment failure events are coded as circles, and death is indicated as an “x.” The dosimetric type of failure for each local treatment failure event is indicated by a surrounding circle, where marginal failures (i.e., disease recurred outside the 80% isodose line) are shown as red circles. Out-of-field failures (i.e., disease recurred outside the 20% isodose line) are shown as green circles.

We prespecified the rate of local, marginal failures as a secondary objective due to heightened concerns about the improved conformality of PBS-PT and the dosimetric uncertainties related to treatment delivery and positional and anatomic changes during treatment. Of the 51 local and/or distant failure events, only 3 (5.8%) were marginal dosimetric failures. These included a base of skull Ewing sarcoma (ID-357), supratentorial high-grade glioma (ID-197), and relapsed fourth ventricle ependymoma (ID-270) (Fig. 4C, D). Although we cannot directly compare treatment failures in each disease and histology, the low rate of local and marginal failures across the cohort suggests that the dosimetric uncertainty in proton delivery made a limited contribution to PBS-PT–attributable failures.

### Risk factors Associated with PBS-PT–Attributable ≥ Grade 3 Toxicity Events

Univariate and multivariate competing risk regression models for the CNS and non-CNS patients were considered separately and reviewed to determine event-specific hazard for any grade ≥3 toxicity event (Table 2, ST3). After correcting for the influence of age, extent of disease, and prior radiotherapy use, only mixed photon/PBS-PT (HR 2.62, 95% CI 1.50-4.54, p=0.0006) predicted for an increased hazard for any grade **≥**3 toxicity event. Multivariate analysis for the non-CNS patient population identified pelvic body site (HR 4.25, 95% CI 1.08-16.72, p=0.038) and baseline TTB (HR 1.1, 95% CI 1.04-1.16, p=0.0015) as the only significant predictors of an increased hazard for any grade **≥**3 toxicity event.

**Table 2.**
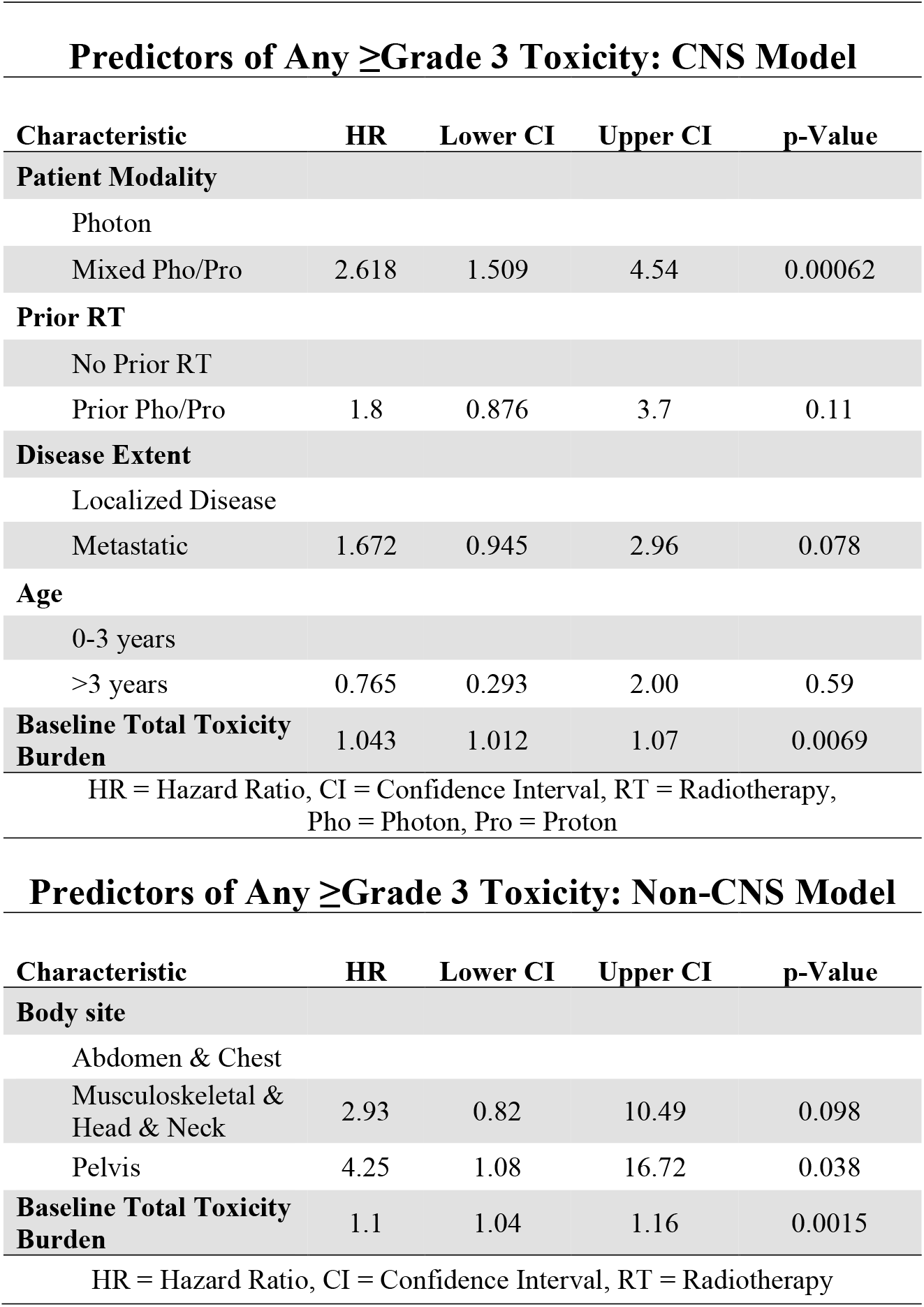
Body Site Stratified Predictors of Any ≥ Grade 3 Proton Attributable Toxicity.

| **Predictors of Any ≥Grade 3 Toxicity: CNS Model** |  |  |  |  |
| --- | --- | --- | --- | --- |
| **Characteristic** | **HR** | **Lower CI** | **Upper CI** | **p-Value** |
| **Patient Modality** |  |  |  |  |
| Photon |  |  |  |  |
| Mixed Pho/Pro | 2.618 | 1.509 | 4.54 | 0.00062 |
| **Prior RT** |  |  |  |  |
| No Prior RT |  |  |  |  |
| Prior Pho/Pro | 1.8 | 0.876 | 3.7 | 0.11 |
| **Disease Extent** |  |  |  |  |
| Localized Disease |  |  |  |  |
| Metastatic | 1.672 | 0.945 | 2.96 | 0.078 |
| **Age** |  |  |  |  |
| 0-3 years |  |  |  |  |
| >3 years | 0.765 | 0.293 | 2.00 | 0.59 |
| **Baseline Total Toxicity Burden** | 1.043 | 1.012 | 1.07 | 0.0069 |
| HR = Hazard Ratio, CI = Confidence Interval, RT = Radiotherapy,<br>Pho = Photon, Pro = Proton |  |  |  |  |

| **Predictors of Any ≥Grade 3 Toxicity: Non-CNS Model** |  |  |  |  |
| --- | --- | --- | --- | --- |
| **Characteristic** | **HR** | **Lower CI** | **Upper CI** | **p-Value** |
| **Body site** |  |  |  |  |
| Abdomen & Chest |  |  |  |  |
| Musculoskeletal & Head & Neck | 2.93 | 0.82 | 10.49 | 0.098 |
| Pelvis | 4.25 | 1.08 | 16.72 | 0.038 |
| **Baseline Total Toxicity Burden** | 1.1 | 1.04 | 1.16 | 0.0015 |
| HR = Hazard Ratio, CI = Confidence Interval, RT = Radiotherapy |  |  |  |  |

## DISCUSSION

The Phase IV SJPROTON1 trial has enrolled the largest pediatric oncology cohort to date to be treated at a single institution with PBS–PT methods. The participants will be monitored for 10 years thereafter via a rigorous follow-up evaluation schedule and multidisciplinary systematic event reviews. Although the use of PBS-PT for children with cancer was rapidly accepted by practitioners, health care systems, and insurance companies, there is still concern about the potential for adverse effects.^1, 20^ Unfortunately, despite decades of referrals to major tertiary care centers, the literature contains a paucity of objective data on the adverse effects of PT in children; most reports include follow-up and selection bias.^22^

Herein we demonstrated that clinically significant acute toxicity events arise in more than 1 of 4 children with CNS tumors and in more than 1 of 5 of those with non-CNS tumors treated with PBS-PT. Furthermore, as many as 1 of 6 to 7 children require additional invasive procedures and/or hospitalization to treat PBS-PT–attributable toxicity. This risk is comparable to that seen in previous studies at our institution^23^, though large proton therapy referral centers have reported lower rates of injury.^22, 24^

We determined the CI of key PBS-PT–attributable adverse events across a range of ages, diseases, and multimodality therapies. Like previous reports, age, posterior fossa tumor location, and the use of PBS-PT for retreatment were key risk factors for CNS necrosis.^25-27^ Our experience demonstrates the potential influence of the region treated and multimodality therapy and the potential contribution of treatment technique. Variabilities in preexisting injury, tumor location, and treatment delivery have precluded smaller disease-focused clinical studies from demonstrating a definitive relationship between treatment technique and increased propensity for end-of-range–related injury.^28, 29^ The present analyses lend credence to these concerns, and providers should consider the potential contribution of linear energy transfer in the planning process.^30^

The early vasculopathy events were all low grade, and their timing was earlier than times reported in the literature.^31, 32^ Evaluations of children who experienced vasculopathy after photon therapy mostly have been conducted using registry survivorship data, which may or may not prespecify the type or timing of follow-up evaluations.^33^ Matched photon/proton comparison studies with comparable evaluation schedules are warranted.^34^

We found few gastrointestinal toxicity events in our cohort; this most likely reflected our preference to avoid delivery uncertainties inherent to abdomino-pelvic tumors and participant-selection bias.^35^ Musculoskeletal events were also infrequent; however, we found a high proportion of vertebral fractures reflecting the potential need to modulate the PBS-PT dose delivered to the vertebral body in cases requiring CSI. These events occurred early and were frequently preceded by prolonged steroid use and preexisting poor bone health when PBS-PT doses were lower than those typically attributable to PBS-PT-induced fracture events.^36^ The transition to PBS-PT has prompted specialty working groups to re-envision the impact of PBS-PT on bone growth and injury in children.^37, 38^

Radiotherapy-attributable hospitalizations and procedures are poorly described in the literature. Perioperative complications are often defined relative to a limited period. We chose to extend the window, given the potential for toxicity events to occur years after treatment.^39^ The present event estimates reflect our broad participant population, and many of the events were only partially attributable to PBS-PT. Refinement in standardized reporting guidelines for PBS-PT–attributable events are needed and will most likely improve surveillance and management guidelines for clinical trials. Finally, we cannot define contraindications for the use of conformal PBS-PT in any disease context, though high-risk clinical situations accompanied by poor prognoses may cause providers to forgo its application.

## CONCLUSION

This first report from our single-institution Phase IV study highlights the potential adverse events related to PBS-PT, defines benchmarks for future clinical studies, and illustrates the need for rigorous post–PBS-PT surveillance for acute and late effects. Further improvement in evaluation, attribution, and reporting standards may improve the detection, classification, and management of PBS-PT–attributable toxicities. With longer follow-up of our cohort, we will determine whether PBS-PT has comparably efficacy, and/or less toxic relative to photon therapy in children with cancer.

## Supporting information

SJPROTON1 Protocol

## Data Availability

All data produced in the present study are available upon reasonable request to the authors

## Supplementary Table and Figure Legends

**Supplementary Table 1. Extended CNS Patient Characteristics**. CNS = Central Nervous System, ALL = Acute Lymphoblastic Leukemia, ATRT = Atypical Teratoid Rhabdoid Tumor, NOS = Not Otherwise Specified, PNET = Primitive Neuroectodermal Tumor, WHO = World Health Organization, RT = Radiotherapy, IQR = Interquartile Range, N = Number.

**Supplementary Table 2. Extended Non-CNS Patient Characteristics**. H&N = Head and Neck, MSK = Musculoskeletal, AML = Acute Myeloid Leukemia, DSCRT = Desmoplastic Small Round Cell Tumor, MPNST = Malignant Peripheral Nerve Sheath Tumor, CNS = Central Nervous System, NOS = Not Otherwise Specified, dx = Diagnosis, RT = Radiotherapy, IQR = Interquartile Range, N = Number.

**Supplementary Table 3. Combined Univariate and Multivariate Analysis Stratified by CNS and Non-CNS Cohorts**. RT = Radiotherapy, HR = Hazard Ratio, CI = Confidence Interval, Dz= Disease, H&N = Head and Neck, MSK = Musculoskeletal.

**Supplementary Figure S1. SJPROTON1 Study Design**. Providers selected eligible patients for mixed photon/PBS-PT or PBS-PT only. Following enrollment, baseline characteristics of the patient, disease, treatment, and toxicity were coded. Patients were assessed for the primary, secondary, and exploratory endpoints at given timepoints after completion of PBS-PT.

**Supplementary Figure S2. Protocol Quality Assurance**. Heatmap of assessment completion and evaluation metrics by timepoint. Yellow represents 100% completion, and purple represents 0% completion.

**Supplementary Figure S3. Nonanatomic CNS Deficits**. (A) Nonanatomic CNS deficit types by association with other toxicities, the associated toxicity type (when present), and the likelihood of attribution to radiotherapy. (B) Proportion of each type of nonanatomic CNS deficit and CTCAE grade by treatment exposure and potential attribution to radiotherapy. (C) Interventions and outcomes for nonanatomic CNS deficits following radiotherapy by medical intervention and toxicity type, resolution and toxicity type, medical intervention and grade, and resolution and grade. (D) Time to nonanatomic CNS deficits from the end of radiotherapy in months (Time 0). (E) CI of the resolution of nonanatomic CNS deficits from the start of the toxicity (Time 0).

**Supplementary Figure S4. Augmentation of Prespecified Toxicities in the Secondary Objectives**. The SJPROTON1 patient list was used to define a set of records and documents within the electronic medical record that could be queried within a set timeline for prespecified search terms. Each search underwent rigorous validation, review, attribution, and coding within the primary MOSAIQ database prior to inclusion in the present analysis. Abbreviations: CNS RN, central nervous system radionecrosis; ORN, osteonecrosis; TIA, transient ischemic attack

**Supplementary Figure S5. SJPROTON1 Attribution Review Process**. Each toxicity prespecified in the secondary objectives underwent review by multiple providers to select the potential conditions for PBS-PT attribution. After the review and annotation, each provider gave an overall score that represented the likelihood that the toxicity event was solely attributable to PBS-PT.

**Supplementary Figure S6. Total Toxicity Burden Timelines Across Patients with CNS Tumors, Solid Tumors, Lymphoma, or Leukemia**. The weighted sum of toxicities that arose in each patient, from baseline to 1 year after completion of PBS-PT, are presented. Patients were stratified by disease group.

**Supplementary Figure S7. Summary Statistics for the Total Toxicity Burden at Each Timepoint Across Patients Grouped by Disease**. Boxplots at each timepoint illustrate the median, interquartile range, and outlier values for all patients in each disease group.

**Supplementary Figure S8. Toxicity (≥ Grade 3) by CNS Disease Group**. CI of ≥ grade 3 toxicity in patients with CNS tumors grouped by their disease.

**Supplementary Figure S9. Toxicity (≥ Grade 3) by Solid Tumor Disease Group**. CI of ≥ grade 3 toxicity in patients with solid tumors grouped by their disease.

**Supplementary Figure S10. Frequency of All Toxicity Events (Grades 1-4) Across Patients with Solid Tumors by Disease Group and Toxicity Class**. Each row in the heatmap represents a single patient’s cumulative frequency of the toxicity event and grade across all trial timepoints. The most common toxicities for each class are represented in column headers, with the contributing toxicity types within that class being shown below the figure. The severity of the grades observed for each patient are shown as a colorimetric heatmap in which higher grade toxicities are progressively darker orange.

**Supplementary Figure S11. Frequency of All Toxicity Events (Grade 1-4) Across Patients with Leukemia/Lymphoma by Disease Group and Toxicity Class**. In the heatmap, each row presents a single patient’s cumulative frequency of the toxicity event and grade across all trial timepoints. The most common toxicities for each class are represented in column headers, and the contributing toxicity types within that class are shown below the figure. The severity of the grades observed for each patient are shown as a colorimetric heatmap in which higher grade toxicities are progressively darker orange.

**Supplementary Figure S12. CNS Necrosis Analysis**. (A) CTCAE grade by Disease Group, Attribution, and Prior radiotherapy use. (B) Number of bevacizumab doses by presence or absence of CNS necrosis symptoms. (C) Rate of symptom and radiographic resolution of CNS necrosis by bevacizumab, hyperbaric oxygen, and steroid utilization. (D) Probability density scatter plot of composite dose (in GyRBE) and time to toxicity. (E) Probability density scatter plot of the time to symptom and radiographic resolution of CNS necrosis. (F) Probability density scatter plot of the time to CNS necrosis and radiographic resolution. (G) Probability density scatter plot of the time to CNS necrosis and symptom resolution. (H) CI of the time to radiographic resolution among all CNS necrosis cases. (I) Forest plot of the univariate Cox proportional hazards model for use of bevacizumab and the hazard of radiographic resolution of CNS necrosis. (J) CI of the time to radiographic resolution of CNS necrosis by use of bevacizumab. (K) Time to CNS necrosis symptom resolution. (L) Forest plot of univariate Cox proportional hazards models evaluating the impact of Sawya grade and chemotherapy exposure on the hazard for CNS necrosis symptom resolution. (M) CI of the time to symptom resolution by Sawya grade.

**Supplementary Figure S13. Vasculopathy Analysis**. (A) Pretherapy exposures. Vasculopathy CTCAE grade by disease group, presence of associated symptoms and NF1 genetic background status in the observed vasculopathy cases. (B) CI of vasculopathy events following radiotherapy among all vasculopathy cases in months. (C) Interventions and outcomes across vasculopathy event types by CTCAE grade, medical intervention, and surgical intervention. (D) CI of vasculopathy by probable attribution to radiotherapy.

**Supplementary Figure S14. Hearing Analysis**. (A) Toxicity grade, therapy-related exposure, and potential attribution to radiotherapy. (B) CTCAE grade by intervention. (C) Proportion of hearing loss cases that were corrected. (D) CTCAE grade of hearing loss by disease category. (E) Time to toxicity event. (F) Probability density scatter plot of composite dose (in GyRBE) and time to toxicity.

**Supplementary Figure S15. Fracture and Osteoradionecrosis**. (A) Pretherapy exposures affecting the risk of fracture and osteoradionecrosis and grade of the event. (B) Treatment exposures affecting bone injury type and toxicity grade. (C) Interventions and outcomes for fracture and osteoradionecrosis events by type, grade, and presence of symptoms. (D) Probability density scatter plot of composite dose (in GyRBE) and time to toxicity. (E) CI of fracture and osteoradionecrosis among events. CI of symptomatic (F) and radiographic (G) resolution of fracture and osteoradionecrosis.

**Supplementary Figure S16. Nutrition and Gastrointestinal-related Toxicity**. (A) Toxicity type by treatment exposure and outcome. (B) Toxicity grade by disease category. (C) CI of each toxicity event among all nutrition and gastrointestinal toxicity events. (D) CI of the resolution of each toxicity event among all nutrition and gastrointestinal toxicity events.

## Supplementary Data

**Supplementary Data 1. Toxicity Categories Supplementary**

**Supplementary Data 2. Swimmer Plot Data Supplementary**

**Supplementary Data 3. Hospitalization and Procedures Supplementary**

**Supplementary Data 4. Diagnoses and Disease Categories Supplementary**

**Supplementary Data 5. Total Toxicity Burden Supplementary Data 6. Vasculopathy**

**Supplementary Data 7. CNS Necrosis Supplementary**

**Supplementary Data 8. CNS Deficit Anatomic Supplementary**

**Supplementary Data 9. CNS Deficit Non-Anatomic**

**Supplementary Data 10. Fracture and Osteoradionecrosis Supplementary**

**Supplementary Data 11. Nutrition and GI**

## References

1. Merchant TE. Clinical controversies: proton therapy for pediatric tumors. Semin Radiat Oncol. Apr 2013;23(2):97–108. doi:10.1016/j.semradonc.2012.11.008

2. Shen CJ, Hu C, Ladra MM, Narang AK, Pollack CE, Terezakis SA. Socioeconomic factors affect the selection of proton radiation therapy for children. Cancer. Oct 15 2017;123(20):4048–4056. doi:10.1002/cncr.30849

3. DeLaney TF, Yock TI, Paganetti H. Assessing second cancer risk after primary cancer treatment with photon or proton radiotherapy. Cancer. Aug 1 2020;126(15):3397–3399. doi:10.1002/cncr.32936

4. Eaton BR, Goldberg S, Tarbell NJ, et al. Long-term health-related quality of life in pediatric brain tumor survivors receiving proton radiotherapy at <4 years of age. Neuro Oncol. Sep 29 2020;22(9):1379–1387. doi:10.1093/neuonc/noaa042

5. Vatner RE, Niemierko A, Misra M, et al. Endocrine Deficiency As a Function of Radiation Dose to the Hypothalamus and Pituitary in Pediatric and Young Adult Patients With Brain Tumors. J Clin Oncol. Oct 1 2018;36(28):2854–2862. doi:10.1200/JCO.2018.78.1492

6. Giantsoudi D, Seco J, Eaton BR, et al. Evaluating Intensity Modulated Proton Therapy Relative to Passive Scattering Proton Therapy for Increased Vertebral Column Sparing in Craniospinal Irradiation in Growing Pediatric Patients. Int J Radiat Oncol Biol Phys. May 1 2017;98(1):37–46. doi:10.1016/j.ijrobp.2017.01.226

7. Yock TI, Yeap BY, Ebb DH, et al. Long-term toxic effects of proton radiotherapy for paediatric medulloblastoma: a phase 2 single-arm study. The Lancet Oncology. Mar 2016;17(3):287–298. doi:10.1016/S1470-2045(15)00167-9

8. Eaton BR, Esiashvili N, Kim S, et al. Endocrine outcomes with proton and photon radiotherapy for standard risk medulloblastoma. Neuro Oncol. Jun 2016;18(6):881–7. doi:10.1093/neuonc/nov302

9. Geng C, Moteabbed M, Xie Y, Schuemann J, Yock T, Paganetti H. Assessing the radiation-induced second cancer risk in proton therapy for pediatric brain tumors: the impact of employing a patient-specific aperture in pencil beam scanning. Phys Med Biol. Jan 7 2016;61(1):12–22. doi:10.1088/0031-9155/61/1/12

10. Kahalley LS, Ris MD, Grosshans DR, et al. Comparing Intelligence Quotient Change After Treatment With Proton Versus Photon Radiation Therapy for Pediatric Brain Tumors. J Clin Oncol. Apr 1 2016;34(10):1043–9. doi:10.1200/JCO.2015.62.1383

11. Vern-Gross TZ. Assessment of LET-Based Models and Changes Seen on Brain Imaging in Pediatric Patients Following Proton Therapy for Primary Central Nervous System or Base of Skull Tumors: ClinicalTrials.gov Identifier: NCT04296617. https://clinicaltrials.gov/ct2/show/NCT04296617?term=04296617&draw=2&rank=1. 2019;

12. Hua C-H. Estimating Setup Uncertainty in Pediatric Proton Therapy Using Volumetric Images: ClinicalTrials.gov Identifier: NCT04125095. https://clinicaltrials.gov/ct2/show/NCT04125095?term=04125095&draw=2&rank=1. 2019.

13. Yock TI, Tarbell NJ. Technology insight: Proton beam radiotherapy for treatment in pediatric brain tumors. Nat Clin Pract Oncol. Dec 2004;1(2):97-103; quiz 1 p following 111. doi:10.1038/ncponc0090

14. MacDonald S. Proton Craniospinal Irradiation With Bone Sparing to Decrease Growth Decrement From Radiation: ClinicalTrials.gov Identifier: NCT03281889. https://clinicaltrials.gov/ct2/show/NCT03281889?term=03281889&draw=2&rank=1. 2017.

15. Lim DH. Registry for Analysis of Quality of Life, Normal Organ Toxicity and Survival of Pediatric Patients Treated With Proton Therapy: ClinicalTrials.gov Identifier: NCT02644993. https://clinicaltrials.gov/ct2/results?cond=&term=02644993&cntry=&state=&cuity=&dist=. 2016;

16. Eaton BR. Vertebral Body Sparing Craniospinal Irradiation for Pediatric Patients With Cancer of the Central Nervous System: ClinicalTrials.gov Identifier: NCT04276194. https://clinicaltrials.gov/ct2/show/NCT04276194?term=04276194&draw=2&rank=1. 2020;

17. Lawell MP, Indelicato DJ, Paulino AC, et al. An open invitation to join the Pediatric Proton/Photon Consortium Registry to standardize data collection in pediatric radiation oncology. Br J Radiol. Mar 2020;93(1107):20190673. doi:10.1259/bjr.20190673

18. Glasser SP, Salas M, Delzell E. Importance and challenges of studying marketed drugs: what is a phase IV study? Common clinical research designs, registries, and self-reporting systems. J Clin Pharmacol. Sep 2007;47(9):1074–86. doi:10.1177/0091270007304776

19. Lucas Jr JT. Evaluation of Proton Therapy in Pediatric Cancer Patients: ClinicalTrials.gov Identifier: NCT03223766. https://clinicaltrials.gov/ct2/show/NCT03223766?term=03223766&draw=2&rank=1. 2017.

20. Weber DC, Habrand JL, Hoppe BS, et al. Proton therapy for pediatric malignancies: Fact, figures and costs. A joint consensus statement from the pediatric subcommittee of PTCOG, PROS and EPTN. Radiother Oncol. Jul 2018;128(1):44–55. doi:10.1016/j.radonc.2018.05.020

21. Timmermann B, Schuck A, Niggli F, et al. Spot-scanning proton therapy for malignant soft tissue tumors in childhood: First experiences at the Paul Scherrer Institute. Int J Radiat Oncol Biol Phys. Feb 1 2007;67(2):497–504. doi:10.1016/j.ijrobp.2006.08.053

22. Schipper MJ, Taylor JM, Smith GL, Jagsi R. Comparing long-term treatment-associated toxicities in cancer patients: approaches, caveats, and recommendations. Int J Radiat Oncol Biol Phys. Jun 1 2014;89(2):232–4. doi:10.1016/j.ijrobp.2014.01.030

23. Gajjar A. Risk-Adapted Therapy for Young Children With Embryonal Brain Tumors, Choroid Plexus Carcinoma, High Grade Glioma or Ependymoma: ClinicalTrials.gov Identifier: NCT00602667. https://clinicaltrials.gov/ct2/show/NCT00602667?term=00602667&draw=2&rank=1. 2008.

24. Brown AP, Barney CL, Grosshans DR, et al. Proton beam craniospinal irradiation reduces acute toxicity for adults with medulloblastoma. Int J Radiat Oncol Biol Phys. Jun 1 2013;86(2):277–84. doi:10.1016/j.ijrobp.2013.01.014

25. Indelicato DJ, Flampouri S, Rotondo RL, et al. Incidence and dosimetric parameters of pediatric brainstem toxicity following proton therapy. Acta oncologica. Oct 2014;53(10):1298–304. doi:10.3109/0284186X.2014.957414

26. Devine CA, Liu KX, Ioakeim-Ioannidou M, et al. Brainstem Injury in Pediatric Patients Receiving Posterior Fossa Photon Radiation. Int J Radiat Oncol Biol Phys. Dec 1 2019;105(5):1034–1042. doi:10.1016/j.ijrobp.2019.08.039

27. Haas-Kogan D, Indelicato D, Paganetti H, et al. National Cancer Institute Workshop on Proton Therapy for Children: Considerations Regarding Brainstem Injury. Int J Radiat Oncol Biol Phys. May 1 2018;101(1):152–168. doi:10.1016/j.ijrobp.2018.01.013

28. Giantsoudi D, Adams J, MacDonald SM, Paganetti H. Proton Treatment Techniques for Posterior Fossa Tumors: Consequences for Linear Energy Transfer and Dose-Volume Parameters for the Brainstem and Organs at Risk. Int J Radiat Oncol Biol Phys. Feb 1 2017;97(2):401–410. doi:10.1016/j.ijrobp.2016.09.042

29. Giantsoudi D, Sethi RV, Yeap BY, et al. Incidence of CNS Injury for a Cohort of 111 Patients Treated With Proton Therapy for Medulloblastoma: LET and RBE Associations for Areas of Injury. Int J Radiat Oncol Biol Phys. May 1 2016;95(1):287–296. doi:10.1016/j.ijrobp.2015.09.015

30. Unkelbach J, Botas P, Giantsoudi D, Gorissen BL, Paganetti H. Reoptimization of Intensity Modulated Proton Therapy Plans Based on Linear Energy Transfer. Int J Radiat Oncol Biol Phys. Dec 1 2016;96(5):1097–1106. doi:10.1016/j.ijrobp.2016.08.038

31. Zaorsky NG, Zhang Y, Tchelebi LT, Mackley HB, Chinchilli VM, Zacharia BE. Stroke among cancer patients. Nat Commun. Nov 15 2019;10(1):5172. doi:10.1038/s41467-019-13120-6

32. Ullrich NJ, Robertson R, Kinnamon DD, et al. Moyamoya following cranial irradiation for primary brain tumors in children. Neurology. Mar 20 2007;68(12):932–8. doi:10.1212/01.wnl.0000257095.33125.48

33. Bowers DC, Liu Y, Leisenring W, et al. Late-occurring stroke among long-term survivors of childhood leukemia and brain tumors: a report from the Childhood Cancer Survivor Study. J Clin Oncol. Nov 20 2006;24(33):5277–82. doi:10.1200/JCO.2006.07.2884

34. Baumann BC, Mitra N, Harton JG, et al. Comparative Effectiveness of Proton vs Photon Therapy as Part of Concurrent Chemoradiotherapy for Locally Advanced Cancer. JAMA Oncol. Feb 1 2020;6(2):237–246. doi:10.1001/jamaoncol.2019.4889

35. Rao AD, Chen Q, Ermoian RP, et al. Practice patterns of palliative radiation therapy in pediatric oncology patients in an international pediatric research consortium. Pediatr Blood Cancer. Nov 2017;64(11)doi:10.1002/pbc.26589

36. Krasin MJ, Constine LS, Friedman DL, Marks LB. Radiation-related treatment effects across the age spectrum: differences and similarities or what the old and young can learn from each other. Semin Radiat Oncol. Jan 2010;20(1):21–9. doi:10.1016/j.semradonc.2009.09.001

37. Hoeben BA, Carrie C, Timmermann B, et al. Management of vertebral radiotherapy dose in paediatric patients with cancer: consensus recommendations from the SIOPE radiotherapy working group. The Lancet Oncology. Mar 2019;20(3):e155–e166. doi:10.1016/S1470-2045(19)30034-8

38. Rao AD, Ladra M, Dunn E, et al. A Road Map for Important Centers of Growth in the Pediatric Skeleton to Consider During Radiation Therapy and Associated Clinical Correlates of Radiation-Induced Growth Toxicity. Int J Radiat Oncol Biol Phys. Mar 1 2019;103(3):669–679. doi:10.1016/j.ijrobp.2018.10.026

39. Abbott TEF, Fowler AJ, Pelosi P, et al. A systematic review and consensus definitions for standardised end-points in perioperative medicine: pulmonary complications. Br J Anaesth. May 2018;120(5):1066–1079. doi:10.1016/j.bja.2018.02.007

