## Supplemental Tables and Figures for "A Phase IV Trial of Proton Therapy in Children: The First Report from the SJPROTON1 Study"

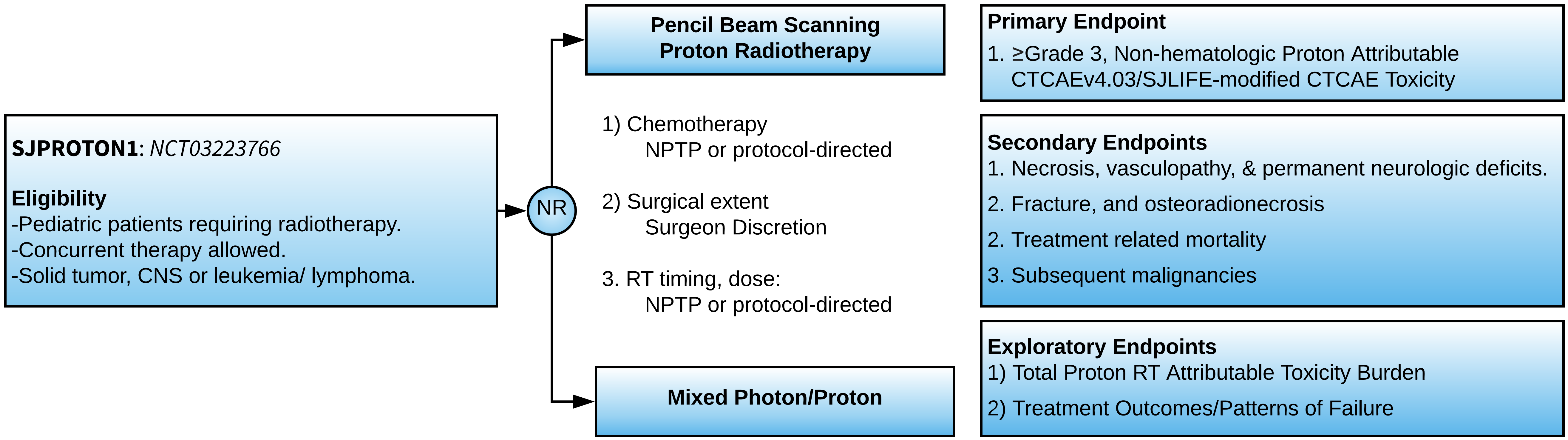
SJPROTON1: NCT03223766
Eligibility
-Pediatric patients requiring radiotherapy.
-Concurrent therapy allowed.
-Solid tumor, CNS or leukemia/ lymphoma.
NR
Pencil Beam Scanning Proton Radiotherapy
1) Chemotherapy
NPTP or protocol-directed
2) Surgical extent
Surgeon Discretion
3. RT timing, dose:
NPTP or protocol-directed
Mixed Photon/Proton
Primary Endpoint
1. ≥Grade 3, Non-hematologic Proton Attributable CTCAEv4.03/SJLIFE-modified CTCAE Toxicity
Secondary Endpoints
1. Necrosis, vasculopathy, & permanent neurologic deficits.
2. Fracture, and osteoradionecrosis
2. Treatment related mortality
3. Subsequent malignancies
Exploratory Endpoints
1) Total Proton RT Attributable Toxicity Burden
2) Treatment Outcomes/Patterns of Failure

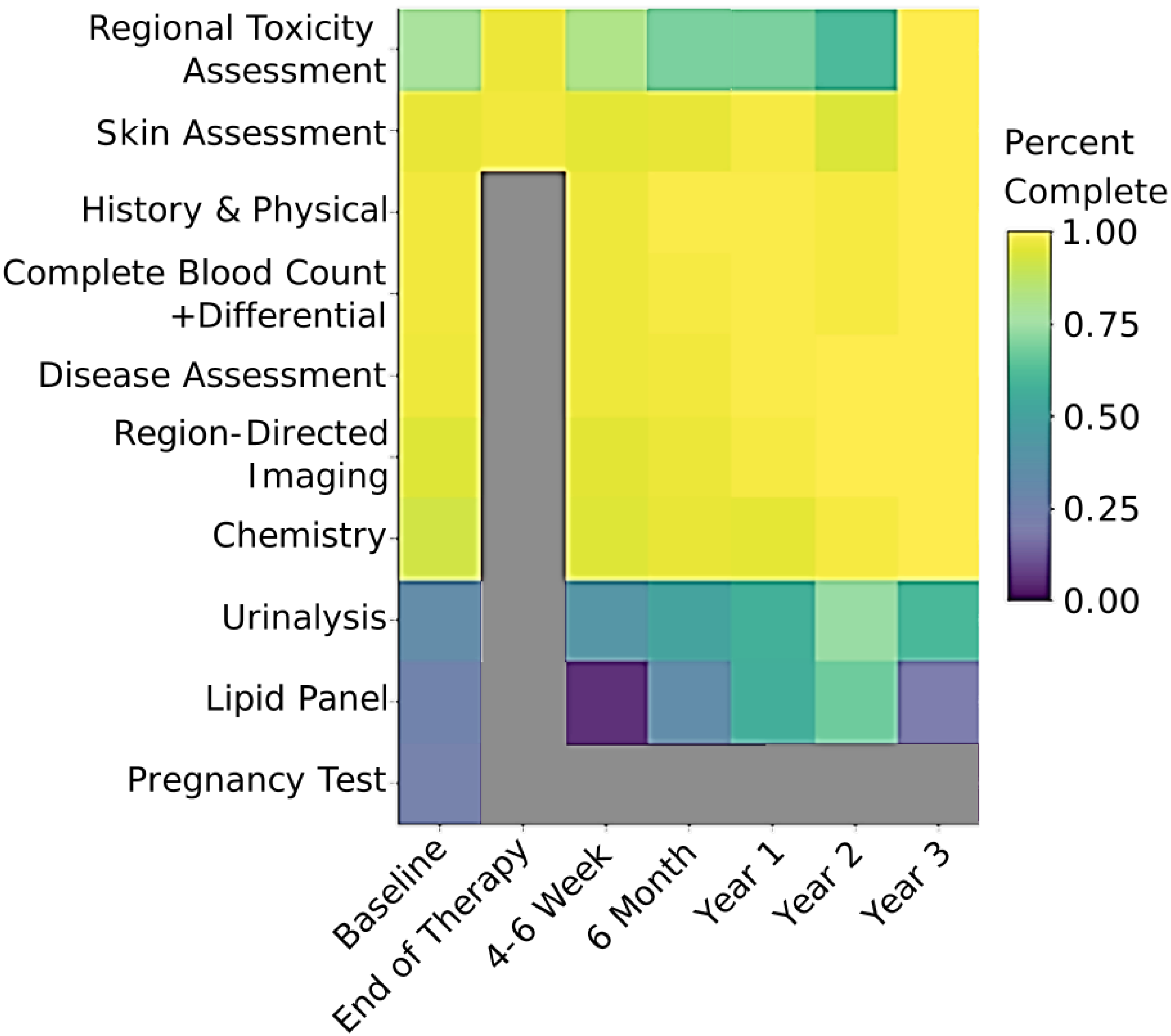

Regional Toxicity Assessment
Skin Assessment
History & Physical
Complete Blood Count +Differential
Disease Assessment
Region-Directed Imaging
Chemistry
Urinalysis
Lipid Panel
Pregnancy Test
Baseline
End of Therapy
4-6 Week
6 Month
Year 1
Year 2
Year 3
Percent Complete
1.00
0.75
0.50
0.25
0.00

A

| Characteristic | Classification | | | | | | | | |
|---|---|---|---|---|---|---|---|---|---|
| | Ataxia, N = 2 | Attention Deficit, N = 1 | Decreased level of consciousness, N = 1 | Dysphasia, N = 1 | Executive function deficit, N = 1 | Headache, N = 3 | Hypersomnia, N = 1 | Memory/executive function/processing speed deficit, N = 1 | Seizures, N = 1 |
| Associated with other toxicity | | | | | | | | | |
| Yes | 2 (100.00%) | 1 (100.00%) | 0 (0.00%) | 0 (0.00%) | 0 (0.00%) | 1 (33.33%) | 0 (0.00%) | 0 (0.00%) | 0 (0.00%) |
| No | 0 (0.00%) | 0 (0.00%) | 1 (100.00%) | 1 (100.00%) | 1 (100.00%) | 2 (66.67%) | 1 (100.00%) | 1 (100.00%) | 1 (100.00%) |
| Associated Toxicity | | | | | | | | | |
| Necrosis | 2 (100.00%) | 1 (100.00%) | 0 (0.00%) | 0 (0.00%) | 0 (0.00%) | 1 (33.33%) | 0 (0.00%) | 0 (0.00%) | 0 (0.00%) |
| None | 0 (0.00%) | 0 (0.00%) | 0 (0.00%) | 0 (0.00%) | 1 (100.00%) | 2 (66.67%) | 1 (100.00%) | 1 (100.00%) | 1 (100.00%) |
| Pseudoprogression | 0 (0.00%) | 0 (0.00%) | 1 (100.00%) | 1 (100.00%) | 0 (0.00%) | 0 (0.00%) | 0 (0.00%) | 0 (0.00%) | 0 (0.00%) |
| Attribution | | | | | | | | | |
| Likely (0.76-0.95) | 1 (50.00%) | 1 (100.00%) | 0 (0.00%) | 0 (0.00%) | 0 (0.00%) | 1 (33.33%) | 0 (0.00%) | 0 (0.00%) | 0 (0.00%) |
| Plausible (0.56-0.75) | 1 (50.00%) | 0 (0.00%) | 0 (0.00%) | 0 (0.00%) | 1 (100.00%) | 1 (33.33%) | 1 (100.00%) | 1 (100.00%) | 0 (0.00%) |
| Unassessable (0.46-0.55) | 0 (0.00%) | 0 (0.00%) | 1 (100.00%) | 1 (100.00%) | 0 (0.00%) | 1 (33.33%) | 0 (0.00%) | 0 (0.00%) | 1 (100.00%) |

[1] n (%)

B

| Characteristic | Toxicity Classification | | | | | | | | | CTCAE Grade | | |
|---|---|---|---|---|---|---|---|---|---|---|---|---|
| | Ataxia, N = 2 | Attention Deficit, N = 1 | Decreased level of consciousness N = 1 | Dysphasia N = 1 | Executive function deficit, N = 1 | Headache N = 3 | Hypersomnia N = 1 | Memory/executive function/processing speed deficit, N = 1 | Seizures N = 1 | Grade 1 N = 5 | Grade 2 N = 4 | Grade 3 N = 3 |
| Patient Modality | | | | | | | | | | | | |
| Mixed | 1 (50.00%) | 0 (0.00%) | 0 (0.00%) | 0 (0.00%) | 0 (0.00%) | 0 (0.00%) | 0 (0.00%) | 0 (0.00%) | 0 (0.00%) | 0 (0.00%) | 1 (25.00%) | 0 (0.00%) |
| Proton | 1 (50.00%) | 1 (100.00%) | 1 (100.00%) | 1 (100.00%) | 1 (100.00%) | 3 (100.00%) | 1 (100.00%) | 1 (100.00%) | 1 (100.00%) | 5 (100.00%) | 3 (75.00%) | 3 (100.00%) |
| RT Retreatment | | | | | | | | | | | | |
| Yes | 1 (50.00%) | 0 (0.00%) | 0 (0.00%) | 0 (0.00%) | 0 (0.00%) | 0 (0.00%) | 0 (0.00%) | 0 (0.00%) | 0 (0.00%) | 0 (0.00%) | 1 (25.00%) | 0 (0.00%) |
| No | 1 (50.00%) | 1 (100.00%) | 1 (100.00%) | 1 (100.00%) | 1 (100.00%) | 3 (100.00%) | 1 (100.00%) | 1 (100.00%) | 1 (100.00%) | 5 (100.00%) | 3 (75.00%) | 3 (100.00%) |
| Chemotherapy Exposure | | | | | | | | | | | | |
| Yes | 1 (50.00%) | 1 (100.00%) | 1 (100.00%) | 1 (100.00%) | 1 (100.00%) | 3 (100.00%) | 1 (100.00%) | 1 (100.00%) | 1 (100.00%) | 5 (100.00%) | 3 (75.00%) | 3 (100.00%) |
| No | 1 (50.00%) | 0 (0.00%) | 0 (0.00%) | 0 (0.00%) | 0 (0.00%) | 0 (0.00%) | 0 (0.00%) | 0 (0.00%) | 0 (0.00%) | 0 (0.00%) | 1 (25.00%) | 0 (0.00%) |
| Attribution | | | | | | | | | | | | |
| Likely (0.76-0.95) | 1 (50.00%) | 1 (100.00%) | 0 (0.00%) | 0 (0.00%) | 0 (0.00%) | 1 (33.33%) | 0 (0.00%) | 0 (0.00%) | 0 (0.00%) | 2 (40.00%) | 1 (25.00%) | 0 (0.00%) |
| Plausible (0.56-0.75) | 1 (50.00%) | 0 (0.00%) | 0 (0.00%) | 0 (0.00%) | 1 (100.00%) | 1 (33.33%) | 1 (100.00%) | 1 (100.00%) | 0 (0.00%) | 3 (60.00%) | 2 (50.00%) | 0 (0.00%) |
| Unassessable (0.46-0.55) | 0 (0.00%) | 0 (0.00%) | 1 (100.00%) | 1 (100.00%) | 0 (0.00%) | 1 (33.33%) | 0 (0.00%) | 0 (0.00%) | 1 (100.00%) | 0 (0.00%) | 1 (25.00%) | 3 (100.00%) |

[1] n (%)

C

| Characteristic | Medical Intervention | | | | | | Resolution | |
|---|---|---|---|---|---|---|---|---|
| | Analgesics N = 1 | Antiepileptics N = 1 | None N = 4 | Occupational Therapy, N = 3 | Physical therapy, N = 1 | Steroids N = 2 | Yes N = 8 | No N = 4 |
| Classification | | | | | | | | |
| Ataxia | 0 (0.00%) | 0 (0.00%) | 0 (0.00%) | 1 (33.33%) | 1 (100.00%) | 0 (0.00%) | 2 (25.00%) | 0 (0.00%) |
| Attention Deficit | 0 (0.00%) | 0 (0.00%) | 0 (0.00%) | 1 (33.33%) | 0 (0.00%) | 0 (0.00%) | 0 (0.00%) | 1 (25.00%) |
| Decreased level of consciousness | 0 (0.00%) | 0 (0.00%) | 0 (0.00%) | 0 (0.00%) | 0 (0.00%) | 1 (50.00%) | 1 (12.50%) | 0 (0.00%) |
| Dysphasia | 0 (0.00%) | 0 (0.00%) | 0 (0.00%) | 0 (0.00%) | 0 (0.00%) | 1 (50.00%) | 1 (12.50%) | 0 (0.00%) |
| Executive function deficit | 0 (0.00%) | 0 (0.00%) | 1 (25.00%) | 0 (0.00%) | 0 (0.00%) | 0 (0.00%) | 0 (0.00%) | 1 (25.00%) |
| Headache | 1 (100.00%) | 0 (0.00%) | 2 (50.00%) | 0 (0.00%) | 0 (0.00%) | 0 (0.00%) | 2 (25.00%) | 1 (25.00%) |
| Hypersomnia | 0 (0.00%) | 0 (0.00%) | 1 (25.00%) | 0 (0.00%) | 0 (0.00%) | 0 (0.00%) | 1 (12.50%) | 0 (0.00%) |
| Memory/executive function/ processing speed deficit | 0 (0.00%) | 0 (0.00%) | 0 (0.00%) | 1 (33.33%) | 0 (0.00%) | 0 (0.00%) | 0 (0.00%) | 1 (25.00%) |
| Seizures | 0 (0.00%) | 1 (100.00%) | 0 (0.00%) | 0 (0.00%) | 0 (0.00%) | 0 (0.00%) | 1 (12.50%) | 0 (0.00%) |
| CTCAE | | | | | | | | |
| 1 | 0 (0.00%) | 0 (0.00%) | 4 (100.00%) | 1 (33.33%) | 0 (0.00%) | 0 (0.00%) | 3 (37.50%) | 2 (50.00%) |
| 2 | 1 (100.00%) | 0 (0.00%) | 0 (0.00%) | 2 (66.67%) | 1 (100.00%) | 0 (0.00%) | 2 (25.00%) | 2 (50.00%) |
| 3 | 0 (0.00%) | 1 (100.00%) | 0 (0.00%) | 0 (0.00%) | 0 (0.00%) | 2 (100.00%) | 3 (37.50%) | 0 (0.00%) |

[1] n (%)

D

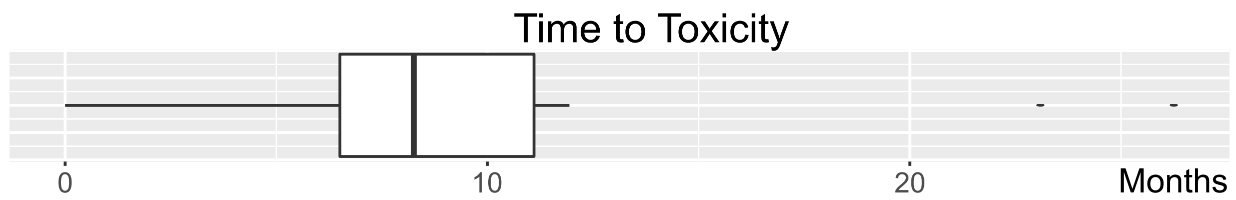


E

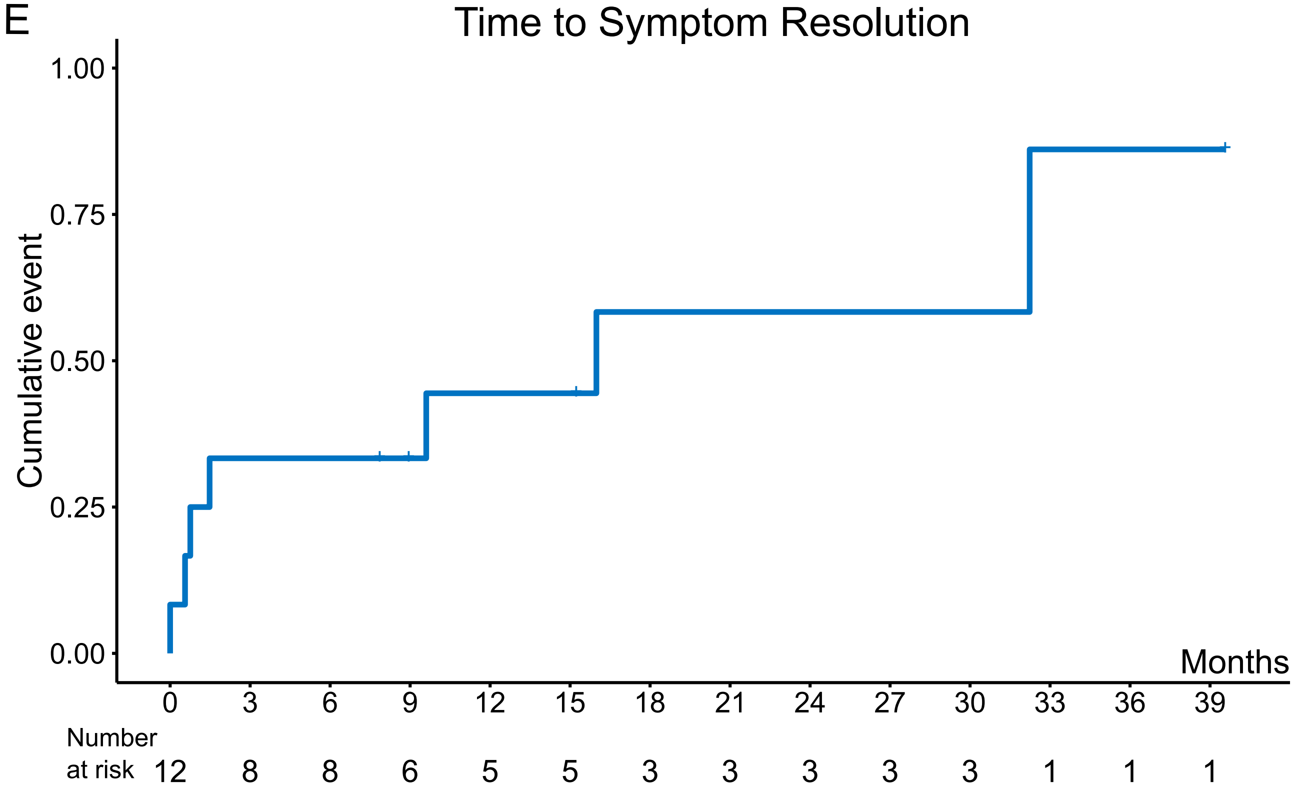

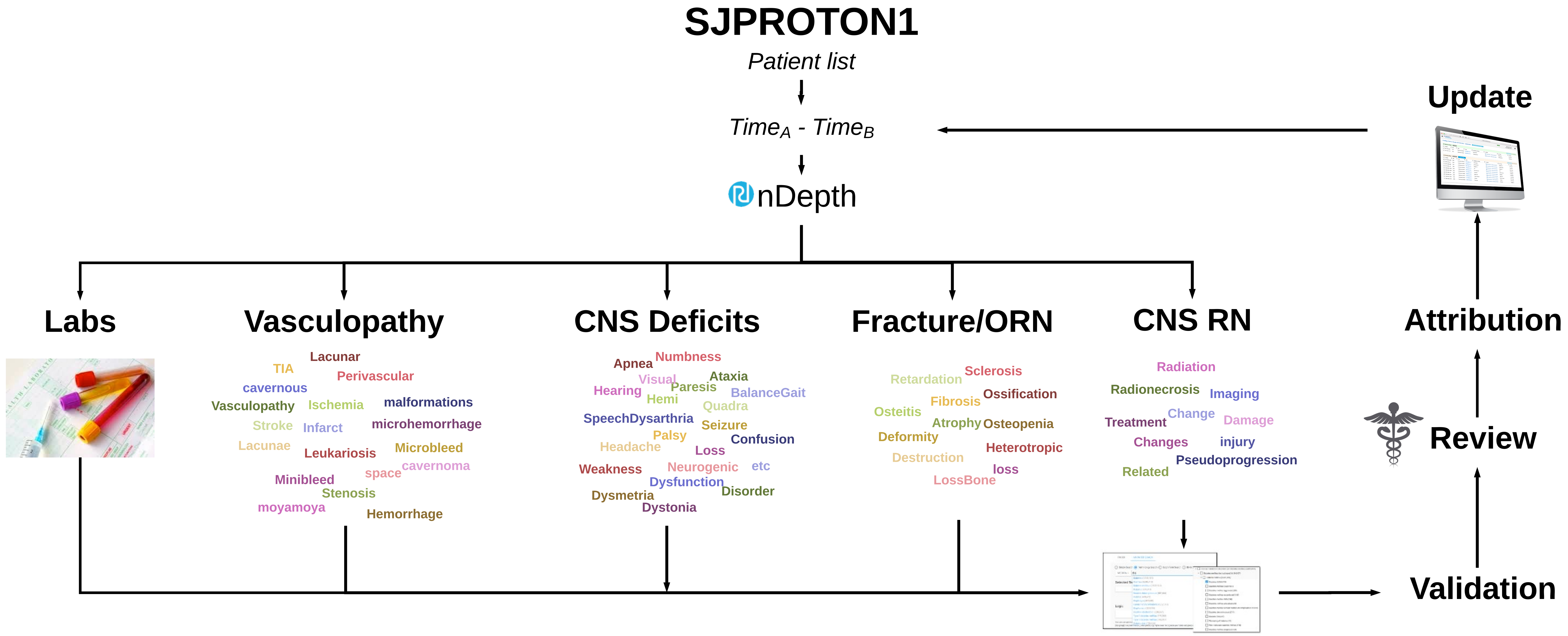

SJPROTON1
Patient list
TimeA - TimeB
nDepth
Labs
Vasculopathy
Lacunar
TIA
Perivascular
cavernous
Vasculopathy
Ischemia
malformations
Stroke
Infarct
microhemorrhage
Lacunae
Leukariosis
Microbleed
space
cavernoma
Minibleed
Stenosis
moyamoya
Hemorrhage
CNS Deficits
Apnea
Numbness
Visual
Ataxia
Hearing
Paresis
BalanceGait
Hemi
Quadra
SpeechDysarthria
Seizure
Palsy
Headache
Confusion
Loss
Weakness
Neurogenic
etc
Dysfunction
Dysmetria
Disorder
Dystonia
Fracture/ORN
Retardation
Sclerosis
Fibrosis
Ossification
Osteitis
Atrophy
Osteopenia
Deformity
Heterotropic
Destruction
loss
LossBone
CNS RN
Radiation
Radionecrosis
Imaging
Treatment
Change
Damage
Changes
injury
Pseudoprogression
Related
Update
Attribution
Review
Validation

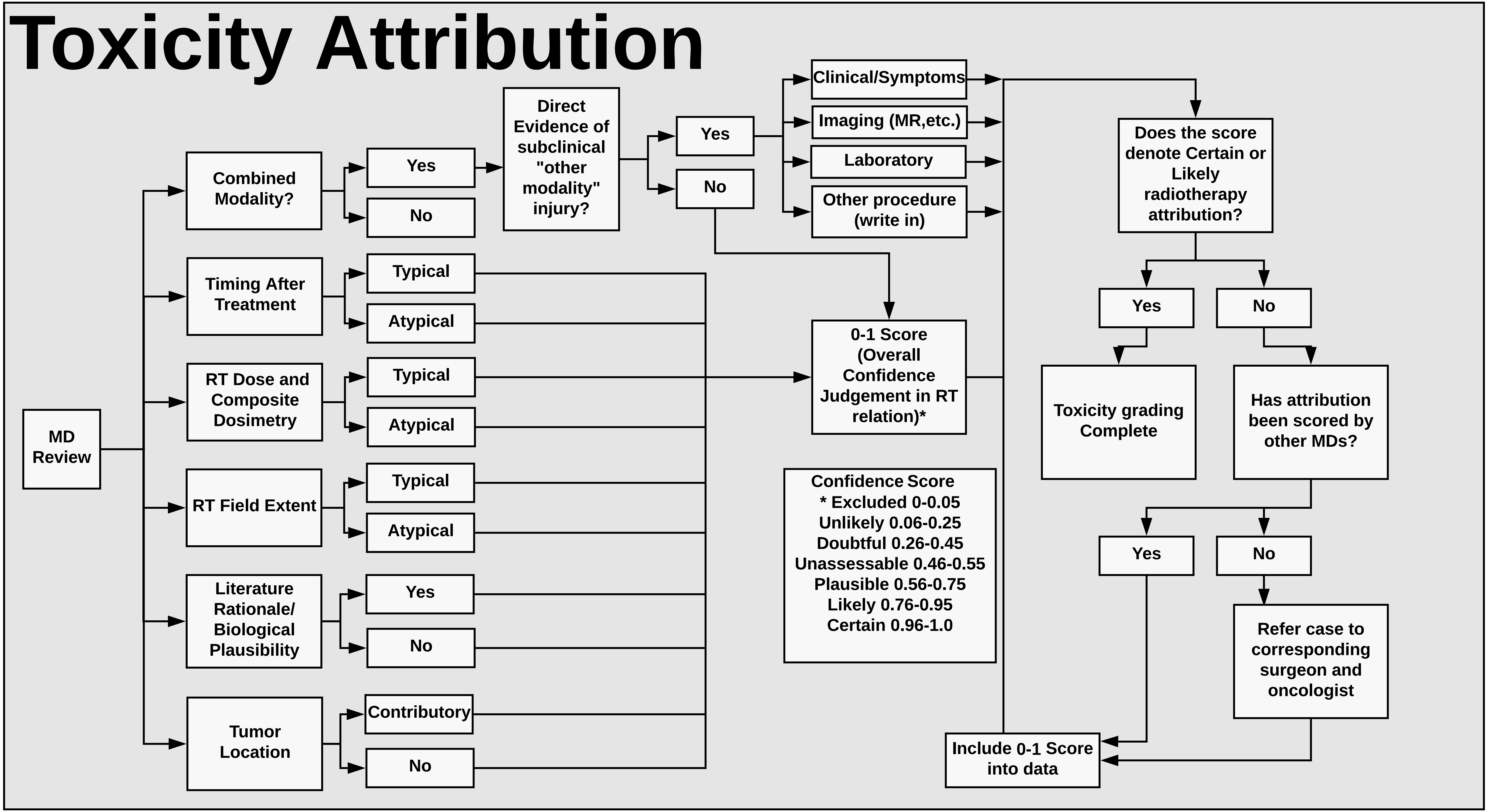
Toxicity Attribution
MD Review
Combined Modality?
Yes
No
Direct Evidence of subclinical "other modality" injury?
Yes
No
Clinical/Symptoms
Imaging (MR,etc.)
Laboratory
Other procedure (write in)
Timing After Treatment
Typical
Atypical
RT Dose and Composite Dosimetry
Typical
Atypical
RT Field Extent
Typical
Atypical
Literature Rationale/ Biological Plausibility
Yes
No
Tumor Location
Contributory
No
0-1 Score (Overall Confidence Judgement in RT relation)*
Confidence Score
* Excluded 0-0.05
Unlikely 0.06-0.25
Doubtful 0.26-0.45
Unassessable 0.46-0.55
Plausible 0.56-0.75
Likely 0.76-0.95
Certain 0.96-1.0
Does the score denote Certain or Likely radiotherapy attribution?
Yes
No
Toxicity grading Complete
Has attribution been scored by other MDs?
Yes
No
Refer case to corresponding surgeon and oncologist
Include 0-1 Score into data

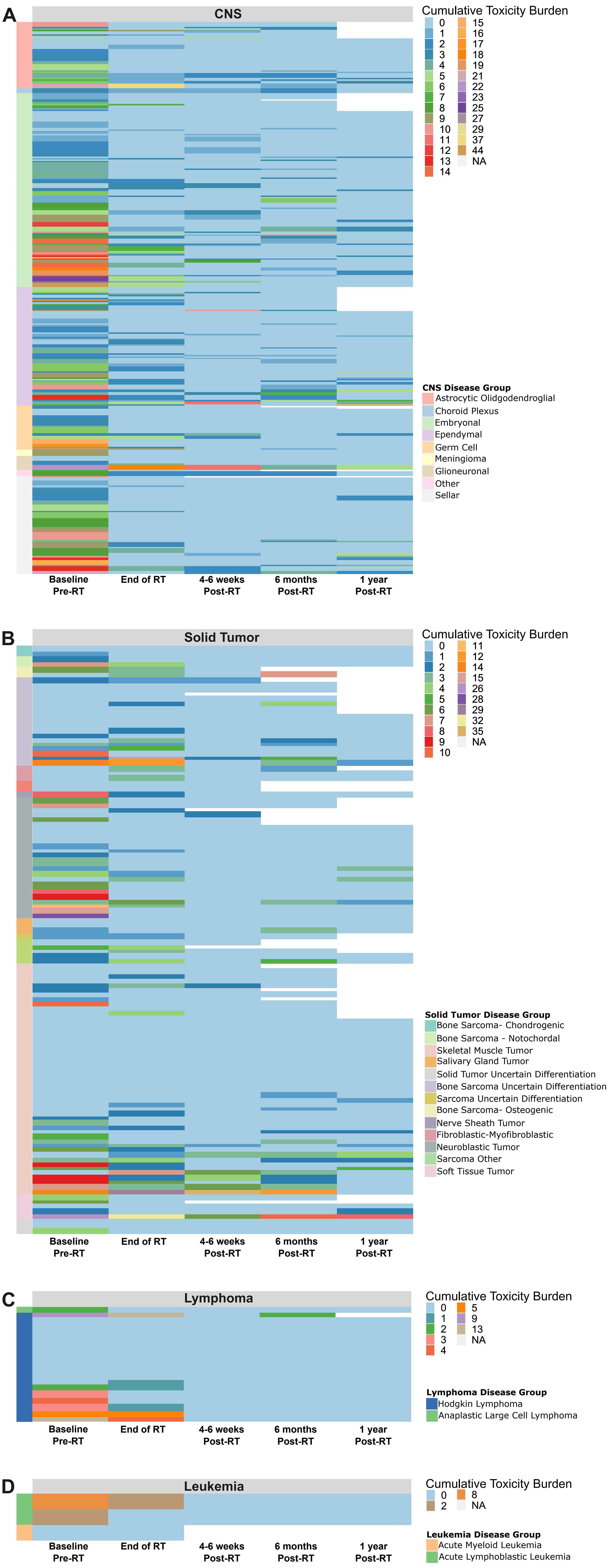
A
CNS
Cumulative Toxicity Burden
0 1 2 3 4 5 6 7 8 9 10 11 12 13 14 15 16 17 18 19 21 22 23 25 27 29 37 44 NA
CNS Disease Group
Astrocytic Olidgodendroglial
Choroid Plexus
Embryonal
Ependymal
Germ Cell
Meningioma
Glioneuronal
Other
Sellar
Baseline Pre-RT
End of RT
4-6 weeks Post-RT
6 months Post-RT
1 year Post-RT
B
Solid Tumor
Cumulative Toxicity Burden
0 1 2 3 4 5 6 7 8 9 10 11 12 14 15 26 28 29 32 35 NA
Solid Tumor Disease Group
Bone Sarcoma- Chondrogenic
Bone Sarcoma - Notochordal
Skeletal Muscle Tumor
Salivary Gland Tumor
Solid Tumor Uncertain Differentiation
Bone Sarcoma Uncertain Differentiation
Sarcoma Uncertain Differentiation
Bone Sarcoma- Osteogenic
Nerve Sheath Tumor
Fibroblastic-Myofibroblastic
Neuroblastic Tumor
Sarcoma Other
Soft Tissue Tumor
Baseline Pre-RT
End of RT
4-6 weeks Post-RT
6 months Post-RT
1 year Post-RT
C
Lymphoma
Cumulative Toxicity Burden
0 1 2 3 4 5 9 13 NA
Lymphoma Disease Group
Hodgkin Lymphoma
Anaplastic Large Cell Lymphoma
Baseline Pre-RT
End of RT
4-6 weeks Post-RT
6 months Post-RT
1 year Post-RT
D
Leukemia
Cumulative Toxicity Burden
0 2 8 NA
Leukemia Disease Group
Acute Myeloid Leukemia
Acute Lymphoblastic Leukemia
Baseline Pre-RT
End of RT
4-6 weeks Post-RT
6 months Post-RT
1 year Post-RT

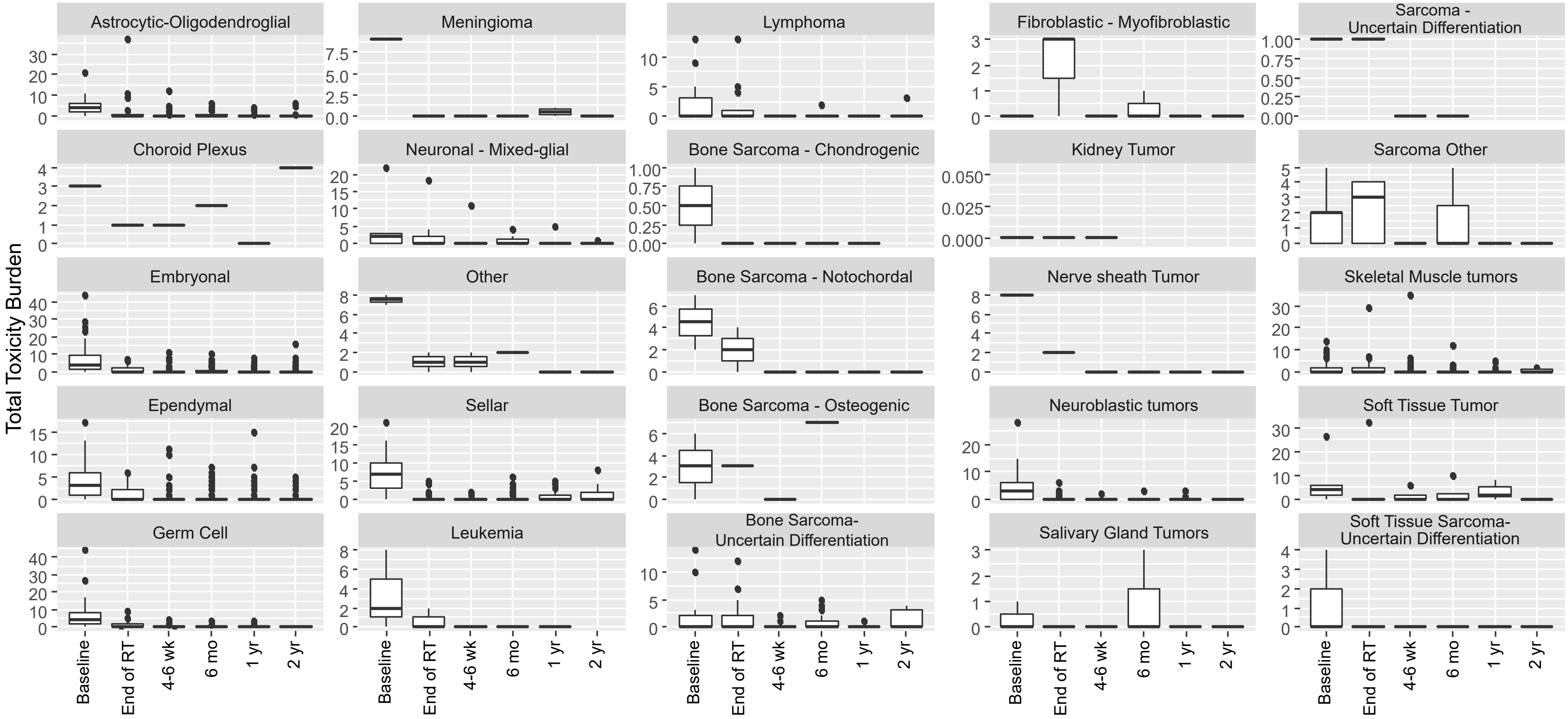

Total Toxicity Burden
Astrocytic-Oligodendroglial
Meningioma
Lymphoma
Fibroblastic - Myofibroblastic
Sarcoma - Uncertain Differentiation
Choroid Plexus
Neuronal - Mixed-glial
Bone Sarcoma - Chondrogenic
Kidney Tumor
Sarcoma Other
Embryonal
Other
Bone Sarcoma - Notochordal
Nerve sheath Tumor
Skeletal Muscle tumors
Ependymal
Sellar
Bone Sarcoma - Osteogenic
Neuroblastic tumors
Soft Tissue Tumor
Germ Cell
Leukemia
Bone Sarcoma- Uncertain Differentiation
Salivary Gland Tumors
Soft Tissue Sarcoma- Uncertain Differentiation
Baseline
End of RT
4-6 wk
6 mo
1 yr
2 yr

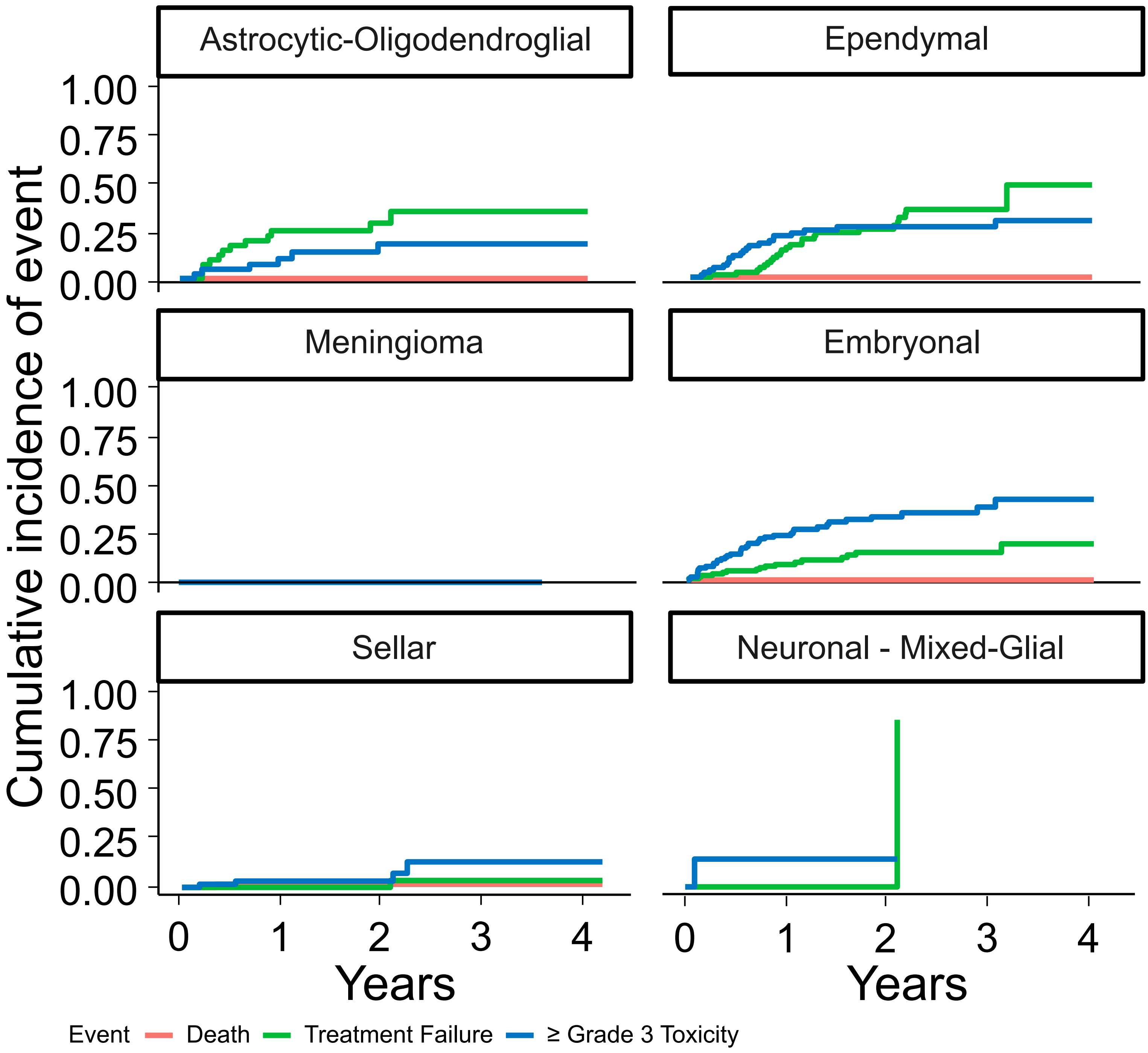

Astrocytic-Oligodendroglial
Ependymal
Meningioma
Embryonal
Sellar
Neuronal - Mixed-Glial
Cumulative incidence of event
1.00
0.75
0.50
0.25
0.00
0
1
2
3
4
Years
Event
Death
Treatment Failure
≥ Grade 3 Toxicity

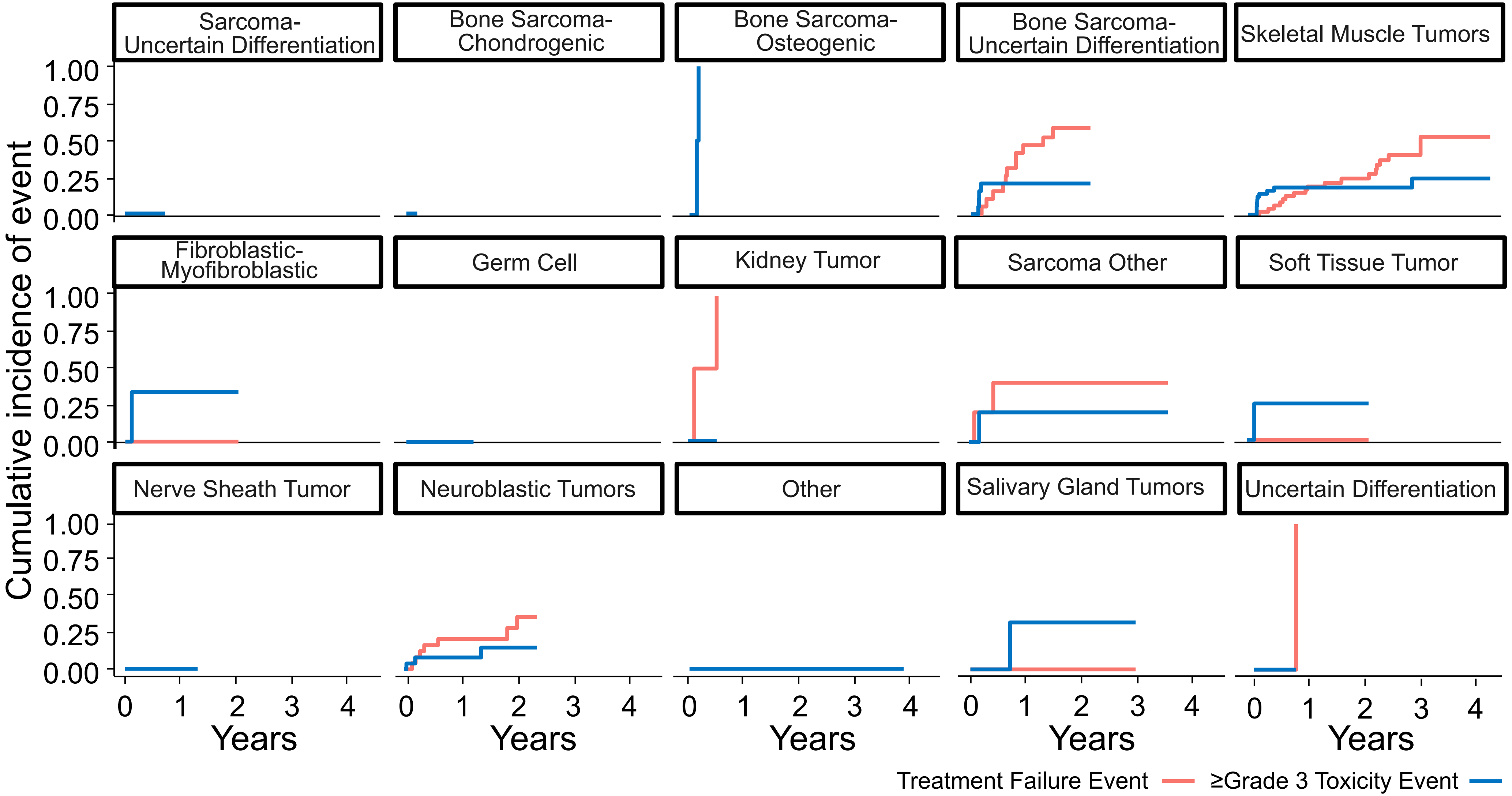
Sarcoma-Uncertain Differentiation
Bone Sarcoma-Chondrogenic
Bone Sarcoma-Osteogenic
Bone Sarcoma-Uncertain Differentiation
Skeletal Muscle Tumors
Fibroblastic-Myofibroblastic
Germ Cell
Kidney Tumor
Sarcoma Other
Soft Tissue Tumor
Nerve Sheath Tumor
Neuroblastic Tumors
Other
Salivary Gland Tumors
Uncertain Differentiation
Cumulative incidence of event
0.00
0.25
0.50
0.75
1.00
Years
0
1
2
3
4
Treatment Failure Event
≥Grade 3 Toxicity Event

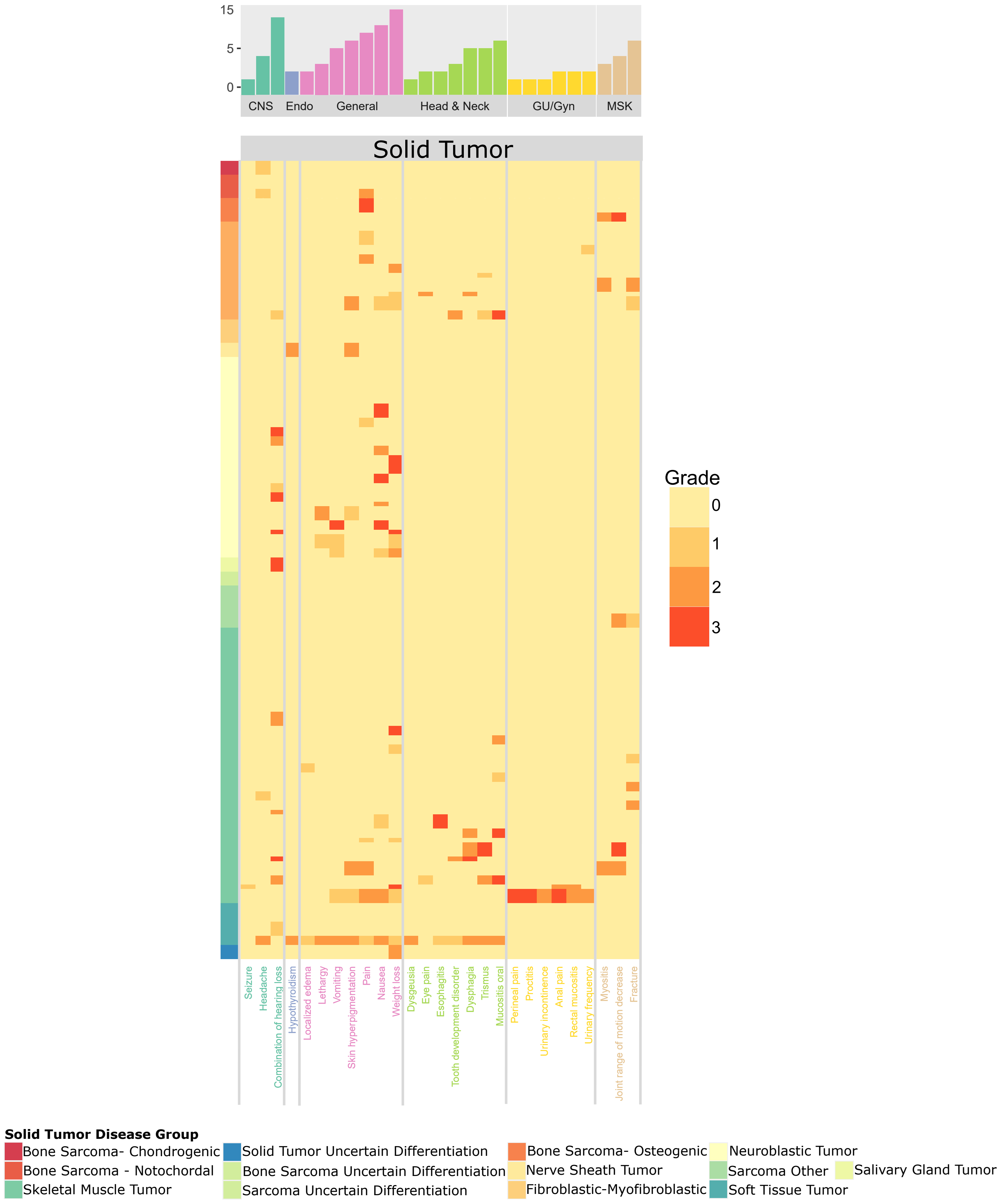
15
5
0
CNS
Endo
General
Head & Neck
GU/Gyn
MSK
Solid Tumor
Grade
0
1
2
3
Seizure
Headache
Combination of hearing loss
Hypothyroidism
Localized edema
Lethargy
Vomiting
Skin hyperpigmentation
Pain
Nausea
Weight loss
Dysgeusia
Eye pain
Esophagitis
Tooth development disorder
Dysphagia
Trismus
Mucositis oral
Perineal pain
Proctitis
Urinary incontinence
Anal pain
Rectal mucositis
Urinary frequency
Myositis
Joint range of motion decrease
Fracture
Solid Tumor Disease Group
Bone Sarcoma- Chondrogenic
Bone Sarcoma - Notochordal
Skeletal Muscle Tumor
Solid Tumor Uncertain Differentiation
Bone Sarcoma Uncertain Differentiation
Sarcoma Uncertain Differentiation
Bone Sarcoma- Osteogenic
Nerve Sheath Tumor
Fibroblastic-Myofibroblastic
Neuroblastic Tumor
Sarcoma Other
Salivary Gland Tumor
Soft Tissue Tumor

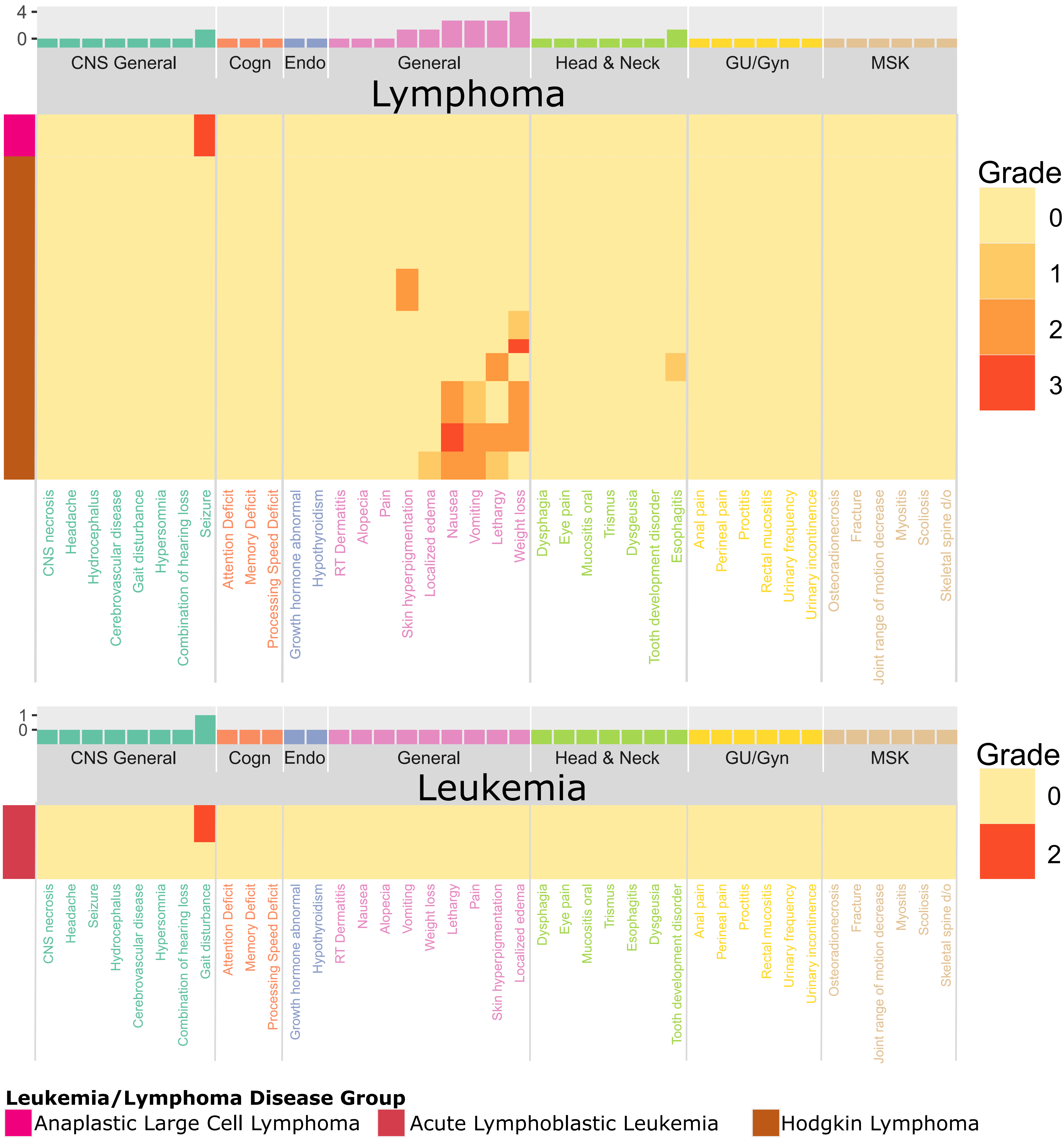
4
0
CNS General
Cogn
Endo
General
Head & Neck
GU/Gyn
MSK
Lymphoma
Grade
0
1
2
3
CNS necrosis
Headache
Hydrocephalus
Cerebrovascular disease
Gait disturbance
Hypersomnia
Combination of hearing loss
Seizure
Attention Deficit
Memory Deficit
Processing Speed Deficit
Growth hormone abnormal
Hypothyroidism
RT Dermatitis
Alopecia
Pain
Skin hyperpigmentation
Localized edema
Nausea
Vomiting
Lethargy
Weight loss
Dysphagia
Eye pain
Mucositis oral
Trismus
Dysgeusia
Tooth development disorder
Esophagitis
Anal pain
Perineal pain
Proctitis
Rectal mucositis
Urinary frequency
Urinary incontinence
Osteoradionecrosis
Fracture
Joint range of motion decrease
Myositis
Scoliosis
Skeletal spine d/o
1
0
CNS General
Cogn
Endo
General
Head & Neck
GU/Gyn
MSK
Leukemia
Grade
0
2
CNS necrosis
Headache
Seizure
Hydrocephalus
Cerebrovascular disease
Hypersomnia
Combination of hearing loss
Gait disturbance
Attention Deficit
Memory Deficit
Processing Speed Deficit
Growth hormone abnormal
Hypothyroidism
RT Dermatitis
Nausea
Alopecia
Vomiting
Weight loss
Lethargy
Pain
Skin hyperpigmentation
Localized edema
Dysphagia
Eye pain
Mucositis oral
Trismus
Esophagitis
Dysgeusia
Tooth development disorder
Anal pain
Perineal pain
Proctitis
Rectal mucositis
Urinary frequency
Urinary incontinence
Osteoradionecrosis
Fracture
Joint range of motion decrease
Myositis
Scoliosis
Skeletal spine d/o
Leukemia/Lymphoma Disease Group
Anaplastic Large Cell Lymphoma
Acute Lymphoblastic Leukemia
Hodgkin Lymphoma

A

| Characteristic | CTCAE Grade<br>Grade 1<br>N = 18 | <br>Grade 2<br>N = 2 | <br>Grade 3<br>N = 19 |
|---|---|---|---|
| Disease Categories | | | |
| CNS - Astrocytic-Oligodendroglial | 2 (11.11%) | 0 (0.00%) | 1 (5.26%) |
| CNS - Embryol | 8 (44.44%) | 0 (0.00%) | 4 (21.05%) |
| CNS - Ependymal | 5 (27.78%) | 1 (50.00%) | 13 (68.42%) |
| CNS - Meningioma | 1 (5.56%) | 0 (0.00%) | 0 (0.00%) |
| CNS - Neurol - Mixed-glial | 1 (5.56%) | 1 (50.00%) | 0 (0.00%) |
| CNS - Sellar | 1 (5.56%) | 0 (0.00%) | 1 (5.26%) |
| Attribution | | | |
| Likely (0.76-0.95) | 14 (77.78%) | 1 (50.00%) | 16 (84.21%) |
| Plausible (0.56-0.75) | 2 (11.11%) | 1 (50.00%) | 3 (15.79%) |
| Unassessable (0.46-0.55) | 2 (11.11%) | 0 (0.00%) | 0 (0.00%) |
| Prior RT | | | |
| No Prior RT | 15 (83.33%) | 1 (50.00%) | 10 (52.63%) |
| Prior Photon | 2 (11.11%) | 0 (0.00%) | 3 (15.79%) |
| Prior Proton | 1 (5.56%) | 1 (50.00%) | 6 (31.58%) |

[1] n (%)

B

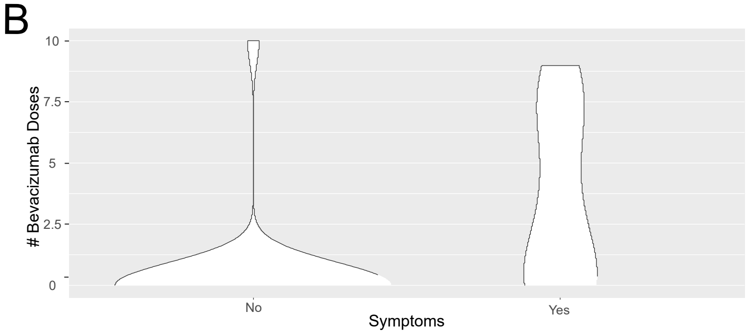


C

| Characteristic | Symptom Resolution<br>Yes<br>N = 9 | <br>No<br>N = 5 | Radiographic Resolution<br>Yes<br>N = 32 | <br>No<br>N = 3 |
|---|---|---|---|---|
| Steroids | | | | |
| Yes | 4 (44.44%) | 4 (80.00%) | 7 (21.88%) | 1 (33.33%) |
| No | 5 (55.56%) | 1 (20.00%) | 25 (78.12%) | 2 (66.67%) |
| Bevacizumab | | | | |
| Yes | 3 (33.33%) | 5 (100.00%) | 7 (21.88%) | 1 (33.33%) |
| No | 6 (66.67%) | 0 (0.00%) | 25 (78.12%) | 2 (66.67%) |
| Hyperbaric Oxygen Therapy | | | | |
| Yes | 4 (44.44%) | 0 (0.00%) | 10 (31.25%) | 0 (0.00%) |
| No | 5 (55.56%) | 5 (100.00%) | 22 (68.75%) | 3 (100.00%) |

[1] n (%)

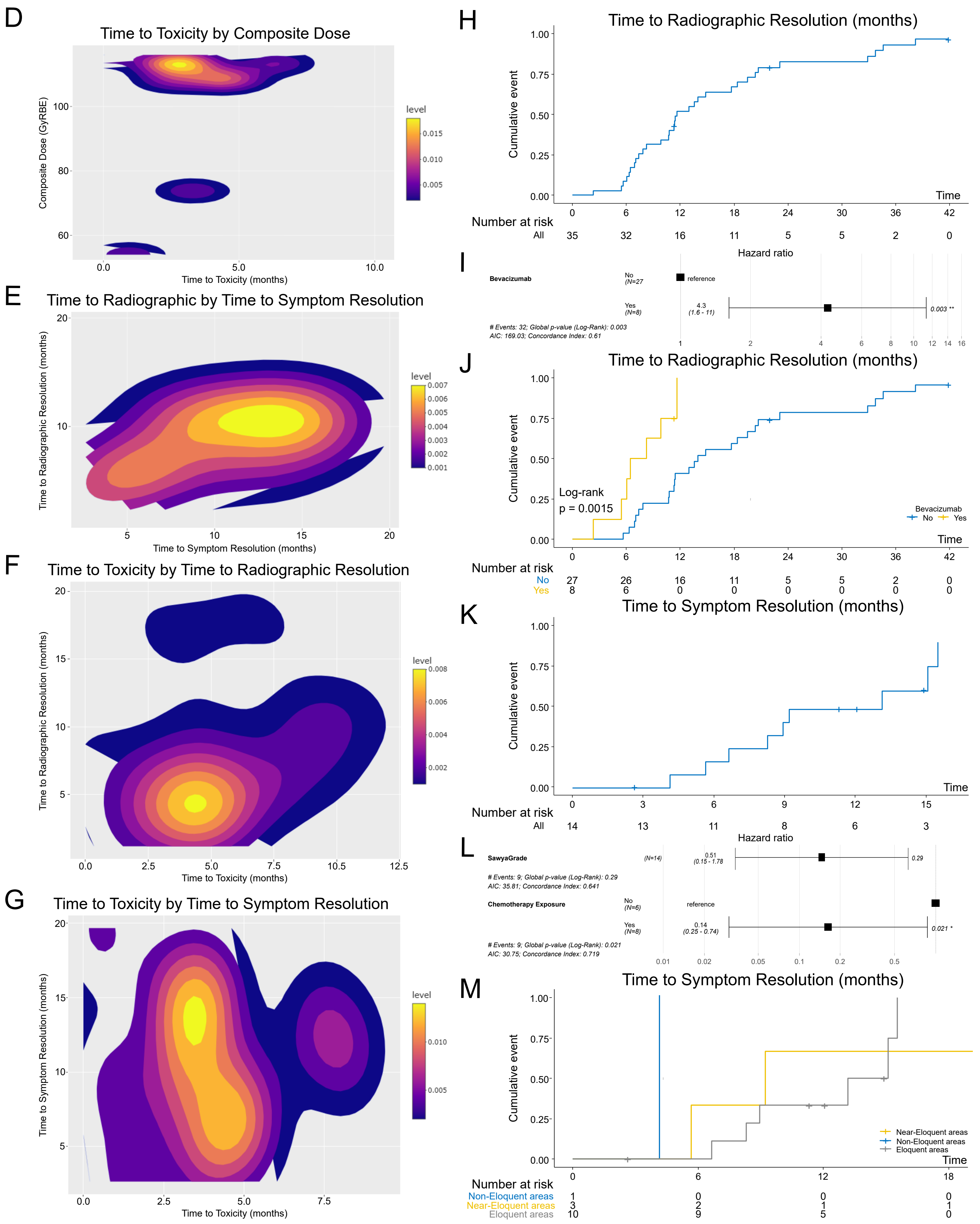

A

| Characteristic | CTCAE Grade<br>Grade 1<br>N = 25 | <br>Grade 2<br>N = 14 | <br>Grade 4<br>N = 2 |
|---|---|---|---|
| Disease Categories | | | |
| CNS - Astrocytic-Oligodendroglial | 4 (16.00%) | 3 (21.43%) | 0 (0.00%) |
| CNS - Embryonal | 2 (8.00%) | 2 (14.29%) | 1 (50.00%) |
| CNS - Ependymal | 2 (8.00%) | 1 (7.14%) | 0 (0.00%) |
| CNS - Neuronal - Mixed-glial | 1 (4.00%) | 0 (0.00%) | 0 (0.00%) |
| CNS - Sellar | 16 (64.00%) | 8 (57.14%) | 1 (50.00%) |
| Classification | | | |
| Aneurysm | 2 (8.00%) | 0 (0.00%) | 0 (0.00%) |
| Dilated perivascular space | 1 (4.00%) | 0 (0.00%) | 0 (0.00%) |
| Intracranial hemorrhage | 1 (4.00%) | 0 (0.00%) | 0 (0.00%) |
| Ischemic stroke | 0 (0.00%) | 2 (14.29%) | 0 (0.00%) |
| Lacunar infarct | 1 (4.00%) | 1 (7.14%) | 0 (0.00%) |
| Microbleed | 3 (12.00%) | 0 (0.00%) | 0 (0.00%) |
| Moyamoya | 1 (4.00%) | 0 (0.00%) | 1 (50.00%) |
| TIA | 1 (4.00%) | 1 (7.14%) | 0 (0.00%) |
| Vascular Stenosis | 15 (60.00%) | 10 (71.43%) | 1 (50.00%) |
| Symptoms? | | | |
| Yes | 1 (4.00%) | 5 (35.71%) | 1 (50.00%) |
| No | 24 (96.00%) | 9 (64.29%) | 1 (50.00%) |
| NF1 | | | |
| No | 25 (100.00%) | 14 (100.00%) | 2 (100.00%) |

[1] n (%)

B

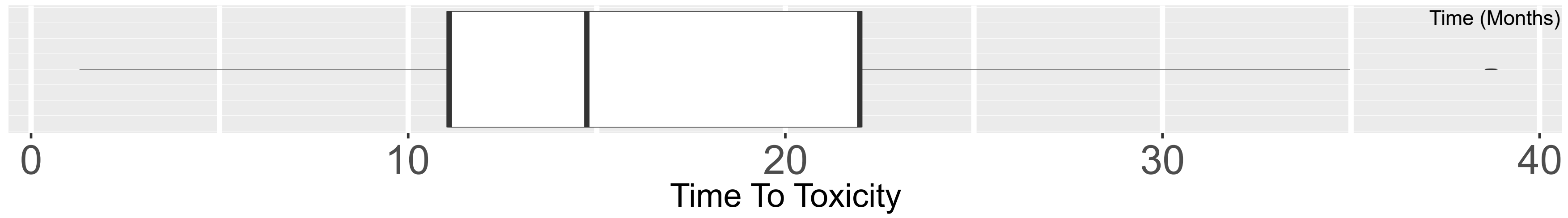


C

| Characteristic | CTCAE Grade<br>Grade 1<br>N = 25 | <br>Grade 2<br>N = 14 | <br>Grade 4<br>N = 2 | Medical Intervention<br>ASA<br>N = 13 | <br>None<br>N = 26 | <br>Steroids<br>N = 2 | Surgical Intervention<br>Indirect Vascularization<br>N = 2 | <br>None<br>N = 39 |
|---|---|---|---|---|---|---|---|---|
| Classification | | | | | | | | |
| Aneurysm | 2 (8.00%) | 0 (0.00%) | 0 (0.00%) | 0 (0.00%) | 2 (7.69%) | 0 (0.00%) | 0 (0.00%) | 2 (5.13%) |
| Dilated perivascular space | 1 (4.00%) | 0 (0.00%) | 0 (0.00%) | 0 (0.00%) | 1 (3.85%) | 0 (0.00%) | 0 (0.00%) | 1 (2.56%) |
| Intracranial hemorrhage | 1 (4.00%) | 0 (0.00%) | 0 (0.00%) | 0 (0.00%) | 1 (3.85%) | 0 (0.00%) | 0 (0.00%) | 1 (2.56%) |
| Ischemic stroke | 0 (0.00%) | 2 (14.29%) | 0 (0.00%) | 2 (15.38%) | 0 (0.00%) | 0 (0.00%) | 0 (0.00%) | 2 (5.13%) |
| Lacunar infarct | 1 (4.00%) | 1 (7.14%) | 0 (0.00%) | 0 (0.00%) | 1 (3.85%) | 1 (50.00%) | 0 (0.00%) | 2 (5.13%) |
| Microbleed | 3 (12.00%) | 0 (0.00%) | 0 (0.00%) | 0 (0.00%) | 3 (11.54%) | 0 (0.00%) | 0 (0.00%) | 3 (7.69%) |
| Moyamoya | 1 (4.00%) | 0 (0.00%) | 1 (50.00%) | 1 (7.69%) | 1 (3.85%) | 0 (0.00%) | 1 (50.00%) | 1 (2.56%) |
| TIA | 1 (4.00%) | 1 (7.14%) | 0 (0.00%) | 0 (0.00%) | 2 (7.69%) | 0 (0.00%) | 0 (0.00%) | 2 (5.13%) |
| Vascular Stenosis | 15 (60.00%) | 10 (71.43%) | 1 (50.00%) | 10 (76.92%) | 15 (57.69%) | 1 (50.00%) | 1 (50.00%) | 25 (64.10%) |

[1] n (%)

D

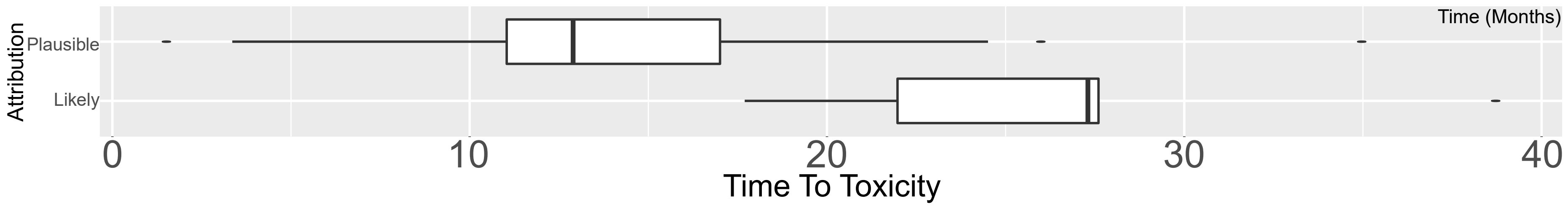

A CNS Deficit Hearing by Ear

| Characteristic | CTCAE | | | | Chang | | | | | | | SIOP | | | | | Attribution | | |
|---|---|---|---|---|---|---|---|---|---|---|---|---|---|---|---|---|---|---|---|
| | 0, N = 4[1] | 1, N = 5[1] | 2, N = 4[1] | 3, N = 37[1] | 0, N = 7[1] | 1a, N = 4[1] | 1b, N = 2[1] | 2, N = 2[1] | 2a, N = 4[1] | 2b, N = 13[1] | 3, N = 18[1] | 0, N = 4[1] | 1, N = 7[1] | 2, N = 9[1] | 3, N = 26[1] | 4, N = 4[1] | Doubtful (0.26-0.45), N = 4[1] | Likely (0.76-0.95), N = 8[1] | Plausible (0.56-0.75), N = 38[1] |
| Patient Modality | | | | | | | | | | | | | | | | | | | |
| mixed | 0 (0.00%) | 2 (40.00%) | 1 (25.00%) | 23 (62.16%) | 3 (42.86%) | 0 (0.00%) | 0 (0.00%) | 0 (0.00%) | 3 (75.00%) | 9 (69.23%) | 11 (61.11%) | 0 (0.00%) | 3 (42.86%) | 4 (44.44%) | 15 (57.69%) | 4 (100.00%) | 0 (0.00%) | 6 (75.00%) | 20 (52.63%) |
| proton | 4 (100.00%) | 3 (60.00%) | 3 (75.00%) | 14 (37.84%) | 4 (57.14%) | 4 (100.00%) | 2 (100.00%) | 2 (100.00%) | 1 (25.00%) | 4 (30.77%) | 7 (38.89%) | 4 (100.00%) | 4 (57.14%) | 5 (55.56%) | 11 (42.31%) | 0 (0.00%) | 4 (100.00%) | 2 (25.00%) | 18 (47.37%) |
| Mean Dose | NA (NA, NA) | 2,700.0 (2,340.0, 3,960.0) | 2,577.0 (1,370.8, 3,600.0) | 3,600.0 (2,340.0, 4,200.0) | 3,960.0 (3,780.0, 3,960.0) | 2,520.0 (2,272.5, 2,750.0) | 4,200.0 (4,200.0, 4,200.0) | 1,187.5 (1,004.2, 1,370.8) | 3,955.5 (3,600.0, 4,583.2) | 3,600.0 (2,340.0, 4,300.0) | 3,600.0 (2,505.0, 3,938.5) | NA (NA, NA) | 2,900.0 (2,520.0, 3,780.0) | 4,140.0 (3,600.0, 4,200.0) | 3,452.5 (2,340.0, 3,960.0) | 3,900.0 (3,285.0, 4,200.0) | 1,187.5 (615.8, 1,915.5) | 3,600.0 (3,600.0, 4,327.5) | 3,600.0 (2,340.0, 4,140.0) |
| Unknown | 4 | 0 | 0 | 0 | 4 | 0 | 0 | 0 | 0 | 0 | 0 | 4 | 0 | 0 | 0 | 0 | 0 | 1 | 3 |
| Chemotherapy_Exposure | | | | | | | | | | | | | | | | | | | |
| Yes | 3 (75.00%) | 4 (80.00%) | 4 (100.00%) | 35 (94.59%) | 6 (85.71%) | 3 (75.00%) | 2 (100.00%) | 2 (100.00%) | 4 (100.00%) | 13 (100.00%) | 16 (88.89%) | 3 (75.00%) | 6 (85.71%) | 9 (100.00%) | 24 (92.31%) | 4 (100.00%) | 2 (50.00%) | 8 (100.00%) | 36 (94.74%) |
| No | 1 (25.00%) | 1 (20.00%) | 0 (0.00%) | 2 (5.41%) | 1 (14.29%) | 1 (25.00%) | 0 (0.00%) | 0 (0.00%) | 0 (0.00%) | 0 (0.00%) | 2 (11.11%) | 1 (25.00%) | 1 (14.29%) | 0 (0.00%) | 2 (7.69%) | 0 (0.00%) | 2 (50.00%) | 0 (0.00%) | 2 (5.26%) |
| Attribution | | | | | | | | | | | | | | | | | | | |
| Doubtful (0.26-0.45) | 0 (0.00%) | 0 (0.00%) | 2 (50.00%) | 2 (5.41%) | 0 (0.00%) | 0 (0.00%) | 0 (0.00%) | 2 (100.00%) | 0 (0.00%) | 0 (0.00%) | 2 (11.11%) | 0 (0.00%) | 0 (0.00%) | 2 (22.22%) | 2 (7.69%) | 0 (0.00%) | | | |
| Likely (0.76-0.95) | 1 (25.00%) | 1 (20.00%) | 1 (25.00%) | 5 (13.51%) | 2 (28.57%) | 1 (25.00%) | 0 (0.00%) | 0 (0.00%) | 3 (75.00%) | 2 (15.38%) | 0 (0.00%) | 1 (25.00%) | 2 (28.57%) | 3 (33.33%) | 2 (7.69%) | 0 (0.00%) | | | |
| Plausible (0.56-0.75) | 3 (75.00%) | 4 (80.00%) | 1 (25.00%) | 30 (81.08%) | 5 (71.43%) | 3 (75.00%) | 2 (100.00%) | 0 (0.00%) | 1 (25.00%) | 11 (84.62%) | 16 (88.89%) | 3 (75.00%) | 5 (71.43%) | 4 (44.44%) | 22 (84.62%) | 4 (100.00%) | | | |

[1] n (%); Median (IQR)

B

| Characteristic | Max CTCAE Grade by Patient | | |
|---|---|---|---|
| | 1, N = 4[1] | 2, N = 1[1] | 3, N = 20[1] |
| Disease Categories | | | |
| CNS - Astrocytic-Oligodendroglial | 0 (0.00%) | 0 (0.00%) | 1 (5.00%) |
| CNS - Embryol | 2 (50.00%) | 0 (0.00%) | 18 (90.00%) |
| CNS - Ependymal | 1 (25.00%) | 1 (100.00%) | 0 (0.00%) |
| Solid Tumor - Neuroblastic tumors | 1 (25.00%) | 0 (0.00%) | 0 (0.00%) |
| Solid Tumor - Salivary Gland Tumors | 0 (0.00%) | 0 (0.00%) | 1 (5.00%) |

[1] n (%)

C

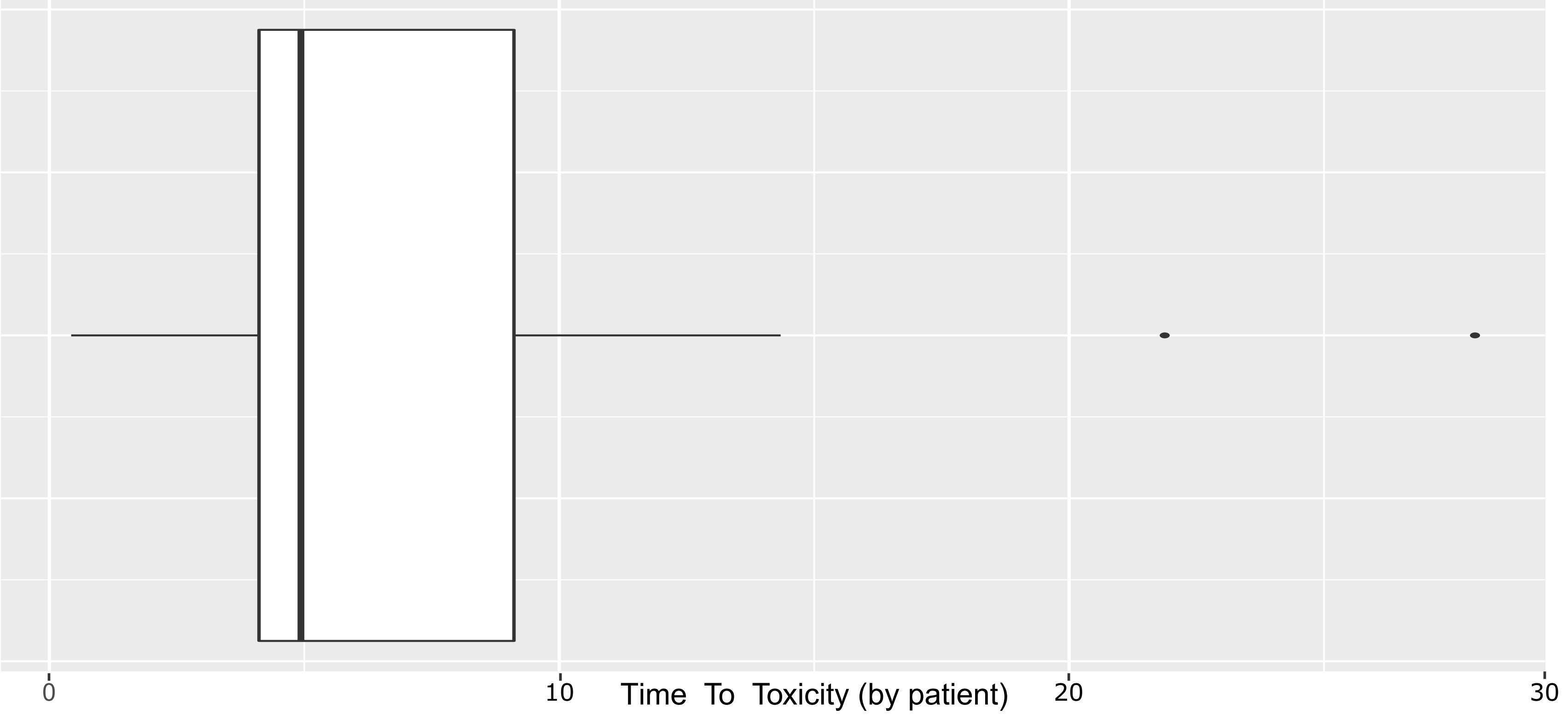


D

| Characteristic | Max CTCAE Grade by Patient | | |
|---|---|---|---|
| | 1, N = 4[1] | 2, N = 1[1] | 3, N = 20[1] |
| Medical Intervention | | | |
| Yes | 1 (25.00%) | 1 (100.00%) | 20 (100.00%) |
| No | 3 (75.00%) | 0 (0.00%) | 0 (0.00%) |
| Hearing Corrected | | | |
| Yes | 1 (100.00%) | 1 (100.00%) | 20 (100.00%) |
| No | 0 (0.00%) | 0 (0.00%) | 0 (0.00%) |

[1] n (%)

E

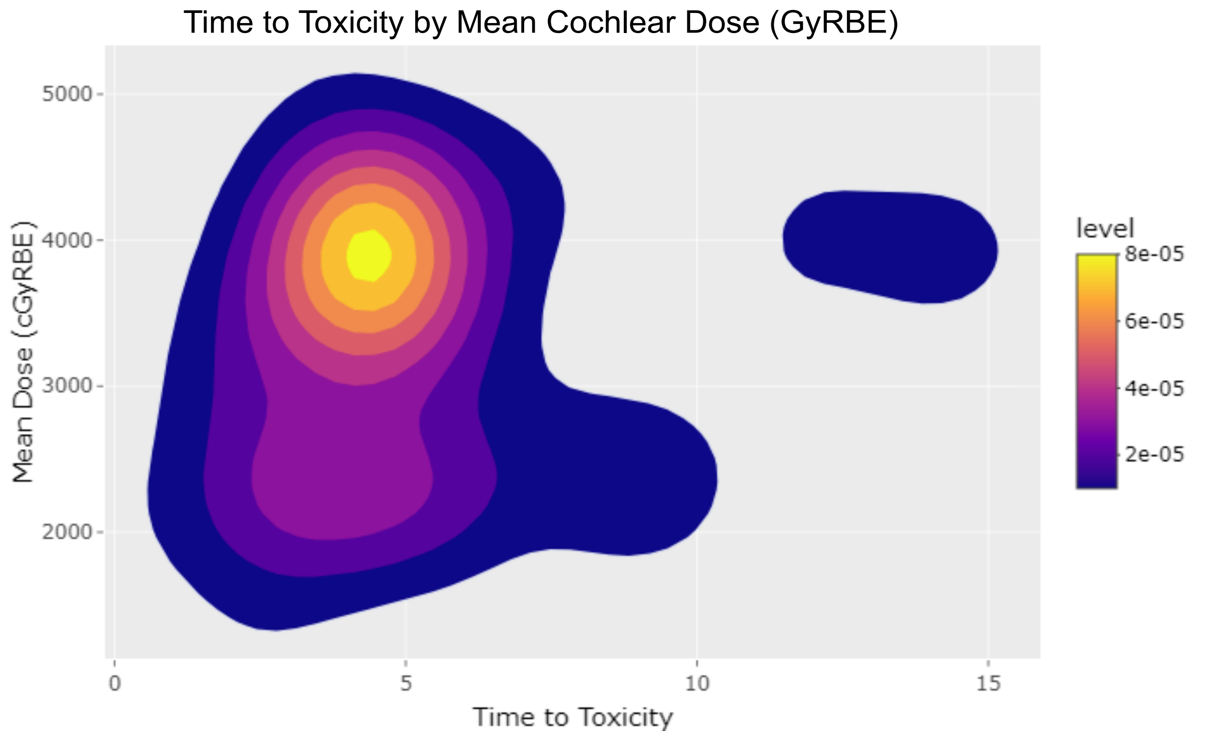

A

| Characteristic | Toxicity Classification: Fracture N = 8 | Toxicity Classification: Osteoradionecrosis N = 9 | CTCAE Grade: Grade 1 N = 13 | CTCAE Grade: Grade 2 N = 3 | CTCAE Grade: Grade 3 N = 1 |
|---|---|---|---|---|---|
| Prior Fracture | | | | | |
| Yes | 1 (12.50%) | 3 (33.33%) | 3 (23.08%) | 1 (33.33%) | 0 (0.00%) |
| No | 7 (87.50%) | 6 (66.67%) | 10 (76.92%) | 2 (66.67%) | 1 (100.00%) |
| Prior Steroids | | | | | |
| Yes | 5 (62.50%) | 3 (33.33%) | 6 (46.15%) | 2 (66.67%) | 0 (0.00%) |
| No | 3 (37.50%) | 6 (66.67%) | 7 (53.85%) | 1 (33.33%) | 1 (100.00%) |
| Region Affected | | | | | |
| Extremity | 1 (12.50%) | 5 (55.56%) | 6 (46.15%) | 0 (0.00%) | 0 (0.00%) |
| Mandible | 0 (0.00%) | 1 (11.11%) | 1 (7.69%) | 0 (0.00%) | 0 (0.00%) |
| Pelvis | 1 (12.50%) | 1 (11.11%) | 2 (15.38%) | 0 (0.00%) | 0 (0.00%) |
| Spine | 6 (75.00%) | 2 (22.22%) | 4 (30.77%) | 3 (100.00%) | 1 (100.00%) |
| Fracture Characteristic | | | | | |
| Avulsion Fracture | 1 (12.50%) | 0 (0.00%) | 0 (0.00%) | 1 (33.33%) | 0 (NA%) |
| Compression fracture | 6 (75.00%) | 2 (100.00%) | 6 (85.71%) | 2 (66.67%) | 0 (NA%) |
| Non-displaced acromial fracture | 1 (12.50%) | 0 (0.00%) | 1 (14.29%) | 0 (0.00%) | 0 (NA%) |

[1] n (%)

B

| Characteristic | Toxicity Classification: Fracture N = 8 | Toxicity Classification: Osteoradionecrosis N = 9 | CTCAE Grade: Grade 1 N = 13 | CTCAE Grade: Grade 2 N = 3 | CTCAE Grade: Grade 3 N = 1 |
|---|---|---|---|---|---|
| Chemotherapy Exposure | | | | | |
| Yes | 7 (87.50%) | 7 (77.78%) | 11 (84.62%) | 3 (100.00%) | 0 (0.00%) |
| No | 1 (12.50%) | 2 (22.22%) | 2 (15.38%) | 0 (0.00%) | 1 (100.00%) |
| Prior Surgery | | | | | |
| Yes | 4 (50.00%) | 5 (55.56%) | 6 (46.15%) | 2 (66.67%) | 1 (100.00%) |
| No | 4 (50.00%) | 4 (44.44%) | 7 (53.85%) | 1 (33.33%) | 0 (0.00%) |
| Patient Modality | | | | | |
| Mixed | 2 (25.00%) | 0 (0.00%) | 2 (15.38%) | 0 (0.00%) | 0 (0.00%) |
| Proton | 6 (75.00%) | 9 (100.00%) | 11 (84.62%) | 3 (100.00%) | 1 (100.00%) |
| Mean Dose (GyRBE) | 44.1 (22.5, 59.4) | 54.0 (54.0, 59.4) | 54.0 (34.2, 59.4) | 57.6 (40.8, 61.2) | 54.0 (54.0, 54.0) |
| Attribution | | | | | |
| Likely (0.76-0.95) | 4 (50.00%) | 6 (66.67%) | 7 (53.85%) | 2 (66.67%) | 1 (100.00%) |
| Plausible (0.56-0.75) | 4 (50.00%) | 3 (33.33%) | 6 (46.15%) | 1 (33.33%) | 0 (0.00%) |

[1] n (%); Median (IQR)

E

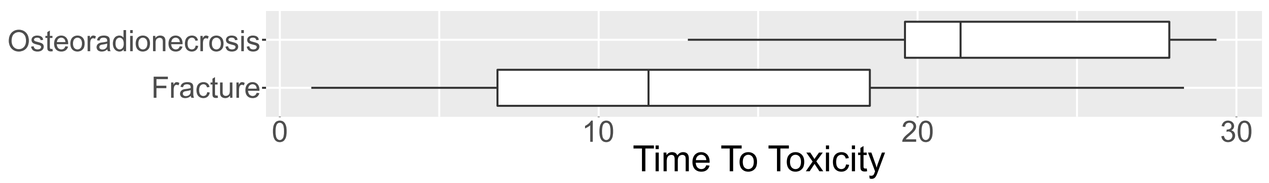


F

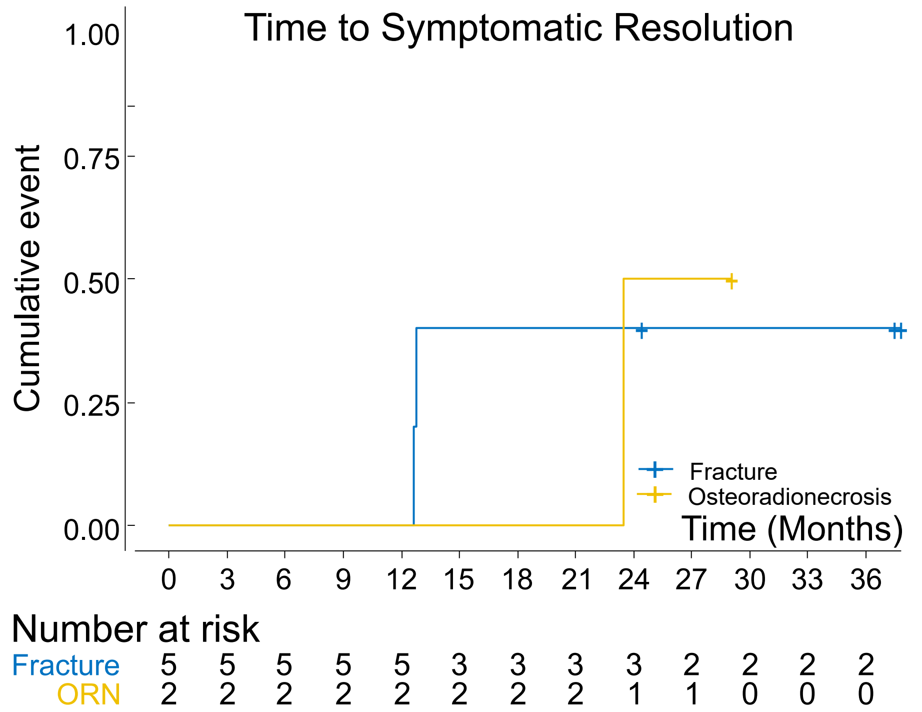


C

| Characteristic | Toxicity Classification: Fracture N = 8 | Toxicity Classification: Osteoradionecrosis N = 9 | CTCAE Grade: Grade 1 N = 13 | CTCAE Grade: Grade 2 N = 3 | CTCAE Grade: Grade 3 N = 1 | Symptoms Associated with Toxicity: Yes N = 7 | Symptoms Associated with Toxicity: No N = 10 |
|---|---|---|---|---|---|---|---|
| Surgical Stabilization | | | | | | | |
| Yes | 0 (0.00%) | 1 (11.11%) | 0 (0.00%) | 0 (0.00%) | 1 (100.00%) | 1 (14.29%) | 0 (0.00%) |
| No | 8 (100.00%) | 8 (88.89%) | 13 (100.00%) | 3 (100.00%) | 0 (0.00%) | 6 (85.71%) | 10 (100.00%) |
| Symptom Resolution | | | | | | | |
| Yes | 2 (40.00%) | 1 (50.00%) | 1 (33.33%) | 2 (66.67%) | 0 (0.00%) | 3 (42.86%) | 0 (NA%) |
| No | 3 (60.00%) | 1 (50.00%) | 2 (66.67%) | 1 (33.33%) | 1 (100.00%) | 4 (57.14%) | 0 (NA%) |
| Radiographic Resolution | | | | | | | |
| Yes | 3 (37.50%) | 2 (22.22%) | 3 (23.08%) | 2 (66.67%) | 0 (0.00%) | 3 (42.86%) | 2 (20.00%) |
| No | 5 (62.50%) | 7 (77.78%) | 10 (76.92%) | 1 (33.33%) | 1 (100.00%) | 4 (57.14%) | 8 (80.00%) |

[1] n (%)

D

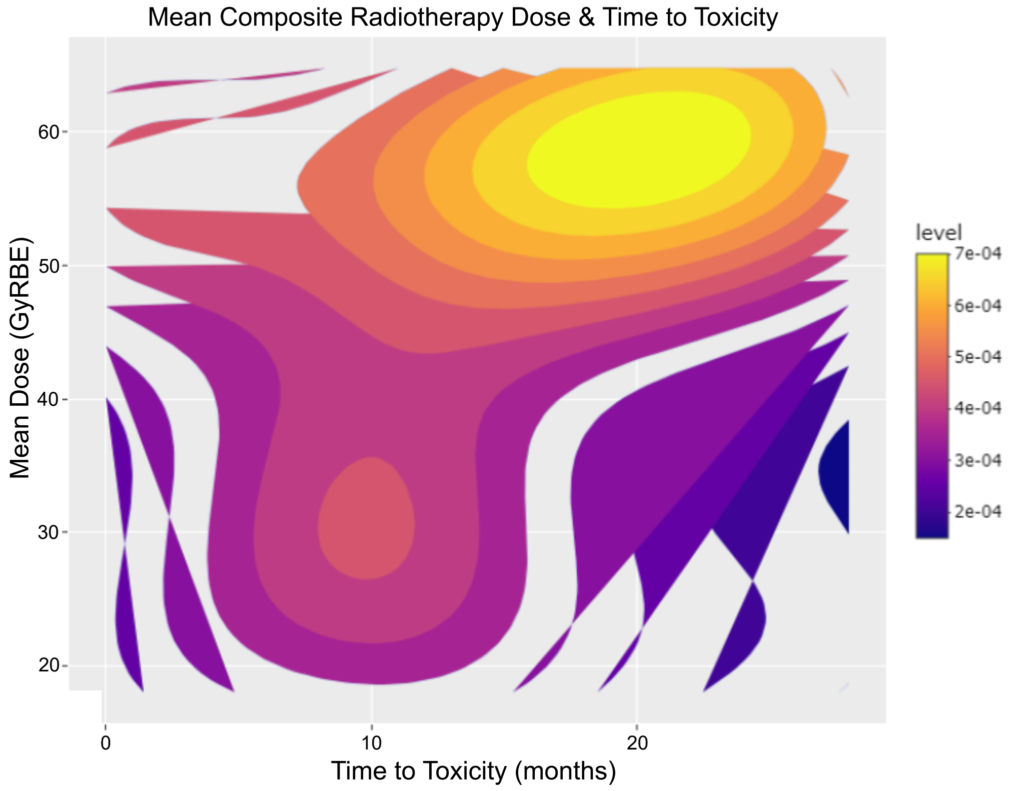


G

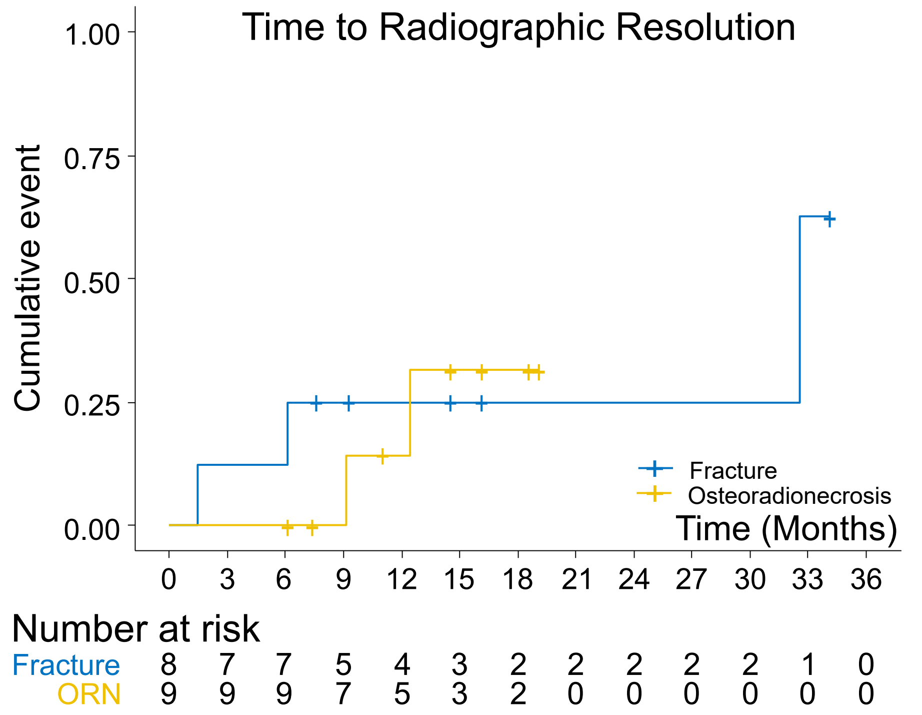

A

| Characteristic | Prior RT | Patient Modality | | Resolved | |
|---|---|---|---|---|---|
| | None, N = 27[1] | mixed, N = 7[1] | proton, N = 20[1] | Yes, N = 22[1] | No, N = 5[1] |
| Toxicity Type | | | | | |
| Anal mucositis | 1 (3.70%) | 0 (0.00%) | 1 (5.00%) | 1 (4.55%) | 0 (0.00%) |
| Anal pain | 2 (7.41%) | 0 (0.00%) | 2 (10.00%) | 2 (9.09%) | 0 (0.00%) |
| Duodenal ulcer | 1 (3.70%) | 0 (0.00%) | 1 (5.00%) | 1 (4.55%) | 0 (0.00%) |
| Dysphagia | 1 (3.70%) | 0 (0.00%) | 1 (5.00%) | 1 (4.55%) | 0 (0.00%) |
| Esophagitis | 2 (7.41%) | 0 (0.00%) | 2 (10.00%) | 2 (9.09%) | 0 (0.00%) |
| Ileus | 1 (3.70%) | 0 (0.00%) | 1 (5.00%) | 0 (0.00%) | 1 (20.00%) |
| Mucositis oral | 3 (11.11%) | 1 (14.29%) | 2 (10.00%) | 3 (13.64%) | 0 (0.00%) |
| Proctitis | 2 (7.41%) | 0 (0.00%) | 2 (10.00%) | 2 (9.09%) | 0 (0.00%) |
| Rectal pain | 1 (3.70%) | 0 (0.00%) | 1 (5.00%) | 1 (4.55%) | 0 (0.00%) |
| Small intestinal obstruction | 1 (3.70%) | 1 (14.29%) | 0 (0.00%) | 1 (4.55%) | 0 (0.00%) |
| Weight loss | 12 (44.44%) | 5 (71.43%) | 7 (35.00%) | 8 (36.36%) | 4 (80.00%) |

[1] n (%)

B

| Characteristic | CTCAE Grade |
|---|---|
| | G3, N = 27[1] |
| Disease Categories | |
| Astrocytic-Oligodendroglial | 1 (3.70%) |
| Bone Sarcoma - Uncertain Differentiation | 1 (3.70%) |
| Embryonal | 9 (33.33%) |
| Ependymal | 1 (3.70%) |
| Germ Cell | 1 (3.70%) |
| Neuroblastic tumors | 3 (11.11%) |
| Skeletal Muscle tumors | 11 (40.74%) |

[1] n (%)

C

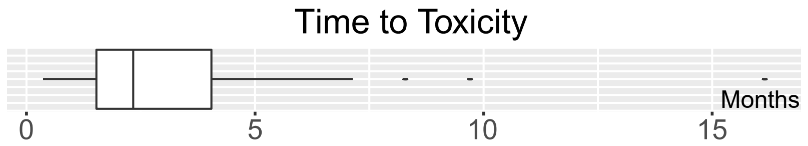


D

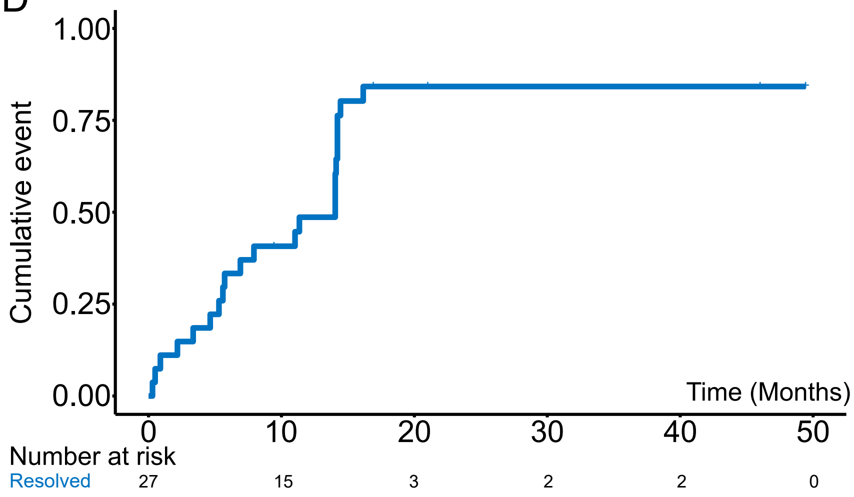

| Characteristic | Overall, N = 349[1] | Mixed, N = 74[1] | Proton, N = 275[1] |
|---|---|---|---|
| Age | 9.2 (5.5, 13.2) | 9.5 (6.4, 14.6) | 9.0 (5.1, 13.0) |
| Gender | | | |
| Female | 157 (44.99%) | 30 (40.54%) | 127 (46.18%) |
| Male | 192 (55.01%) | 44 (59.46%) | 148 (53.82%) |
| Insurance | | | |
| International Patient | 65 (18.62%) | 12 (16.22%) | 53 (19.27%) |
| Private | 78 (22.35%) | 15 (20.27%) | 63 (22.91%) |
| Public | 186 (53.30%) | 42 (56.76%) | 144 (52.36%) |
| Self Pay | 20 (5.73%) | 5 (6.76%) | 15 (5.45%) |
| BodySite | | | |
| CNS | 349 (100.00%) | 74 (100.00%) | 275 (100.00%) |
| Diagnoses | | | |
| ALL | 1 (0.29%) | 0 (0.00%) | 1 (0.36%) |
| Anaplastic Astrocytoma | 8 (2.29%) | 0 (0.00%) | 8 (2.91%) |
| ATRT | 15 (4.30%) | 3 (4.05%) | 12 (4.36%) |
| Brain, NOS | 1 (0.29%) | 1 (1.35%) | 0 (0.00%) |
| Brainstem Glioma | 1 (0.29%) | 1 (1.35%) | 0 (0.00%) |
| Chordoma | 1 (0.29%) | 0 (0.00%) | 1 (0.36%) |
| Choroid Plexus Carcinoma | 1 (0.29%) | 0 (0.00%) | 1 (0.36%) |
| CNS Embryonal Neoplasm | 3 (0.86%) | 0 (0.00%) | 3 (1.09%) |
| CNS Germ Cell Tumor | 11 (3.15%) | 1 (1.35%) | 10 (3.64%) |
| CNS Germinoma | 13 (3.72%) | 3 (4.05%) | 10 (3.64%) |
| CNS Glioneural Neoplasm | 2 (0.57%) | 0 (0.00%) | 2 (0.73%) |
| CNS PNET | 4 (1.15%) | 3 (4.05%) | 1 (0.36%) |
| Craniopharyngioma | 63 (18.05%) | 1 (1.35%) | 62 (22.55%) |
| Ependymoma | 71 (20.34%) | 10 (13.51%) | 61 (22.18%) |
| Ganglioglioma | 5 (1.43%) | 0 (0.00%) | 5 (1.82%) |
| Glioblastoma Multiforme | 8 (2.29%) | 2 (2.70%) | 6 (2.18%) |
| Glioneuronal Tumor | 1 (0.29%) | 1 (1.35%) | 0 (0.00%) |
| High Grade Astrocytoma | 4 (1.15%) | 1 (1.35%) | 3 (1.09%) |
| Lymphoma | 1 (0.29%) | 1 (1.35%) | 0 (0.00%) |
| Medulloblastoma | 103 (29.51%) | 44 (59.46%) | 59 (21.45%) |
| Meningioma | 2 (0.57%) | 0 (0.00%) | 2 (0.73%) |
| Myxopapillary Ependymoma | 6 (1.72%) | 1 (1.35%) | 5 (1.82%) |
| Non Germinomatous Germ Cell Tumor | 1 (0.29%) | 0 (0.00%) | 1 (0.36%) |
| Oligodendroglioma | 1 (0.29%) | 0 (0.00%) | 1 (0.36%) |
| Optic Pathway Glioma | 5 (1.43%) | 0 (0.00%) | 5 (1.82%) |
| Pilocytic Astrocytoma | 10 (2.87%) | 0 (0.00%) | 10 (3.64%) |
| Pineoblastoma | 1 (0.29%) | 0 (0.00%) | 1 (0.36%) |
| Retinoblastoma | 1 (0.29%) | 1 (1.35%) | 0 (0.00%) |
| Thoracic cord | 1 (0.29%) | 0 (0.00%) | 1 (0.36%) |
| WHO II Astrocytoma | 4 (1.15%) | 0 (0.00%) | 4 (1.45%) |
| Disease Categories | | | |
| Astrocytic-Oligodendroglial | 42 (12.03%) | 4 (5.41%) | 38 (13.82%) |
| Bone Sarcoma-Notochordal | 1 (0.29%) | 0 (0.00%) | 1 (0.36%) |
| Choroid Plexus | 1 (0.29%) | 0 (0.00%) | 1 (0.36%) |
| Embryonal | 126 (36.10%) | 50 (67.57%) | 76 (27.64%) |
| Ependymal | 77 (22.06%) | 11 (14.86%) | 66 (24.00%) |
| Germ Cell | 25 (7.16%) | 4 (5.41%) | 21 (7.64%) |
| Leukemia | 1 (0.29%) | 0 (0.00%) | 1 (0.36%) |
| Lymphoma | 1 (0.29%) | 1 (1.35%) | 0 (0.00%) |
| Meningioma | 2 (0.57%) | 0 (0.00%) | 2 (0.73%) |
| Neuronal-Mixed-glial | 7 (2.01%) | 1 (1.35%) | 6 (2.18%) |
| Other | 2 (0.57%) | 1 (1.35%) | 1 (0.36%) |
| Sellar | 63 (18.05%) | 1 (1.35%) | 62 (22.55%) |
| Soft Tissue Tumor | 1 (0.29%) | 1 (1.35%) | 0 (0.00%) |
| Disease Extent CNS | | | |
| Localized Disease | 241 (69.05%) | 26 (35.14%) | 215 (78.18%) |
| Metastatic | 67 (19.20%) | 38 (51.35%) | 29 (10.55%) |
| No evidence of disease | 41 (11.75%) | 10 (13.51%) | 31 (11.27%) |
| Extent of Resection CNS | | | |
| Gross total resection | 184 (52.72%) | 50 (67.57%) | 134 (48.73%) |
| Near-total resection | 41 (11.75%) | 6 (8.11%) | 35 (12.73%) |
| Sub-total resection | 71 (20.34%) | 13 (17.57%) | 58 (21.09%) |
| Unresected | 53 (15.19%) | 5 (6.76%) | 48 (17.45%) |
| Prior RT | | | |
| No Prior RT | 317 (90.83%) | 64 (86.49%) | 253 (92.00%) |
| Prior Photon | 15 (4.30%) | 5 (6.76%) | 10 (3.64%) |
| Prior Proton | 17 (4.87%) | 5 (6.76%) | 12 (4.36%) |

[1] Median (IQR); n (%)

| Characteristic | Overall, N = 151[1] | Mixed, N = 17[1] | Proton, N = 134[1] |
|---|---|---|---|
| Age | 12.1 (4.8, 16.4) | 11.6 (8.8, 15.6) | 12.2 (4.6, 16.4) |
| Gender | | | |
| Female | 71 (47.02%) | 7 (41.18%) | 64 (47.76%) |
| Male | 80 (52.98%) | 10 (58.82%) | 70 (52.24%) |
| Insurance | | | |
| International Patient | 12 (7.95%) | 0 (0.00%) | 12 (8.96%) |
| Private | 26 (17.22%) | 5 (29.41%) | 21 (15.67%) |
| Public | 102 (67.55%) | 11 (64.71%) | 91 (67.91%) |
| Self Pay | 11 (7.28%) | 1 (5.88%) | 10 (7.46%) |
| BodySite | | | |
| Abdomen | 28 (18.54%) | 6 (35.29%) | 22 (16.42%) |
| Chest | 22 (14.57%) | 1 (5.88%) | 21 (15.67%) |
| H&N | 51 (33.77%) | 6 (35.29%) | 45 (33.58%) |
| MSK | 22 (14.57%) | 3 (17.65%) | 19 (14.18%) |
| Pelvis | 28 (18.54%) | 1 (5.88%) | 27 (20.15%) |
| Diagnoses | | | |
| AML | 1 (0.66%) | 0 (0.00%) | 1 (0.75%) |
| Chondrosarcoma | 2 (1.32%) | 0 (0.00%) | 2 (1.49%) |
| Chordoma | 1 (0.66%) | 0 (0.00%) | 1 (0.75%) |
| Desmoplastic Small Round Cell Tumor (DSCRT) | 1 (0.66%) | 1 (5.88%) | 0 (0.00%) |
| Ewing's Sarcoma | 19 (12.58%) | 0 (0.00%) | 19 (14.18%) |
| Extraosseous Ewing's Sarcoma | 2 (1.32%) | 1 (5.88%) | 1 (0.75%) |
| Fibrosarcoma | 3 (1.99%) | 0 (0.00%) | 3 (2.24%) |
| Hodgkin's Disease | 24 (15.89%) | 4 (23.53%) | 20 (14.93%) |
| Maxillary sinus | 1 (0.66%) | 0 (0.00%) | 1 (0.75%) |
| MPNST | 1 (0.66%) | 0 (0.00%) | 1 (0.75%) |
| Mucoepidermoid Carcinoma | 2 (1.32%) | 0 (0.00%) | 2 (1.49%) |
| Myeloid Sarcoma | 1 (0.66%) | 0 (0.00%) | 1 (0.75%) |
| Nasopharyngeal Carcinoma | 3 (1.99%) | 1 (5.88%) | 2 (1.49%) |
| Neuroblastoma | 26 (17.22%) | 4 (23.53%) | 22 (16.42%) |
| Non-CNS Germ Cell Tumor | 2 (1.32%) | 0 (0.00%) | 2 (1.49%) |
| Osteosarcoma | 2 (1.32%) | 0 (0.00%) | 2 (1.49%) |
| Parotid gland | 1 (0.66%) | 0 (0.00%) | 1 (0.75%) |
| Rhabdoid Tumor | 1 (0.66%) | 0 (0.00%) | 1 (0.75%) |
| Rhabdomyosarcoma | 50 (33.11%) | 5 (29.41%) | 45 (33.58%) |
| Sarcoma (NOS) | 5 (3.31%) | 1 (5.88%) | 4 (2.99%) |
| Undifferentiated Carcinoma | 1 (0.66%) | 0 (0.00%) | 1 (0.75%) |
| Wilms' Tumor | 2 (1.32%) | 0 (0.00%) | 2 (1.49%) |
| Disease Categories | | | |
| Bone Sarcoma-Chondrogenic | 2 (1.32%) | 0 (0.00%) | 2 (1.49%) |
| Bone Sarcoma-Notochordal | 1 (0.66%) | 0 (0.00%) | 1 (0.75%) |
| Bone Sarcoma-Osteogenic | 2 (1.32%) | 0 (0.00%) | 2 (1.49%) |
| Bone Sarcoma-Uncertain Differentiation | 19 (12.58%) | 0 (0.00%) | 19 (14.18%) |
| Embryonal | 1 (0.66%) | 0 (0.00%) | 1 (0.75%) |
| Fibroblastic-Myofibroblastic | 3 (1.99%) | 0 (0.00%) | 3 (2.24%) |
| Germ Cell | 2 (1.32%) | 0 (0.00%) | 2 (1.49%) |
| Kidney Tumor | 2 (1.32%) | 0 (0.00%) | 2 (1.49%) |
| Leukemia | 2 (1.32%) | 0 (0.00%) | 2 (1.49%) |
| Lymphoma | 24 (15.89%) | 4 (23.53%) | 20 (14.93%) |
| Nerve sheath Tumor | 1 (0.66%) | 0 (0.00%) | 1 (0.75%) |
| Neuroblastic tumors | 26 (17.22%) | 4 (23.53%) | 22 (16.42%) |
| Salivary Gland Tumors | 3 (1.99%) | 0 (0.00%) | 3 (2.24%) |
| Sarcoma-Uncertain Differentiation | 1 (0.66%) | 1 (5.88%) | 0 (0.00%) |
| Sarcoma Other | 5 (3.31%) | 1 (5.88%) | 4 (2.99%) |
| Skeletal Muscle tumors | 50 (33.11%) | 5 (29.41%) | 45 (33.58%) |
| Soft Tissue Tumor | 4 (2.65%) | 1 (5.88%) | 3 (2.24%) |
| Uncertain Differentiation | 3 (1.99%) | 1 (5.88%) | 2 (1.49%) |
| Disease Extent Solid Tumor | | | |
| Diffuse Metastatic Disease | 85 (56.29%) | 12 (70.59%) | 73 (54.48%) |
| Local-Regional | 22 (14.57%) | 4 (23.53%) | 18 (13.43%) |
| Localized | 38 (25.17%) | 0 (0.00%) | 38 (28.36%) |
| Oligometastatic | 6 (3.97%) | 1 (5.88%) | 5 (3.73%) |
| Local Disease Burden | | | |
| No evidence of local disease | 11 (7.28%) | 1 (5.88%) | 10 (7.46%) |
| Residual <10% of dx volume | 15 (9.93%) | 1 (5.88%) | 14 (10.45%) |
| Residual >60% of dx volume | 24 (15.89%) | 0 (0.00%) | 24 (17.91%) |
| Residual 10-60% of dx volume | 50 (33.11%) | 8 (47.06%) | 42 (31.34%) |
| Residual microscopic disease | 16 (10.60%) | 2 (11.76%) | 14 (10.45%) |
| Untreated local disease | 35 (23.18%) | 5 (29.41%) | 30 (22.39%) |
| Prior RT | | | |
| No Prior RT | 136 (90.07%) | 15 (88.24%) | 121 (90.30%) |
| Prior Photon | 15 (9.93%) | 2 (11.76%) | 13 (9.70%) |

[1] Median (IQR); n (%)

### Predictors of Any ≥Grade 3 Toxicity: CNS Univariate Analysis

| Characteristics | HR | Lower CI | Upper CI | p-value |
|---|---|---|---|---|
| Age | | | | |
| group(0-3) | | | | |
| group(3-8) | 1.230 | 0.474 | 3.200 | 0.67 |
| group(8+) | 1.090 | 0.430 | 2.780 | 0.85 |
| Gender | | | | |
| Female | | | | |
| Male | 0.930 | 0.592 | 1.460 | 0.75 |
| Insurance | | | | |
| Private | | | | |
| Public | 0.995 | 0.567 | 1.750 | 0.99 |
| SelfPay | 0.995 | 0.521 | 1.900 | 0.99 |
| Patient modality | | | | |
| Mixed | | | | |
| Proton | 0.295 | 0.187 | 0.466 | 1.6e-07 |
| Enrollment Category | | | | |
| Prospective | | | | |
| Retrospective | 1.250 | 0.781 | 1.990 | 0.35 |
| Disease extent | | | | |
| Localized Disease | | | | |
| Metastatic | 3.340 | 2.030 | 5.500 | 2.1e-06 |
| No evidence of disease | 1.800 | 0.940 | 3.440 | 0.076 |
| Prior RT | | | | |
| No Prior RT | | | | |
| Prior Photon/Proton | 2.050 | 1.090 | 3.890 | 0.027 |
| Extent of resection | | | | |
| Gross total resection | | | | |
| Near-total resection | 0.754 | 0.352 | 1.610 | 0.47 |
| Sub-total resection | 0.562 | 0 292 | 1.080 | 0.085 |
| Unresected | 0.641 | 0.318 | 1.290 | 0.21 |
| Systemic Disease Status | | | | |
| No evidence of disease | | | | |
| Progressive | 2.170 | 1.110 | 4.240 | 0.023 |
| Stable | 1.350 | 0.780 | 2.340 | 0.28 |
| Systemic Dz Treatment Status | | | | |
| No systemic disease | | | | |
| Treated, Prior chemotherapy | 1.190 | 0.691 | 2.040 | 0.54 |
| Local disease status | | | | |
| No evidence of disease | | | | |
| Progressive | 1.220 | 0.711 | 2.110 | 0.46 |

### Predictors of Any ≥Grade 3 Toxicity: CNS Multivariate Model

| Characteristics | HR | Lower CI | Upper CI | p-value |
|---|---|---|---|---|
| Patient modality | | | | |
| Proton | | | | |
| Mixed | 2.618 | 1.509 | 4.54 | 0.00062 |
| Prior RT | | | | |
| No Prior RT | | | | |
| Prior Photon/Proton | 1.800 | 0.876 | 3.70 | 0.11000 |
| Disease extent | | | | |
| Localized Disease | | | | |
| Metastatic | 1.672 | 0.945 | 2.96 | 0.07800 |
| Age | | | | |
| group(0-3) | | | | |
| group(3+) | 0.765 | 0.293 | 2.00 | 0.59000 |
| Baseline total toxicity burden | 1.043 | 1.012 | 1.07 | 0.00690 |

### Predictors of Any ≥Grade 3 Toxicity: Non-CNS Univariate Analysis

| Characteristics | HR | Lower CI | Upper CI | p-value |
|---|---|---|---|---|
| Age | | | | |
| group(0-3) | | | | |
| group(3-8) | 1.380 | 0.348 | 5.49 | 0.650 |
| group(8+) | 1.070 | 0.307 | 3.73 | 0.910 |
| Gender | | | | |
| Female | | | | |
| Male | 1.250 | 0.561 | 2.78 | 0.590 |
| Insurance | | | | |
| Private | | | | |
| Public | 1.050 | 0.357 | 3.11 | 0.920 |
| SelfPay | 1.160 | 0.296 | 4.57 | 0.830 |
| Enrollment Category | | | | |
| Prospective | | | | |
| Retrospective | 0.792 | 0.347 | 1.81 | 0.580 |
| Disease extent | | | | |
| Diffuse Metastatic Disease | | | | |
| Localized | 1.290 | 0.583 | 2.85 | 0.530 |
| Extent of resection | | | | |
| Residual 10-60% of dx volume | | | | |
| Residual microscopic disease | 0.914 | 0.312 | 2.67 | 0.870 |
| Systemic Dz Treatment Status | | | | |
| No systemic disease | | | | |
| Treated, Prior chemotherapy | 0.717 | 0.325 | 1.58 | 0.410 |
| Local disease treatment status | | | | |
| Treated, Prior chemo & surgery | | | | |
| Untreated local disease | 1.640 | 0.719 | 3.73 | 0.240 |
| Bodysite | | | | |
| Abdomen | | | | |
| Head & Neck / Musculoskeletal | 2.330 | 0.770 | 7.05 | 0.130 |
| Pelvis | 3.420 | 1.020 | 11.50 | 0.047 |

### Predictors of Any ≥Grade 3 Toxicity: Non-CNS Multivariate Model

| Characteristics | HR | Lower CI | Upper CI | p-value |
|---|---|---|---|---|
| Bodysite | | | | |
| Abdomen-Chest | | | | |
| Head & Neck / Musculoskeletal | 2.93 | 0.82 | 10.49 | 0.0980 |
| Pelvis | 4.25 | 1.08 | 16.72 | 0.0380 |
| Baseline total toxicity burden | 1.10 | 1.04 | 1.16 | 0.0015 |
