## Supplemental Methods for "A Phase IV Trial of Proton Therapy in Children: The First Report from the SJPROTON1 Study"

*Variables Examined*

Formatted: Numbering: Continuous

Patient, disease, and treatment variables of interest were prespecified in the protocol and documented at baseline and in follow-up according to mCODE standards[1] in MOSAIQ Radiation Oncology (2021 Elekta). Toxicity characteristics were documented at baseline (pre-radiotherapy) and post-radiotherapy toxicity was graded as per CTCAE v4.0 at the end of proton therapy, either 4-6 weeks or 6 months. Starting at one year following proton therapy, toxicities occurring at annual evaluations (years 1-5, 10) were graded according to the SJLIFE-modified CTCAE.[2] Toxicities from each scale were categorized as shown in (SD1).

Quarterly quality assurance review ensured data integrity (SF2). Participant clinical trial status was regularly reviewed and categorized as on trial-on therapy, on trial in follow-up, progressed-on trial, off trial-records retained. Participants experiencing a cancer related event were followed for the cause of death, but no further protocol mandated follow-up visits were required, and no further toxicities were documented. Participants who died or were no longer in follow-up due to no potential observable toxicity events were labelled as off trial-records retained (Table 1).

The following toxicities were captured, regardless of grade: CNS necrosis, vasculopathy, endocrinopathy, permanent neurologic deficit, osteoradionecrosis, and fracture. Neurologic deficits after PBS-PT were categorized as “anatomic” when a deterministic relation between radiation dose and the deficit was biologically plausible (e.g., hearing or vision loss) or “nonanatomic” when the relationship between anatomic dose distribution and biological cause of the deficit was not apparent (e.g., seizure, ataxia, cognitive changes, hypersomnia).

Toxicity events were reconstructed relative to the PBS-RT timeline and other clinical events. The raw data used to generate each swimmer plot is shown in Supplementary Data – Swimmer Plot Data (SD2). To ensure that prespecified toxicity events were not omitted, provider entered data were augmented with an EMR search (nDepth, 2020 Regenstrief Institute, Inc.) using prespecified search terms for each prespecified toxicity event and underwent additional review to better understand the circumstances contributing to the event and facilitate attribution (SF4). This required the collection of additional detailed information regarding known risk factors for disease and treatment related morbidity. The details of these risk factors are in the corresponding supplementary figures and supplementary data for each event type.[3-7] A prespecified process for toxicity attribution was utilized to classify each potentially radiotherapy attributable event as either [excluded (0-0.05), unlikely (0.06-0.25), doubtful (0.26-0.45), unassessable (0.46-0.55), plausible (0.56-0.75), likely (0.76-0.95), or certain (0.96-1.0)]. Further details on this process are documented in SF5. The total toxicity burden at each timepoint for each patient was calculated by taking the sum of the product of each event and grade. Radiotherapy toxicity attributable hospitalization and procedure events were documented with the date of admission, duration, and reason. Further details on the type and indication of the hospitalization and procedures are illustrated in Supplementary Data – Hospitalization-Procedures (SD3).

Patient and disease characteristics were recorded at protocol-scheduled clinical evaluations and augmented with electronic medical record (Cerner PowerChart, 2020) review. Patient characteristics included age at diagnosis and enrollment, birth date, zip code, race, insurance type, and ethnicity. Insurance category was used as a surrogate for access to care and patients were categorized as private, international, public, or self-pay. Disease characteristics were categorized according to tumor origin (central nervous system (CNS), solid tumor, and leukemia/lymphoma), body location (CNS, chest, head and neck, abdomen, pelvis, musculoskeletal), disease category (ICD-O), extent/stage (localized or metastatic), cancer disease status (refractory, progressive or stable), local disease status (NED, stable, progressive, relapse). Disease categories were defined according to World Health Organization groupings (SD4)[8, 9] and were utilized to avoid the unmasking of disease specific trials conducted at St. Jude Children's Research Hospital.[10] Treatment characteristics were categorized according to extent of resection, prior treatment (treated, undergoing treatment or untreated), prior radiotherapy (prior photon, prior proton, no prior radiotherapy) and concurrent trial enrollment status (NPTP, disease specific protocol).

*Patient Outcomes*

The time to first ≥ grade 3 toxicity was the study-specified primary endpoint. Patients who experienced treatment failure were followed for cause of death thereafter. Failure events were categorized as local, regional, and/or distant, according to established disease- and protocol-specific standards. Local failure events were categorized as either central/infield (i.e., new tumor detected within the 80% isodose line) or marginal (i.e., new tumor detected outside the 80% isodose line) after co-registration and review of each radiotherapy plan and image revealing treatment failure.[11] Treatment-related mortality was defined according to Alexander *et al.*[1]

Works Cited


1. Alexander S, Pole JD, Gibson P, et al. Classification of treatment-related mortality in children with cancer: a systematic assessment. The Lancet Oncology. Dec 2015;16(16):e604-10. doi:10.1016/S1470-2045(15)00197-7
2. Schipper MJ, Taylor JM, Smith GL, Jagsi R. Comparing long-term treatment-associated toxicities in cancer patients: approaches, caveats, and recommendations. Int J Radiat Oncol Biol Phys. Jun 1 2014;89(2):232-4. doi:10.1016/j.ijrobp.2014.01.030
3. Gajjar A. Risk-Adapted Therapy for Young Children With Embryonal Brain Tumors, Choroid Plexus Carcinoma, High Grade Glioma or Ependymoma: ClinicalTrials.gov Identifier: NCT00602667. https://clinicaltrials.gov/ct2/show/NCT00602667?term=00602667&draw=2&rank=1. 2008.
4. Brown AP, Barney CL, Grosshans DR, et al. Proton beam craniospinal irradiation reduces acute toxicity for adults with medulloblastoma. Int J Radiat Oncol Biol Phys. Jun 1 2013;86(2):277-84. doi:10.1016/j.ijrobp.2013.01.014
5. Indelicato DJ, Flampouri S, Rotondo RL, et al. Incidence and dosimetric parameters of pediatric brainstem toxicity following proton therapy. Acta oncologica. Oct 2014;53(10):1298-304. doi:10.3109/0284186X.2014.957414
6. Devine CA, Liu KX, Ioakeim-Ioannidou M, et al. Brainstem Injury in Pediatric Patients Receiving Posterior Fossa Photon Radiation. Int J Radiat Oncol Biol Phys. Dec 1 2019;105(5):1034-1042. doi:10.1016/j.ijrobp.2019.08.039
7. Haas-Kogan D, Indelicato D, Paganetti H, et al. National Cancer Institute Workshop on Proton Therapy for Children: Considerations Regarding Brainstem Injury. Int J Radiat Oncol Biol Phys. May 1 2018;101(1):152-168. doi:10.1016/j.ijrobp.2018.01.013
8. Giantsoudi D, Adams J, MacDonald SM, Paganetti H. Proton Treatment Techniques for Posterior Fossa Tumors: Consequences for Linear Energy Transfer and Dose-Volume Parameters for the Brainstem and Organs at Risk. Int J Radiat Oncol Biol Phys. Feb 1 2017;97(2):401-410. doi:10.1016/j.ijrobp.2016.09.042
9. Giantsoudi D, Sethi RV, Yeap BY, et al. Incidence of CNS Injury for a Cohort of 111 Patients Treated With Proton Therapy for Medulloblastoma: LET and RBE Associations for Areas of Injury. Int J Radiat Oncol Biol Phys. May 1 2016;95(1):287-296. doi:10.1016/j.ijrobp.2015.09.015
10. Yock TI, Tarbell NJ. Technology insight: Proton beam radiotherapy for treatment in pediatric brain tumors. Nat Clin Pract Oncol. Dec 2004;1(2):97-103; quiz 1 p following 111. doi:10.1038/ncponc0090
11. McDonald MW, Shu HK, Curran WJ, Jr., Crocker IR. Pattern of failure after limited margin radiotherapy and temozolomide for glioblastoma. Int J Radiat Oncol Biol Phys. Jan 1 2011;79(1):130-6. doi:10.1016/j.ijrobp.2009.10.048
