## Supplementary material for "A Phase IV Trial of Proton Therapy in Children: The First Report from the SJPROTON1 Study": SJPROTON1 Protocol

SJCRH
SJPROTON1
Initial version, dated: 00-00-00, (IRB Approved: 00-00-00)

### Phase IV Clinical Trial of Proton Therapy in Pediatric Cancer

**Principal Investigator**
**John T. Lucas, Jr., MS, MD[1]**

**Co-Investigators**
Thomas E. Merchant, DO, PhD[1]
Matthew J. Krasin, MD[1]
Christopher L. Tinkle, MD, PhD[1]
Jonathan B. Farr, PhD[1]
Chia-Ho Hua, PhD[1]
Weiguang Yao, PhD[1]
Noah D. Sabin, MD, JD[2]
Sue Kaste, MD[2]
Kathleen J. Helton, MD[2]
Gregory T. Armstrong, MD, MSCE[3]
Gang Wu, PhD[5]
Suzanne J. Baker, PhD[6]
Mary V. Relling, PhD[7]
Brent A. Orr, PhD[8]
Yimei Li, PhD[4]
Shengjie Wu, MS[4]

St. Jude Children's Research Hospital
262 Danny Thomas Place
Memphis, Tennessee 38105-3678

[1]Department of Radiation Oncology
[2]Department of Diagnostic Imaging
[3]Department of Epidemiology and Cancer Control
[4]Department of Biostatistics
[5]Department of Computational Biology
[6]Department of Developmental Neurobiology
[7]Department of Pharmaceutical Sciences
[8]Department of Pathology

Contents of this document may not be extracted without permission from the Principal Investigator.

**Protocol Summary**

| |
|---|
| **SJPROTON1 - Phase IV Clinical Trial of Proton Therapy in Pediatric Cancer** |
| **Principal Investigator:** John T. Lucas Jr., MS, MD |
| **IND Holder:** N/A |
| **Brief Overview:** This is a Phase IV clinical trial evaluating the safety of proton therapy in children. The radiation targeting, planning, prescribed dose, fractionation, schedule, and use of other forms of therapy will be at the discretion of the treating physicians. Prior, concurrent, and/or sequential enrollment on disease- or site-specific therapeutic protocols is permitted. When feasible, a baseline assessment of disease and patient characteristics will be performed before proton therapy; this will be followed by serial standard-of-care clinical assessments to evaluate acute and late complications, disease control, treatment-related mortality, and overall survival. |
| **Intervention:** Proton therapy targeting, planning, prescribed dose, fractionation, and sequencing will be delivered according to the tumor type, extent of disease, and factors coincident with the patient's overall treatment plan. |
| **Brief Outline of Treatment Plan:** Proton therapy prescription parameters will be chosen according to the tumor type and disease status. Serial assessments will document toxicity according to the current version of the CTCAE, as well as organ-specific complications outlined in secondary objectives. In the event that proton therapy results in an increased incidence of complications within a patient cohort, compared to the expected incidence when using photon-based radiation therapy, an extensive review will be conducted to determine the attribution to proton therapy.<br><br><u>Primary Objective:</u><br><br>1.1.1 To estimate the incidence of radiation associated grade 3 and grade 4 non-hematologic toxicities at 1, 3, 5, and 10 years in a radiated region specific manner after the initiation of proton therapy.<br>**Responsible Investigator**: John T. Lucas, MS, MD<br>**Biostatisticians:** Yimei Li, PhD, Shengje Wu, MS<br>**Estimated date for completion of data collection**: June 2037.<br><br><u>Secondary Objectives</u><br><br>1.2.1 To estimate the incidence of necrosis, vasculopathy, and symptomatic and permanent neurologic deficits at 1, 3, 5, and 10 years after the initiation of proton therapy in children treated for CNS tumors in an irradiated region directed manner.<br><br>**Responsible Investigators**: John T. Lucas Jr, MD, Christopher L. Tinkle, MD, PhD, Thomas E. Merchant, DO, PhD, Noah Sabin, MD, and Kathleen Helton, MD<br>**Biostatisticians:** Yimei Li, PhD<br>**Estimated date for completion of data collection:** Three-year outcomes will be reported once a cohort of 100 to 200 patients has reached a median follow-up of 3 years. |

**SJPROTON1 - Phase IV Clinical Trial of Proton Therapy in Pediatric Cancer**

1.2.2 To estimate the incidence of treatment-related mortality at 5 and 10 years after the initiation of proton therapy.

**Responsible Investigator**: John T. Lucas, MS, MD, Gregory T. Armstrong, MD, MSCE
**Biostatisticians:** Yimei Li, PhD, Shengjie Wu, MS
**Estimated date for completion of data collection**: June 2037.

1.2.3 To estimate the incidence of secondary malignancies at 5 and 10 years after the initiation of proton therapy in an irradiated region directed manner.

**Responsible Investigator**: John T. Lucas, MS, MD, Gregory T. Armstrong, MD, MSCE
**Biostatisticians:** Yimei Li, PhD, Shengjie Wu, MS
**Estimated date for completion of data collection**: June 2037.

1.2.4 To estimate the incidence of fracture, and osteonecrosis at 1, 3, 5, and 10 years after the initiation of proton therapy in children treated for musculoskeletal tumors in an irradiated region directed manner.

**Responsible Investigators**: Sue Kaste, DO, Matthew Krasin, MD, and John Lucas, MD
**Biostatisticians:** Yimei Li, PhD
**Estimated date for completion of data collection:** Three-year outcomes will be reported once a cohort of 100 to 200 patients has reached a median follow-up of 3 years.

<u>Exploratory Objectives</u>

1.3.1 To evaluate the role of treatment delivery and planning parameters in local control and toxicity in an irradiated region directed manner.

**Responsible Investigator**: John T. Lucas, MS, MD
**Biostatisticians:** Yimei Li, PhD, Shengjie Wu, MS
**Estimated date for completion of data collection**: June 2037

1.3.2 To evaluate the mutational signature in patients with radiation associated high grade gliomas.

**Responsible Investigator**: John T. Lucas, MS, MD, Suzanne J. Baker, PhD, Brent A. Orr, PhD
**Bioinformatician:** Gang Wu, PhD
**Estimated date for completion of data collection**: Spring 2019

1.3.3 To evaluate the contribution of systemic therapies in perpetuating radiation associated toxicities.

**SJPROTON1 - Phase IV Clinical Trial of Proton Therapy in Pediatric Cancer**

**Responsible Investigator**: John T. Lucas, MS, MD, Mary V. Relling, PhD
**Biostatisticians:** Yimei Li, PhD, Shengjie Wu, MS
**Estimated date for completion of data collection**: Reporting of the impact of systemic treatment parameters on radiation associated toxicities will be limited to frequent combinations of systemic therapies in higher-volume disease sites with larger expected accrual rates to ensure appropriate statistical power.

1.3.4 To compare specific proton and photon radiotherapy associated toxicities specified in 1.2.1 and 1.2.4.

**Responsible Investigator**: John T. Lucas, MS, MD, Christopher L. Tinkle, MD, PhD, Thomas E. Merchant, DO, PhD, Matthew Krasin, MD
**Biostatisticians:** Yimei Li, PhD, Shengjie Wu, MS
**Estimated date for completion of data collection**: Reporting of the impact of treatment delivery parameters on local control and toxicity will be limited to higher-volume disease sites with larger expected accrual rates to ensure appropriate statistical power.

**Study Design:** Phase IV clinical trial

**Sample Size:** An enrollment threshold is not specified. Approximately 1000 patients are projected to be enrolled over 10 years. The time interval for study enrollment is 10 years, with 10 years of follow-up for each enrolled participant.

**Randomization:** N/A

**Inclusion Criteria:**

1. The patient was or will be treated with proton therapy at St. Jude Children's Research Hospital on or after November 18, 2015.

**Anticipated Primary Completion Date:** June 2037

**Anticipated Study Completion Date:** June 2037

**Timeframe for Primary Outcome:** June 2037

**Data Management:** Data management and statistical analysis will be provided locally by the Department of Radiation Oncology and the Department of Biostatistics at St. Jude Children's Research Hospital.

**Human Subjects:** The risks associated with enrollment on this clinical trial are minimal. The risks associated with proton therapy depend on host, clinical, and treatment factors and will be considered individually. Common side effects during proton therapy include nausea, vomiting, fatigue, loss of appetite, hair loss, and headache. Cyst expansion during treatment may result in transient or permanent neurologic symptoms or deficits. Common

| SJPROTON1 - Phase IV Clinical Trial of Proton Therapy in Pediatric Cancer |
| --- |
| side effects after treatment include hormone deficiencies and weight gain, fatigue and sleep disorders, cognitive effects (affecting memory, attention, behavior, learning, or global intelligence), and headache. Less common are secondary tumor formation (benign or malignant); vasculopathy and stroke; neurologic effects such as hearing loss, vision loss, seizures, basal ganglia syndrome, and dystonia; and structural effects such as necrosis or myelopathy that may lead to permanent disability or death. Patients will be informed of these and other possible side effects during the informed consent process. Adverse events will be reported, and treated appropriately. Patients will sign informed consent to participate in this research. |

### 1.0 OBJECTIVES

#### 1.1 Primary Objective

1.1.1 To estimate the incidence of radiation associated grade 3 and grade 4 non-hematologic toxicities at 1, 3, 5, and 10 years in a radiated region specific manner after the initiation of proton therapy.

Statement: Variations in the transfer of energy to tissue in the path of proton beams may be associated with complications of proton therapy. Efforts to systematically assess the impact of differential particle energy deposition at depth have been limited. It is unknown if the incidence of complications and rates of tumor control for proton therapy are comparable to those for photon therapy.

#### 1.2 Secondary Objectives

1.2.1 To estimate the incidence of necrosis, vasculopathy, and symptomatic and permanent neurologic deficits at 1, 3, 5, and 10 years after the initiation of proton therapy in children treated for CNS tumors in an irradiated region directed manner.

1.2.2 To estimate the incidence of treatment-related mortality at 5 and 10 years after the initiation of proton therapy.

1.2.3 To estimate the incidence of secondary malignancies at 5 and 10 years after the initiation of proton therapy in an irradiated region directed manner.

1.2.4 To estimate the incidence of fracture, and osteonecrosis at 1, 3, 5, and 10 years after the initiation of proton therapy in children treated for musculoskeletal tumors in an irradiated region directed manner.

#### 1.3 Exploratory Objectives

1.3.1 To evaluate the role of treatment delivery and planning parameters in local control and toxicity in an irradiated region directed manner.

1.3.2 To evaluate the mutational signature in patients with radiation associated high grade gliomas.

1.3.3 To evaluate the contribution of systemic therapies in perpetuating radiation associated toxicities.

1.3.4 To compare specific proton and photon radiotherapy associated toxicities specified in 1.2.1, and 1.2.4

#### 1.4 Definitions

1.4.1 *Clinical factors* include but are not limited to vital statistics, auxological measures, and tumor variables that uniquely identify the patient and the natural history of their disease. Clinical factors also include laboratory measures of host and tumor biology and response to treatment.

1.4.2 *Treatment factors* include but are not limited to the type and extent of surgical interventions, including operative complications. Treatment factors also include the full range of proton therapy treatment planning and delivery parameters.

1.4.3 *Measures of radiation effects* refers to the broad categories of different objective clinical or laboratory methods used to evaluate the acute and late effects of proton therapy in this protocol (including evaluations of neuropsychologic effects; growth factors and cytokines; endocrine aspects; physical performance and movement; host and tumor genomics; structural, functional, and vascular imaging; audiology, neurology, and ophthalmology; sleep and fatigue; and quality of life). Each of these categories includes smaller or more specific measures. Some of these measures may be used to evaluate tumor response to treatment (blood tests, tumor and host genomics, and imaging).

### 2.0 BACKGROUND AND RATIONALE

#### 2.1 Background

Radiation therapy with protons may have significant advantages over radiation therapy with photons for children with brain tumors, solid tumors, lymphoma, or other conditions. The primary advantage of proton therapy appears to be the ability to reduce the radiation dose to normal tissue, which is derived from the ability to control the depth of penetration of protons in tissue (Figure 1). With the advent of discrete proton spot scanning, another advantage of proton therapy is the ability to precisely control the position of the beam to increase the conformity of the prescription dose to the targeted volume and increase the radiation dose to selected areas of the targeted volume.

The background for this protocol is organized according to the following topics:

- Problem definition and unknowns in the use of proton therapy in children
- The promise of proton therapy in children
- Physical dose differences between protons and photons
- Biological dose differences between protons and photons
- The use of proton therapy in children
- The role of changing technology in proton therapy delivery

- Proton therapy and local tumor control
- Proton therapy and treatment-related morbidity and mortality
- The role of clinical trials in monitoring treatment-related adverse events: Why a phase IV trial and not a registry?

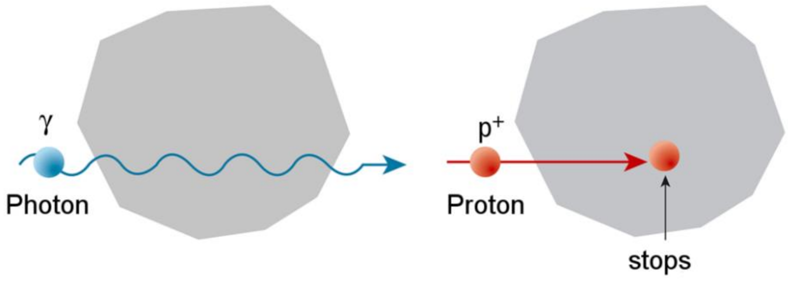

*Figure 1. Protons and their properties | Paul Scherrer Institut (PSI)*

The Problem

Despite the potential advantages of proton therapy and more than 50 years of experience with the modality, there remain unanswered questions about its safety and efficacy in children [1]. The physical and biological characteristics of proton beams have not been adequately studied. The US Food and Drug Administration considers proton therapy and its various delivery methods to be substantially equivalent to photon therapy and its delivery methods. The commercial availability of clinical proton therapy systems and the recognition by the Centers for Medicare and Medicaid Services of their financial responsibility to support the use of this treatment in appropriate cases serves as recognition of proton therapy as a mainstream form of treatment for pediatric and adult cancers and special conditions.

The Promise of Proton Therapy in Children

The promise of proton therapy is largely associated with the reduction in the volume of normal tissue exposed to collateral irradiation [2]. Over the past 20 years, St. Jude investigators have designed institutional clinical trials for treating a variety of tumor types in which they have demonstrated that the targeted volume may be safely reduced without affecting the rate or pattern of failure. Noteworthy examples include reducing the boost volume for medulloblastoma from the cooperative group standard of the anatomic posterior fossa (the pre-1996 standard) to the postoperative tumor bed plus a 2-cm margin in the SJMB96 trial [3], to the postoperative tumor bed plus a 1-cm margin in the SJMB03 trial [4], and to the postoperative tumor bed plus a 0.5-cm margin in the SJMB12 trial [5]; the treatment of ependymoma from the cooperative group standard of the preoperative tumor plus a 1.5-cm to 2-cm margin to the postoperative tumor bed plus a 1-cm margin in the RT1 trial [6] and to the postoperative tumor bed plus a 0.5-cm margin in the SJYC07 trial [7]; the treatment of rhabdomyosarcoma and other soft-tissue and bone tumors of childhood from the cooperative group standard of the anatomic compartment or pre-therapy tumor plus a 2-cm margin to the pre-/postoperative tumor bed plus a 1.5-cm to 1-cm margin in the RTSARC trial [8] and to the current postoperative tumor bed plus a 0.5-cm margin in the RMS13 [9] and ESFT13 [10] trials. In pursuing target volume reduction for these and other tumor types, models of radiation dose-volume effects have been developed for a large number of organ systems (**Table 1**) that demonstrate the impact of the volume receiving both high and low doses of radiation. Intensity-modulated proton therapy delivered using the discrete spot-scanning method that is currently available at St. Jude has the potential to reduce the volume that receives the highest dose through superior prescription dose conformity and, in some cases, to avoid normal tissue structures altogether. At the very least, proton therapy reduces the volume of normal tissues exposed to the lowest doses when compared to photon therapy; this advantage is based on the unique physical characteristics of protons that allow them to stop within the targeted volume [11].

*Table 1: Sample models of radiation dose and volume effects developed by St. Jude investigators.*

| **Organ (diagnosis)** | **Effect** | **Reference** |
|---|---|---|
| Bone (bone and soft-tissue sarcoma) | Fracture | [12] |
| Bone (bone and soft-tissue sarcoma) | Growth reduction | [13] |
| Brain (craniopharyngioma) | Cognitive decline | [14] |
| Brain (ependymoma) | Cognitive decline | [15] |
| Brain (low-grade glioma) | Cognitive decline | [16] |
| Brain (medulloblastoma) | Cognitive decline | [17] |
| Cochlea (brain tumor) | Hearing loss | [18] |
| Hypothalamus (brain tumor) | Growth hormone deficiency | [19] |
| Joint (bone and soft-tissue sarcoma) | Motion restriction | [20] |
| Lung (Hodgkin lymphoma) | Pneumonitis | [21] |
| Skin (bone and soft-tissue sarcoma) | Fibrosis | [22] |

##### Physical Dose Differences between Protons and Photons

In proton therapy, charged particles interact with matter and deposit energy in tissue. The physical manner in which the protons interact with tissue is entirely different from that of photons (Figure 2).

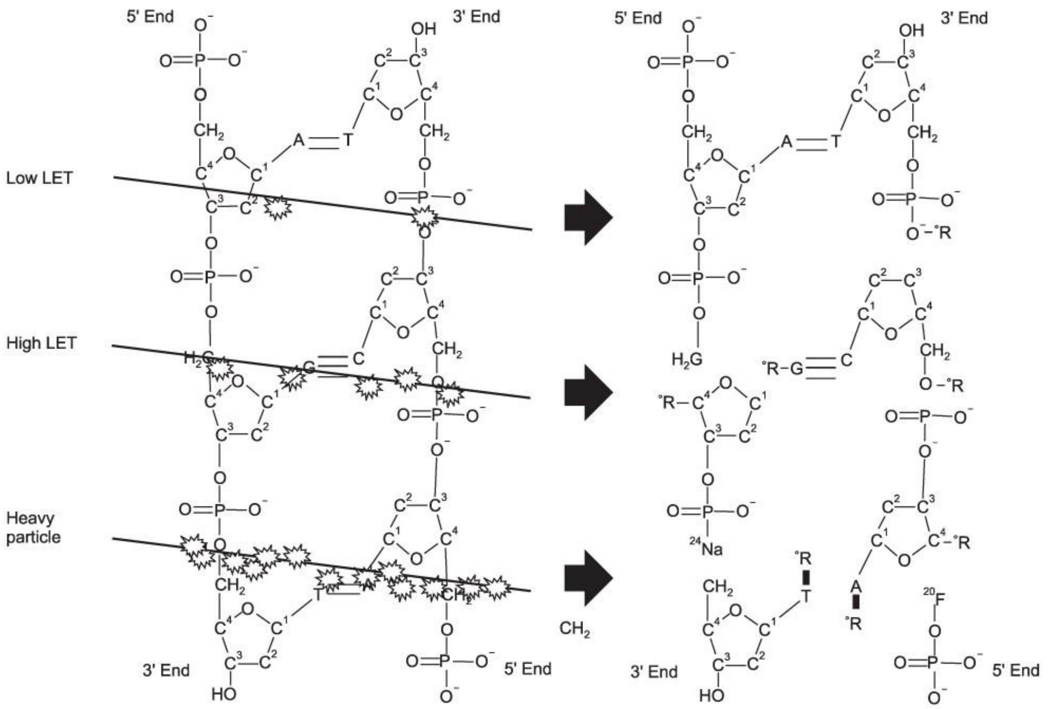

*Figure 2. Relation of energy transfer of radiation (damage) to the frequency of DNA damage per unit length*

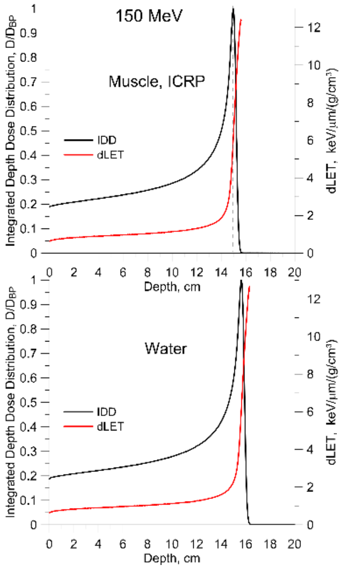

*Figure 3. Relation of dose-weighted linear energy transfer to prescribed depth dose.*

Whereas protons interact with atomic nuclei and outer-shell electrons, efficiently exciting or ionizing atoms with little deflection, photons interact less efficiently with tissue, transferring energy elastically or inelastically to electrons with a higher degree of deflection. The incident energy of the proton determines its maximum range (depth) in tissue; tissue beyond the target is not exposed. In contrast, the incident energy of the photon determines its depth profile in tissue, and tissues beyond the target are always exposed.

Linear energy transfer (LET) describes the transfer of energy of the radiation to tissue as a function of distance. LET is associated with radiobiological effectiveness and is dependent on the cell or tissue type. The relative biological effectiveness (RBE) is the ratio of the biological effectiveness of one type of radiation to that of another, given the same amount of absorbed energy. Both protons and photons are low-LET radiations; however, at the end of the range, the proton LET increases (Figure 3). Areas of increased LET correspond to the most distal and lateral aspects of an applied proton beam. Areas of high LET located in the target may be advantageous [23], whereas areas of high LET located in non-target tissue may result in complications [24]. The normalized product of the dose and LET, the dose-weighted LET (dLET), may be better correlated with complications than LET alone, as high LET in a region of very low dose would not be expected to create a biological effect. In contrast, high LET in the presence of high dose could be of concern, depending on the RBE-LET relation, e.g., of sensitive nervous system tissues. Current methods of treatment planning and plan evaluation do not include an assessment of LET, dLET, or RBE.

Biological Dose Differences between Protons and Photons

The physical properties of photons and protons are well known; however, new developments in proton delivery suggest that there are biological differences that may be relevant to both tumor control and normal tissue toxicity. Whereas photons exhibit only slight variations in LET along their path, protons have more pronounced changes in LET over the particle path as a result of their differential deposition according to depth. This results in the differential biological effectiveness that has been observed in vitro. After the Bragg peak, the dose-weighted LET quickly ramps up and may lead to increased radiation-induced damage, which may manifest as a decreased surviving fraction [25-28] or reduced repair of potentially lethal damage [19, 29-31]. In a study by Guan et al., the differential LET observed proximal to, at, and beyond the Bragg peak varied almost 10-fold [32]. This nonlinear effect suggests that the technical delivery parameters may be optimized to minimize toxicity and enhance efficacy. The most pertinent applications to improve tumor control have been those suggested for radiation-resistant tumors that exhibit regions of hypoxia [23].

Our understanding of how to model the biological effect at depth is poor, and efforts to understand the relation of dose-weighted LET to the clinical context have largely failed to discern any differential toxicity that could be explained by the physical map of the differential RBE and LET [23]. Although toxicity has not yet been observed and verified in the clinical setting, concerns remain, and efforts are being made to better understand the relation of the physical dose, the LET weighted dose, and the RBE to treatment changes in both normal and tumor tissue [24, 28].

The Use of Proton Therapy in Children

Since 1998[1], the Children's Oncology Group, the world's leading pediatric oncology cooperative group, has enrolled only 364 children, adolescents, and young adults on 15 different disease-specific prospective clinical trials, none of which were designed to report proton-specific disease-control outcomes or complications. Clearly, individual proton therapy centers have performed nonprotocol irradiation on substantially larger numbers of patients.

The Particle Therapy Cooperative Oncology Group (PTCOG) reported that 137,000 patients were treated with particle therapy between 1954 and 2014 [33], with pediatric patients representing approximately 10% of the total. The Pediatric Proton Foundation

[1] Data reported 6/20/2015

[34] reported increasing numbers of pediatric patients undergoing proton therapy in the United States, with 465 patients being treated in 2010 and 722 in 2013. As noted in Table 2, most of these patients were not enrolled on cooperative group trials. Disease control estimates and toxicity reporting for children, regardless of participation in a cooperative group clinical trial, are lacking for a number of reasons, including the proportionally smaller number of pediatric patients with cancer, research priorities, and funding constraints.

*Table 1. US pediatric proton therapy statistics 2010–2013.*

| Year | No. of patients enrolled on COG protocols | Total no. of US patients‡ |
|---|---|---|
| 2013 | 49 | 722 |
| 2012 | 45 | 694 |
| 2011 | 45 | 613 |
| 2010 | 19 | 465 |

‡Pediatric Proton Foundation

The lack of enrollment of children on pediatric cooperative group trials and single-institution monitored prospective clinical trials circumvents the established mechanisms for reporting disease control and toxicity information to regulatory agencies. The reporting of outcomes and the reliability of published information is further compromised by the referral of patients to remote centers that are unable to follow patients after treatment.

The D9803 protocol for intermediate-risk rhabdomyosarcoma was the first pediatric cooperative group trial to specifically allow proton therapy [35]. The study was activated in 1999. During the past 15 years, the number of patients treated on COG protocols using proton therapy has steadily increased (Figure 4), and the tumors commonly treated with proton therapy are rhabdomyosarcoma, medulloblastoma, ependymoma, CNS germ cell tumor, and Ewing sarcoma (Figure 5).

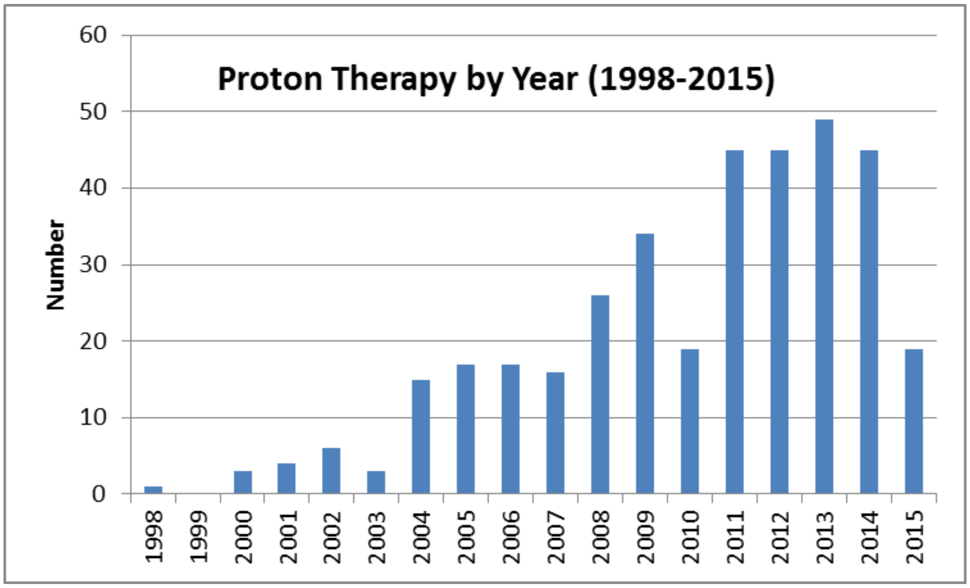

*Figure 4. Patients treated with proton therapy on COG protocols.*

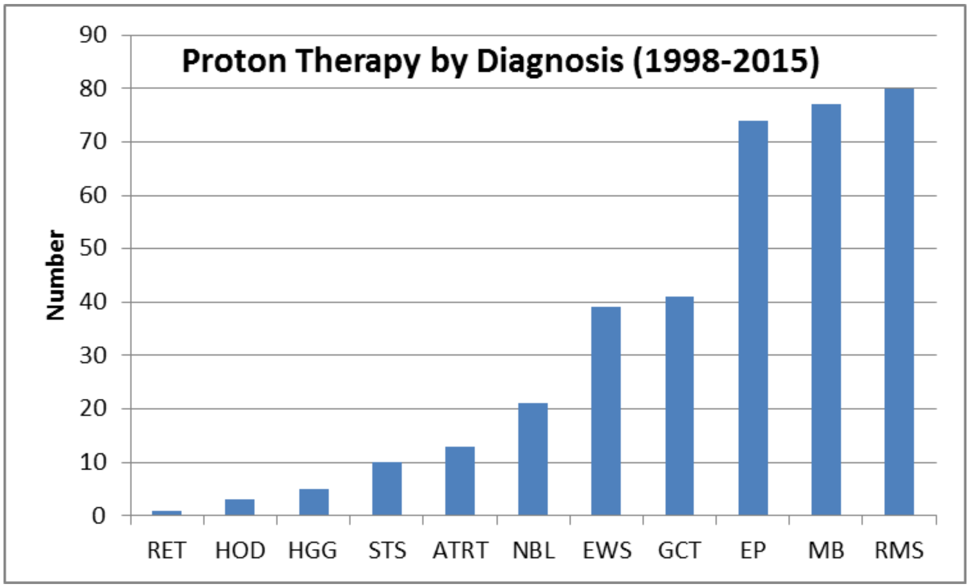

*Figure 5. Tumors treated with proton therapy on COG protocols.*

Legend: RET, retinoblastoma; HOD, Hodgkin lymphoma; HGG, high-grade glioma; STS, soft-tissue sarcoma; ATRT, atypical teratoid rhabdoid tumor; NBL, neuroblastoma; EWS, Ewing sarcoma; GCT, CNS germ cell tumor; EP, ependymoma; MB, medulloblastoma; RMS, rhabdomyosarcoma.

The Role of Changing Technology in Proton Therapy Delivery

Photon therapy for children has evolved during the past two decades with the advent of 3-dimensional treatment planning and, subsequently, 3-dimensional conformal and intensity-modulated photon therapy methods. Proton therapy for children has evolved from limited-conformity, passive-scattering methods that require compensator-based distal range definition and aperture-based collimation to the more advanced and recently introduced discrete spot scanning method, otherwise known as modulated scanning, pencil beam scanning, and intensity-modulated proton therapy (IMPT). Proton spot scanning represents a significant advance over passive-scattering methods, does not require beam-modifying devices, and will continue to evolve as the platform for the foreseeable future, akin to intensity-modulated radiation therapy using photons and its multiple delivery formats. Advances in proton delivery methods, such as the use of pencil beam scanning, now facilitate delivery in a remarkably well-defined distribution [26, 27]. The combined experience with scanning particle therapy in pediatric patients treated at proton centers in the United States represents less than 1500 person years. The unique capabilities of proton therapy and improvements in treatment delivery techniques are not without potential limitations. Poor understanding of the events that occur along the particle path represents a gap in our knowledge of how to equate physical dose with biological effect. Preclinical experiments have shown the potential for an increase in LET along the particle path, which is correlated with reduced cell survival [32]. The implications of this for normal tissue are a potential concern. Are the advantages of this highly conformal method of proton delivery offset by an unpredictable and disproportionate increase in normal tissue damage? The early clinical experience with protons suggests that these effects may be avoided or enhanced by changing the planning techniques that alter the LET distribution [23, 25]. Modeled distributions of LET as it relates to the proton path are theoretical, and clinical observations relating these projections to real findings remain anecdotal.

Proton Therapy and Local Tumor Control

Targeting in radiation therapy is the practiced art of the radiation oncologist. Defining the volume at risk in order to secure local tumor control depends on a number of factors. Large treatment volumes ensure that the area at risk will receive the appropriate radiation dose, but they also increase the risk of treatment-related complications. With the advent of conformal radiation therapy treatment planning and delivery methods, investigators have sought to reduce the target volumes, which include the actual tumor and/or tumor bed (the gross tumor volume [GTV]), the margins surrounding the GTV that account for subclinical microscopic disease (the clinical target volume [CTV] margin), and the margins surrounding the CTV that are meant to account for uncertainties in certain aspects of treatment planning, patient positioning, and beam delivery (the planning target volume [PTV]). Photon therapy and proton therapy share many of the same uncertainties; however, proton beams are more susceptible to differences and changes in tissue composition, as these may alter the path length of the beam and contribute to range uncertainty. The inherent reduction in the dose at depth has the potential to limit the normal tissue and integral dose and the potential secondary complications. If unaccounted for, uncertainties unique to proton therapy might limit tumor control if the estimation of the dosimetry and target volume coverage is insufficient. Inhomogeneity in the target tissue due to the placement of metallic markers or other hardware can also potentially limit the dose at depth [36]. Although no such findings have been documented with proton therapy, reductions in the target margins in photon therapy have been the subject of intensive investigation. Avoiding marginal failures remains a primary concern among radiation oncologists who research ways of reducing the target volume in order to limit toxicity. A notable example is that of single-fraction photon radiosurgery, in which the absence of "dose blurring" as a consequence of the single administration and the reduced target margins inherent to the effort to reduce the volume of tumor that receives a high dose per fraction treatment have led to reports of reduced control rates [37]. Efforts to catalog tumor control rates relative to historical photon-treated cohorts are imperative.

Proton Therapy and Treatment-Related Morbidity and Mortality

Historically, radiation therapy for pediatric cancers has engendered large treatment volumes in many cases, as effective systemic therapies were lacking. While this approach may yield improved short-term control rates at disease-related time points, the late complications of therapy have resulted in a "second slide" in survival due to a combination of damage to normal tissue and second cancers [38]. Whereas large-field treatments persist in specific clinical situations (e.g., treatment of the whole lung or flank) or in scenarios involving metastatic disease or tumor spillage into body cavities, the radiation therapy methods applied to most tumors include volumetric planning and targeting, with the goal of delivering the prescribed dose to the target [29]. The use of multiple photon conformal therapy methods has led to increased integral doses. Proton therapy has the potential to reduce the volume of tissue receiving higher doses but also the lower doses mentioned above [31]. There may be real differences in the normal tissue injury–related effects and second cancers that result from proton and photon therapy, with corresponding long-term effects.

Recent reports of unexpected clinical findings or toxicity related to proton therapy in pediatric patients with brain tumors [30, 39-41] have raised concerns about the guidelines for proton therapy in children and have highlighted uncertainties regarding proton therapy physics and the related biological effects.

The Role of Clinical Trials in Evaluating Treatment-Related Adverse Events

There have been few efforts to systematically assess the impact of differential particle energy deposition at depth. Some have proposed the use of registries to improve our understanding of the clinical implications of proton therapy (Table 3).

*Table 2. Current efforts to study the efficacy and safety of proton therapy.*

| **Study** | **Study type** | **Disease/Condition** | **Sponsor** | **Identifier** |
|---|---|---|---|---|
| Proton normal tissue toxicity | Registry | NA | MDACC | NCT01502150 |
| QOL, normal tissue toxicity, & outcomes in proton therapy | Registry | NA | Samsung Medical Center | NCT02644993 |
| PPCR | Registry | NA | MGH | NCT01696721 |
| CN01 | Registry | Pediatric brain, head, & neck | Univ. Florida | NCT01067196 |
| Proton rhabdomyosarcoma | Phase II | Rhabdomyosarcoma | MGH | NCT01115777 |
| Proton neuroblastoma | Phase II | Neuroblastoma | MGH | NCT02112617 |
| Proton bone and non-RMS STS | Phase II | Bone and non-RMS STS | MGH | NCT00592293 |
| Proton mediastinal lymphoma | Registry | Lymphoma | MGH | NCT01751412 |
| Proton partial brain RT | Registry | Brain tumors | MGH | NCT01288235 |

Published reports of outcomes gleaned from registries have addressed certain questions, including the incidence of second cancers in particle therapy patients [42] and general disease site–specific outcomes. However, the registry approach has insufficient depth and lacks a systematic estimation of the impact of relevant factors that may relate to proton therapy toxicity. Furthermore, registry studies lack rigorous statistical design. Others have argued for disease site–specific phase III trials to compare proton and photon therapy. These efforts, however, are limited by numerous practical problems, including patients' refusal to randomize, potential violations of clinical equipoise, and the difficulty of obtaining a sufficient sample size to power the study for endpoints that are assumed to differ across treatment modalities. Phase IV clinical trials offer an attractive approach in that they are directed as systematically characterizing toxicity in a broader non–disease site–specific manner. Furthermore, they offer the opportunity to review the impact of other contributing factors, which in the case of radiation therapy may include a number of technical and treatment-related factors.

We propose to evaluate tumor control and treatment-related complications in children receiving proton therapy with reference to historical cohorts treated with photons on institutional protocols for well-represented disease-specific subsets.

**Rationale**

Rationale for CTCAE as a Toxicity Scale

To facilitate the recognition, grading, and communication of therapy-related adverse events, the “Common Terminology Criteria for Adverse Events” (CTCAE) were developed in 1983 [43]. The second version of these criteria (CTCAE v2.0), published in 1998, featured improved grading of acute effects.

Discord between the concurrently used WHO, RTOG/EORTC, and LENT scales prompted additional revisions at the LENT IV conference in Petersburg, Florida, in 2002 [44]. During this conference, there was added support for revision from a multidisciplinary team spanning the NCI, the pharmaceutical industry, biologists, all oncologic subspecialties, and statisticians. This conference oversaw the incorporation of surgery- and radiation therapy–related adverse events, thus making the CTCAE one of the few multimodality grading systems for reporting the acute and late effects of cancer treatment (see Table 4). Although this system was designed to span modalities, the toxicities included are not decidedly modality-specific, thereby avoiding the principle of attribution, which may often be problematic or multifactorial.

For the field of pediatric oncology, the most substantial update at this conference was the incorporation of late effects and pediatric criteria [45], which was made possible by the attendance of several members of the Children’s Oncology Group. Multiple existing late-effect criteria/gradings were incorporated during the conference [46, 47]. The resulting system codified event reporting into a systematic common parlance for use in comparisons across clinical studies and was ultimately endorsed by multiple organizations involved in conducting clinical research and trials (RTOG, EORTC, ACOSOG, ESTRO, and ASTRO).

*Table 3. Evolution of toxicity grading systems.*

The Evolution of Toxicity Grading Systems(1979-1998)

| *System* | *No. of Criteria* | *No. of Organs* | *Modality* | *Phase* |
|---|---|---|---|---|
| WHO (1979) | 28 | 9 | Chemo | Acute |
| CTC (1983) | 18 | 13 | Chemo | Acute |
| RTOG/EORTC-Acute (1984) | 14 | 13 | RT Acute | Acute |
| RTOG/EORTC-Late (1984) | 16 | 13 | RT Late | Late |
| LENT (1995) | 152 | 22 | RT Late | Late |
| CTC v 2.0 (1998) | 260 | 22 | All* | Acute |

Abbreviation: WHO, World Health Organization.
*Limited pediatric and surgical criteria.

The LENT IV conference defined toxicity according to the severity and temporal nature of the adverse event. CTCAE v3.0 defined grade 3 as reflecting severe and very undesirable events that prompted interventions, surgery, or hospitalization. Grade 4 toxicities were defined as potentially life threatening, catastrophic, disabling, or resulting in the loss of an organ, organ function, or limb. The temporality of an effect has always been difficult to define, as an adverse event may occur at varying times and to varying degrees in patients. For this reason, the definition of an acute versus a late effect has been removed from the CTCAE, and any permanent disabling event is deemed a late event. Furthermore, the duration of an adverse event was defined according to serial chronological assessments rather than by assigning a grade for each anticipated or preceding duration.

As with any graded objective scale, clinical relevance or impact on the quality of life is poorly correlated. The CTCAE criteria are decidedly not a means of defining an acceptable toxicity profile, as different patients or populations may have different definitions of what constitutes an acceptable adverse event. For this reason, the NCI has commissioned and developed a means of collecting patient-reported outcomes that gauge the physical, mental, and social well-being of an individual. Whereas the Patient Reported Outcomes Measurement Information System (PROMIS) has been validated for the adult population, the pediatric scale has not yet been validated and will be considered for inclusion as an amendment in a future revision of the protocol.

We have proposed the CTCAE v4.0 serial grading and reporting of CTCAE grade > 3 nonhematologic toxicities across all patients treated with proton therapy. The serial toxicities will be reviewed and compared to historical controls who underwent photon therapy. When historical photon-treated cohorts are unavailable as a result of insufficient sample size, we will estimate the toxicities for that population.

Rationale for Timing and Duration of Follow-up

The pediatric population is especially vulnerable to both acute and late toxicities, many of which are not apparent during and immediately after treatment. Furthermore, whereas radiation therapy causes only minimal acute effects in many cases, late effects in developing children may be severe, deforming, or limiting with respect to function. There are contrasting opinions on the type, severity, and extent of proton therapy side effects as compared to photon therapy side effects. Many argue that the limited integral radiation therapy dose suggests that toxicity might be reduced by limiting the applied dose. Others argue that the physical properties and inhomogeneities in the LET and RBE discussed above and visually represented in Figure 3 may lead to untoward severe late effects that would not be expected with a similar physical dose in photon therapy. Thus, rigorous prospectively specified serial evaluations are needed to define the incidence, duration, and chronicity of each event.

Although at least one late effect is described in most survivors of childhood cancer treatment, few details are available regarding the duration, chronicity, and means of managing the event. Many late effects could be classified in terms of their relation to the symptomatic burden, impact on function, contribution to current health status, and ultimate role in determining late mortality. Thus, although toxicity grading will provide crude descriptive measures of the event, further classification will aid our understanding of how to mitigate these late events [48]. For example, technical delivery of radiation therapy is likely accomplishable via any number of potential beam angles, path lengths, doses, and techniques (passive scatter, pencil beam scanning, etc.). The path of entry may have unrealized profound implications for late effects in the form of vasculopathy [14], radiation necrosis [49], second cancers, or vital organ function (e.g., cardiac wall motion abnormalities or accelerated neurocognitive decline) [50]. Thus, serial evaluation of not only the type of toxicity but also the relevant limitations bestowed by that toxicity will be essential to reducing the late burden of cancer care.

Most late effects develop with varying degrees of severity within 5 years of treatment; however, perhaps the most severe late effect, treatment-related cancer, will not become apparent until 5 to 10 years after therapy [51]. Furthermore, the natural history of second cancers suggests that although most second cancers present during the 5 to 10–year window, the risk of second cancers does not reach a plateau.

Rationale for Vasculopathy Assessment

Pediatric patients with brain tumors treated with cranial radiation have the potential to experience significant late morbidity. Vascular complications can take many forms, including stroke, vascular occlusion with resultant Moyamoya syndrome, and hemorrhage. Several long-term studies of pediatric patients with brain tumors treated with photons have documented the incidence of stroke as 5.6% at 25 years after treatment [52, 53]. This corresponds to approximately 4 cases per 1000 patient years. The impact of cranial radiation therapy on late mortality in pediatric patients as a result of vasculopathy is second only to the excess mortality from second cancers. Circulatory disturbances from vascular injury have been estimated to cause an absolute excess risk of 2 extra deaths (95% CI 2–3) per 1000 person years at 5 to 14 years after diagnosis and an absolute excess risk of 29 excess deaths (95% CI 16–56) at more than 45 years after diagnosis [54]. These results should be compared with the absolute excess risk of death from second cancers of 8 extra deaths (95% CI 7–10) at 5 to 14 years after diagnosis and 58 extra deaths (95% CI 38–90) at more than 45 years after diagnosis.

The risk factors for cranial radiation–induced vasculopathy include the patient's age at treatment, the volume of brain treated, the dose of radiation used, and the coincident or sequential use of chemotherapy [55-58]. The risk of vasculopathy secondary to proton therapy relative to that with photon therapy is unclear, as there are no long-term data on this issue. In a review of 101 patients treated for craniopharyngioma, the cumulative incidence of abnormal MRA findings at 3 years was 16% ± 5.82% vs. 21.8% ± 4.14% for the proton therapy and photon therapy cohorts, respectively. Correspondingly, the 3-year cumulative incidence of clinically significant vasculopathy was 2.22% ± 2.22% vs. 3.98% ± 1.96% for the proton therapy and photon therapy cohorts, respectively. Conversely, our brain tumor study, SJYC07, yielded an excess risk of vasculopathy events for the proton cohort relative to the photon therapy cohort, although this comparison is limited by the small sample size and by heterogeneities in the patient and tumor-related factors, as well as clinical risk factors.

Rationale for Necrosis Assessment

In many cases, higher doses of radiation are required to control local disease, resulting in an increased risk of parenchymal necrosis. Although necrosis is typically a pathologic phenomenon, intracranial radiation necrosis is typically diagnosed after a review of clinical and radiographic findings, and it is rare that a pathologic diagnosis is pursued. Clinical symptoms may be nonexistent; subtle without significant morbidity; prominent, requiring medication; or refractory to medications, possibly leading to death. Symptoms are largely related to the severity and location of the insult. As such, the presenting symptoms vary and can be as mild as transient fatigue or more substantial with refractory seizures. The burden of radiation necrosis on pediatric patients is, thankfully, limited, as the cumulative incidence is less than 5% across all intracranial tumors [59]. The cumulative incidence of serious injury related to radiation necrosis is even lower, with approximately 2% of patients experiencing toxicity of grade 3 or higher.

The management of symptomatic radiation necrosis may include the use of steroids, antioxidants and anti-inflammatories, anti-vascular agents such as bevacizumab, and/or hyperbaric oxygen [60, 61]. Complicating the matter is the potential for therapies such as hyperbaric oxygen to facilitate early recurrence, as hyperoxic conditions are thought to be tumorigenic relative to the anoxic/hypoxic conditions present in many high-grade brain tumors [62]. Additionally, the level of evidence for the management of radiation necrosis is largely based on small series and professional opinion with little or no randomized data [63]. Efforts to expand our knowledge of the contributing factors and possible mitigating strategies are needed.

Radiation necrosis secondary to radiation therapy is related to the dose and volume treated, as well as to the presence or absence of peri–radiation therapy chemotherapy. The impact of the type of radiation therapy (proton vs. photon) on the risk of radiation

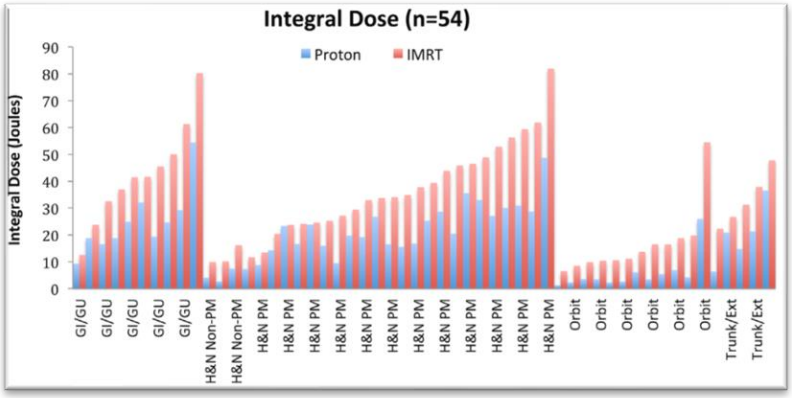

*Figure 6. Integral dose across disease sites in rhabdomyosarcoma*

necrosis is unclear, although some series have shown a shorter time to development of necrosis with proton as opposed to photon therapy [61]. The same study also identified a differential effect of age and the use of protons on the risk of radiation necrosis, with younger children being more susceptible to developing necrosis. Because of the inherent differences in the biology, type, and location of intracranial tumors in children, testing the effect of age on the risk of radiation necrosis may be challenging in the absence of a true matched cohort [64].

The literature on the relative risk of radiation necrosis with photons and protons is sparse. In our review of 101 patients treated for craniopharyngioma with photons and protons, we identified 3-year cumulative incidences of 1.98% ± 1.39% with photons and 2.68% ± 1.89% with protons (unpublished data). Conversely, we observed an excess of cranial radiation necrosis and vasculopathy in our infant brain tumor study, SJYC07. Confounding factors such as age, dose, volume, and the use of chemotherapy likely affect the risk and differential findings across studies; therefore, further study is needed.

Rationale for Assessment of Treatment-Related Malignancies

Radiation therapy–associated secondary cancers represent a clinically challenging subgroup of malignant neoplasms with a dismal prognosis. In fact, relative to primary cancers of the same tissue of origin and histologic subtype, second cancers have an estimated survival rate of only a quarter to a half of that reported for their comparison cohort. Many childhood cancers now have a dramatically different prognosis compared to just 20 years ago, so coordinated efforts to understand severe late effects, such as terminal second cancers, are crucial to stopping the second slump in survival that is seen in survivors of childhood cancer [65, 66].

Proton therapy offers the potential to reduce the integral dose to uninvolved tissues [67]. “Integral dose” refers to the total energy (in Joules) deposited over a volume of tissue (in $cm^3$) with a specified mass (in grams) (Figure 6). Prior studies have demonstrated that proton therapy has an advantage over IMRT at multiple disease sites, which may have profound implications for the risk of second cancers [68, 69]. A relation between the integral dose and the risk of second cancers has not been established, although the concept has a sound rationale.

Rationale for Characterization of Pediatric Intracranial Treatment-Related Tumors

To date, no study has systematically evaluated the incidence of treatment-related second cancers relative to that in patients treated with photon therapy. The characteristics of megavoltage energy treatment–related mutational events are poorly described, although preliminary data indicate the clear limitations of describing these mutational events, including the variable probability of the event according to treatment-dependent and treatment-independent factors [70]. Treatment-independent factors include the patient’s lifespan, tissue type, age, and genetic background. Treatment-related factors such as the length of exposure, the irradiated volume, and varied radiation energy compound the complexity of describing the probability of mutagenesis.

Analyses of mutational types based on next-generation sequencing data from radiation-associated cancers have revealed a distinctive mutational signature that is not typical of other genotoxic mutagens. For example, although ultraviolet light exposure has been known for some time to cause characteristic mutational signatures [71], megavoltage radiation appears to be capable of causing one of three distinct mutational signatures, each with characteristic types of base substitutions. Furthermore, different tumor types have differential base-substitution patterns that appear to be contingent on the tissue type irradiated [72] and on the genetic background [73].

Radiation-associated cancers also appear to show distinctive copy-number abnormalities that inform the clinical behavior, latency, and treatment resistance of the cancers [73]. In an analysis of Chernobyl survivors with papillary thyroid cancer, copy-number gains of characteristic chromosomal regions [74] were common to 29% of cases. Similar changes in the same chromosomal region [74] have been observed in a similar proportion of post-irradiation sarcomas [75]. Additionally, copy-number gains of well-known tumor suppressor genes, including NF2 and CHEK2, were noted in cases of thyroid cancer with prolonged latency [73]. Correspondingly, increases in RET amplification are associated with reduced latency in Chernobyl-associated thyroid cancers [76]. Similarly, prior reports from our institution noted that patients with irradiation-associated diffuse intrinsic pontine gliomas had characteristic copy-number alterations, including gain of 1q, and PDGFR amplification at recurrence [77]. Therefore, further understanding of the incidence of and contributing factors to treatment-related second cancers will facilitate risk reduction in radiation therapy planning and improve the subsequent management and treatment of these challenging malignances.

Rationale for Dose-Weighted Linear Energy Transfer Modeling in Local Control and Toxicity

Dose-weighted linear energy transfer (dLET) is a useful quantitative criterion for late-effect analysis in proton therapy with the use of beam-spread devices, a patient-specific aperture, or compensator (AC). In our early attempts to model the effect of dLET by using intensity-modulated proton therapy (IMPT) with a discrete spot scanning beam system without the use of an AC, we identified differential effects of double scatter vs. scanning techniques at the distal edge of the Bragg peak (Figure 7). The implications of this for late effects are unknown, although preliminary findings from clinical studies suggest there is no adverse impact on treatment-related toxicity across central nervous system malignancies [78]. It has been suggested that the differential impact of protons at the distal range may have advantages and can be optimized for treatment planning and tumor killing [23]. In brief, dLET maps will be garnered for all patients treated with proton therapy in cases where toxicities are observed. Furthermore, disease-specific subset analyses to estimate the impact of the volume and location of dLET on tumor control are planned.

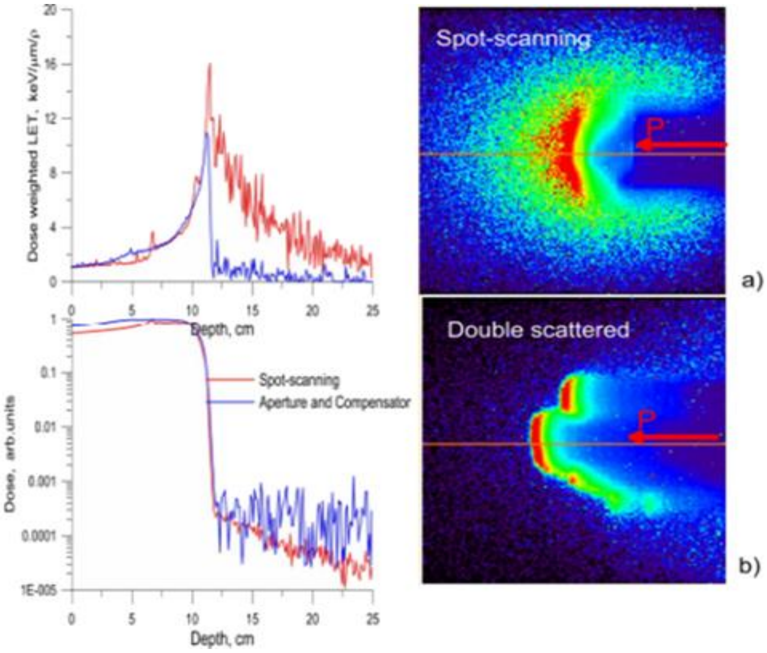

*Figure 7. a) Depth dose and dLET distribution for spot scanning and double scattered beam. b) TOPAS simulation of dLET from clinical fields. IMPT with spot scanning and double scatter with an aperture and compensator.*

Rationale for Assessment of Treatment-Related Deaths

Approximately 82% of patients who are treated for childhood cancer in the United States will be cured [79]. As treatments have improved, the underlying late morbidity of these efficacious interventions has been unmasked. Unfortunately, patients without adequate support or sufficient health maintenance services may have increased potential for early mortality as a result of treatment-induced late morbidity [80-83]. Conversely, these observations have been instructive and have led to a systematic reduction in those treatments that are thought to result in disproportionately high morbidity [84-87]. Chief among the interventions yielding undue morbidity is radiation therapy, although it is likely that no modality has been more systematically refined to improve the therapeutic ratio. Multiple strategies have been employed to reduce the radiation therapy dose and volume used in pediatric patients with cancer. These strategies have included response-adapted therapy in Hodgkin lymphoma [88], the use of induction chemotherapy to facilitate radiotherapy dose reduction in germ cell tumors [89], the use of concurrent and adjuvant chemotherapy to reduce the dose of craniospinal radiation therapy in patients with medulloblastoma [90], and the systematic reduction of the target volume margins with the advent of conformal photon and proton therapy [91]. Although the impact of each of these strategies is nearly impossible to demonstrate because of multiple confounders (population differences, the era of treatment, concurrent therapies, the genetic background, etc.), it is clear from studies that were less restricted with regard to the disease site that substantial morbidity reductions have been possible with the systematic reduction in radiation therapy use, dose, and volume [84]. Rigorously documenting treatment-related mortality by using established criteria will help our understanding of the role of radiation therapy in treatment morbidity and mortality, as well as guide further refinements of the therapeutic ratio [92].

Rationale for Evaluating the Role of Treatment Delivery and Planning Parameters and Their Impact on Local Control and Toxicity

The evolution of radiation therapy delivery techniques has brought about a sea change in our understanding of clinically acceptable therapeutic targets. Before the advent of 3-dimensional imaging with CT or MRI, crude bony borders were used to delineate a space approximating the normal tumor location (Figure 8A). Although this was practical and efficacious, given the absence of volumetric imaging, the therapeutic ratio was poor as a result of the large size of the field, which increased the morbidity associated with the treatment and limited the delivery of an adequate therapeutic dose. Target definitions were crude and were limited by the inability to define specific anatomic regions (ICRU-29).

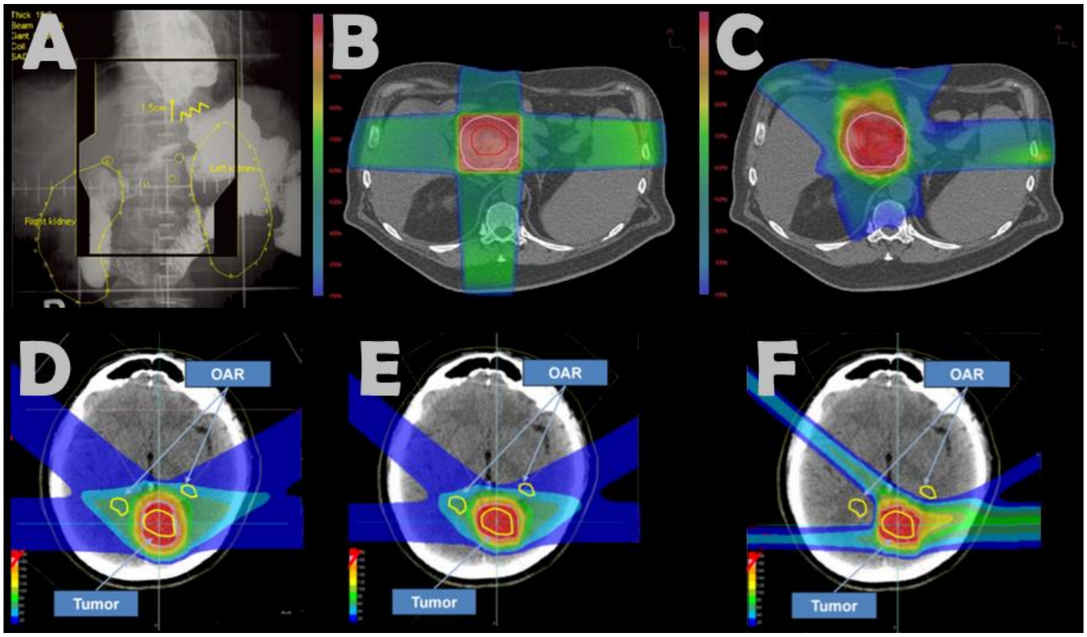

*Figure 8. Treatment delivery and planning according to modality and technique. A. Two-dimensional X-ray–based target delineation. B. Three-dimensional conformal radiation therapy using CT-based planning. C. Three-dimensional planning with an intensity-modulated photon radiation delivery technique. D. 3D Passive Scatter Proton. E. Pencil Beam Scanning Single Field Uniform Dose. F. Intensity Modulated Proton Radiotherapy*

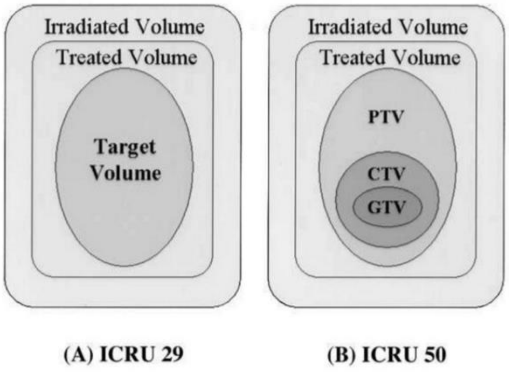

*Figure 9. ICRU target volume definitions.*

The shift to 3D planning led to a broader discussion as to what constituted the anatomic target in DICOM space. ICRU-50 (Figure 9) specified the prescription of a radiation therapy dose to a volume of tissue delineated on anatomic imaging rather than to a region based on 2D planning. ICRU-50 also defined our modern understanding of target volumes, including the GTV, CTV, PTV, treated volume, and irradiated volume. The GTV was designated to represent the extent and location of the malignant lesion. The GTV could be subdivided into GTVs specific to the primary tumor (GTV primary), lymphadenopathy (GTV nodal), or metastasis (GTV M). The GTV could be defined by imaging, clinically via a physical examination, or as visualized via alternative means. Uncertainties in tumor delineation led to the broad acceptance of the CTV concept, which added a margin around what was visible by imaging and was meant to encompass the region of spread that was not readily identifiable on anatomic imaging. Two types of subclinical extension were defined; extension from the primary tumor into surrounding normal tissues and extension from involved lymphatic deposits that are readily apparent on imaging. The CTV is often anatomically constrained and defined by known patterns of spread or by limiting anatomic barriers, such as bone or cavities. The PTV was defined to be an isometric expansion around the CTV, representing the net effect of all possible geometric variations in space due to motion (breathing, swallowing, and set-up error) or inaccuracies in the target delineation process (misregistration). The PTV is affected by internal motion of the organ or tumor and by the treatment technique (beam orientation, patient fixation, set-up error). The errors can be intrafractional errors (occurring during a single treatment session) or interfractional (occurring from one treatment session to another). The treated volume (TV) is defined as the volume enclosed by the prescription isodose surface (usually the 95% isodose curve). The TV is intended to be as close to the volume of the PTV as possible, but limitations in techniques were prohibitive until the adoption of IMRT. For instance, although the PTV is the same in Figure 9 and Figure 10, the treated volumes differ as a result of the adoption of multiple beam angles and the modulation of the intensity of the field at each gantry position. The irradiated volume (IRV) is defined as the volume that receives a dose considered to be significant in relation to normal tissue tolerance. Historically, this volume has been defined as the volume of the 50% isodose curve, although this definition may need to be redefined according to the adverse event type. This is the volume that is most heavily influenced by the treatment technique, as shown in Figure 8D, E, and F. Even across proton therapy plans, divergent delivery techniques (passive scatter, single-field optimized spot scanning, and multi-field optimized spot scanning) have resulted in substantial discrepancies in the IRV.

Over time, the technological improvements in CT, PET, and MR imaging have led to further refinement and increased confidence about what constitutes the tumor on anatomic imaging and what volume adequately approximates the geometric variability of a target region, accounting for motion. Four-dimensional CT and MR imaging techniques have been instrumental in defining the anatomic envelope of motion within which a tumor or at-risk target region moves in DICOM space. For this reason, ICRU-62 added the concept of an internal target volume that encompasses the CTV and internal margin. The internal margin (IM) is defined as variations in the size, shape, and position of the CTV relative to anatomic reference points (Figure 10).

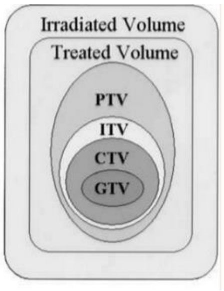

*Figure 10. ICRU-62 target definitions*

The successive evolution of treatment, delivery, and target delineation methods necessitates continual re-evaluation, as unanticipated limitations can lead to discrepancies in the planned and delivered radiation therapy dose, potentially increasing the risk of treatment failure. For instance, contrast is commonly used in simulation planning because it can aid in delineating target and normal structures (Figure 11).

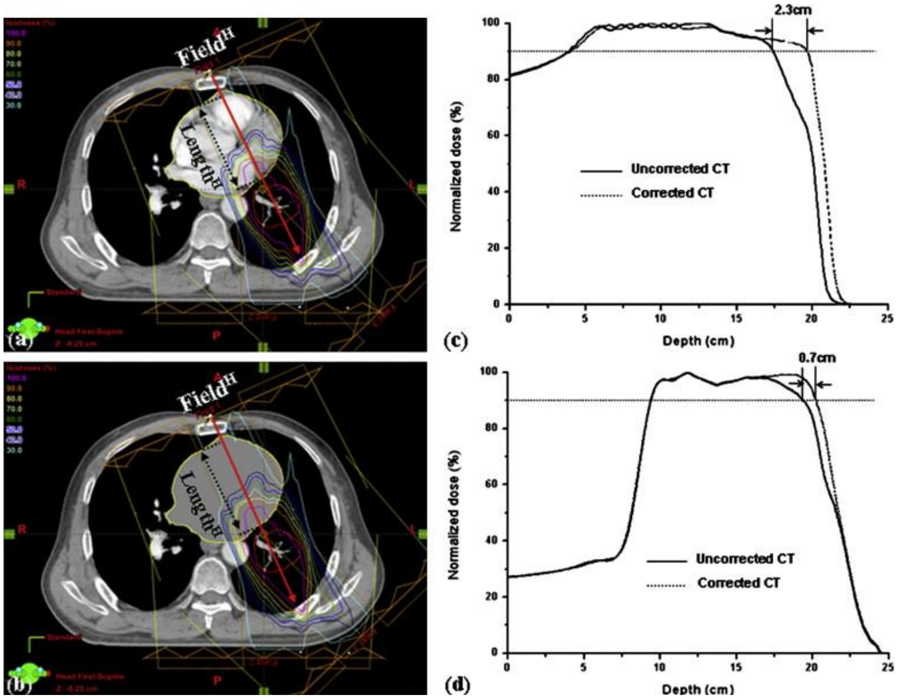

*Figure 11. Treatment plan for Patient 2, showing the distal fall-off of the proton beam in lung tissue. (a) Plan using uncorrected CT images. (b) Verification plan using corrected CT images. (c) Comparison of depth-dose curves along the red line of the un-corrected images. (d) Comparison of depth-dose curves along the red line of the corrected images. [93]*

However, the increased density leads to a difference in the measured Hounsfield units, which are used to determine the attenuation of the radiation therapy dose across tissue. In this case, the increased attenuation was temporary, as the contrast cleared within minutes of the planning scan time. An uncorrected scan can lead to a divergent distribution of the planned radiation therapy dose, potentially leading to an increased dose to organs at risk, and thus to untoward effects, or to a decreased dose to the target structures, potentially resulting in treatment failure. Similar discrepancies have been noted when surgical clips lie in the beam path.

Additional discrepancies between the delivered and intended doses may arise when there is organ motion. The “interplay effect” is a phenomenon in which intensity-modulated photon fields have different dose distributions depending on the phase of treatment delivery and the phase of organ motion. Although the interplay effect has been discussed in the photon IMRT literature, adverse clinical outcome correlates have not been observed. The effect may be magnified, however, in the proton literature because of the increased sensitivity of the dose distribution to the path length and the energy of the protons (Figure 12). With a single fraction, the prescription dose is delivered to only 85% of the treatment volume. Although this effect is reduced with fractionation, regions of tumor may be under- or overdosed, depending on the phase of the proton scanning and organ motion. For this reason, uniform-field dose plans have been favored for thoracic locations and pencil beam scanning is only now being prospectively evaluated in areas with anticipated organ motion [94].

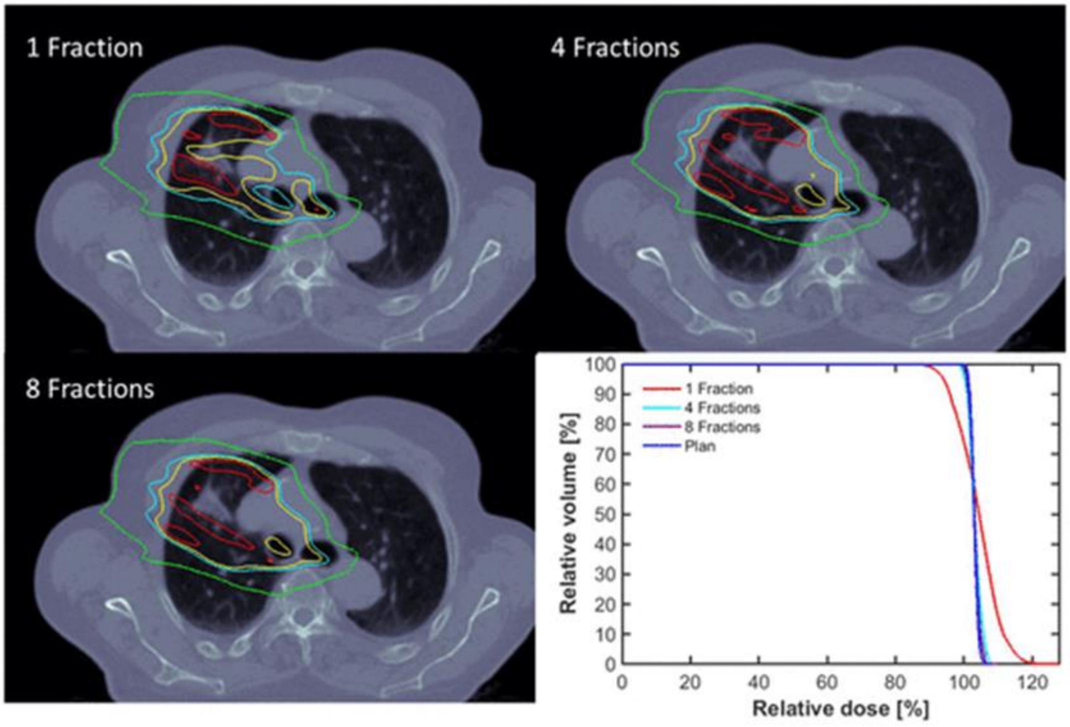

*Figure 12. Interplay effect in thoracic malignant tumors according to the number of fractions. Increasing fractionation leads to improved target coverage, as represented in the dose-volume histogram at bottom right.*

To understand the treatment delivery–related factors related to disease control when proton therapy is the treatment modality, we proposed incorporating the following definitions of treatment failure in the protocol: central failure, in-field failure, marginal failure, out-of-field failure, and distant failure. Central failure is defined as progression within the >95% prescription isodose line. In-field failure is defined as progression within the 80% prescription isodose line that is not completely encompassed by the 95% prescription isodose line. Marginal failure is defined as progression within the 20% to 50% prescription isodose line. Regional failure is defined as progression within the first-, second-, or third-echelon draining lymphatics. Distant organ failure is defined as progression outside the 20% isodose prescription volume but within the treated organ of tumor origin. Distant extra-organ failure is defined as failure outside the organ of tumor origin. Treatment patterns of failure will be reported according to matched photon-treated cohorts from prospective trials conducted at St. Jude Children's Research Hospital.

#### 3.0 RESEARCH PARTICIPANT ELIGIBILITY CRITERIA AND STUDY ENROLLMENT

In accordance with institutional and NIH policy, the study will enroll research participants regardless of gender or ethnic background. Our institutional experience confirms broad representation in this regard.

##### 3.1 Inclusion Criteria

3.1.1 The patient is planned for treatment or has been treated with proton therapy at St. Jude Children's Research Hospital on or after November 18, 2015.

##### 3.2. Exclusion Criteria

3.2.1 Pregnant female patients will not be enrolled in the study and are thus excluded. Radiation has teratogenic or abortifacient effects.

##### 3.3 Research Participant Recruitment and Screening

3.3.1 Recruitment

All patients for whom proton therapy is planned, regardless of indication or setting, will be considered for enrollment. If a patient is planned for proton radiotherapy but is subsequently switched to photon radiotherapy due to technical constraints or patient preference, their screening will be documented but will be subsequently removed from the trial. Although it is preferred that patients enroll at the time they consent to proton therapy, enrollment will also be allowed at other time points. Patients who have already completed therapy will be allowed to enroll retroactively.

##### 3.4 Enrollment on Study

A member of the study team will confirm potential participant eligibility as defined in Section 3.1-3.2, complete and sign the 'Participant Eligibility Checklist'. The study team will enter the eligibility checklist information into the Patient Protocol Manager (PPM) system. Eligibility will be reviewed, and a research participant-specific consent form and assent document (where applicable) will be generated. The signed consent/assent form must be faxed or emailed to the CPDMO at 595-6265 in order to complete the enrollment.

The CPDMO is staffed 7:30 am-5:00 pm CST, Monday through Friday. A staff member from the Milli helpline is on call Saturday, Sunday, and holidays from 8:00 am to 6:00 pm. If you have a prospective research enrollment and need assistance releasing your consent, please call the Milli helpline on call number (901-338-0596).

#### 4.0 TREATMENT PLAN

##### 4.1 Treatment

The proton therapy dose, fractionation, and schedule on SJPROTON1 will be specified at the discretion of the treating physician or in accordance with other disease- or site-specific protocols. Target volumes and normal tissue structure names will be transformed to conventions applied to this study.

##### 4.2 Dose Modifications

Radiation therapy dose modifications, early treatment discontinuation, and any other change in the radiation therapy plan will be at the discretion of the treating physician and will be subject to the recommendations of any concurrently enrolled disease site– or body region–specific treatment protocols. Any modifications to the projected treatment plan made before the onset of therapy should be documented. Specific reasons should be stated for the treatment plan modification, and corresponding information that supports the modification should be included in the modification report. (Refer to the Appendix.)

##### 4.3 Definitions of Dose-Limiting Toxicity

In the event of grade 4 or 5 toxicity that is possibly attributable to proton therapy, a thorough evaluation of the dose delivery, treatment plan, imaging/laboratory correlates, possible contributing factors, and disease status, along with a comprehensive evaluation of patient symptoms, will be documented and reported.

##### 4.4 Concomitant Therapy

Concomitant therapies are permitted within the context of this protocol as long as they are permitted on concurrently enrolled disease site–specific protocols and/or are considered acceptable according to the non-protocol treatment standards of the institution.

##### 4.5 Management of Treatment-Related Effects

When early signs of progressive parenchymal changes are present on imaging, we may consider a referral for hyperbaric oxygen therapy (HBOT). HBOT will be recommended when progressive parenchymal changes are associated with symptoms, regardless of severity. Steroid therapy, most often with dexamethasone, may be initiated and tapered according to the symptoms. When the dose of dexamethasone has been tapered to approximately 0.5 mg daily, a taper of hydrocortisone will be initiated at approximately 25 mg daily, administered in divided doses. Dexamethasone will be discontinued within 2 to 3 days of initiating hydrocortisone. Patients are not required to remain on steroid therapy during HBOT. The use of HBOT or corticosteroids does not mean that the patient has radiation necrosis, as both approaches are used to treat non–radiation-induced normal tissue damage resulting from mechanical, ischemic, or other toxic insults.

#### 5.0 PROTON THERAPY

The guidelines for proton therapy have been developed to ensure coverage of the volume at risk and to minimize the side effects of treatment. The guidelines are standard in terms of dose and fractionation. There is limited data concerning disease control and functional outcomes after proton therapy administered with systematic targeting. None of the published reports concerning proton therapy describe in detail the method of targeting, immobilization, or verification, and there are no reports on treatment assessment of target volume deformity. Proton therapy is not considered investigational; it is FDA cleared and is covered by third-party payers. Intensity-modulated proton therapy will be used for all patients. Intensity-modulated proton therapy using discrete spot scanning is the newest form of proton therapy and includes intensity modulation with iterative planning, as well as single-field uniform dose methods. As a requirement, the intermittent use of photons in the event of proton system unavailability will be accounted for in the data set. The guidelines for this study are generalized to provide consistency in targeting, dosimetry, and reporting. The investigator's discretion will be used for minor aspects of treatment, including dose conformity and normal tissue dose-volume constraints.

##### 5.1 Indications for Proton Therapy

All patients meeting the eligibility criteria will be treated with proton therapy. Some patients may be treated with photon therapy for a portion of their treatment course.

##### 5.2 Timing

Radiation therapy will be delivered according to the timing specified by the concurrent disease-related protocol, the physician's discretion, the disease site–specific nonprotocol treatment, or institutional standards or as determined by logistical issues related to patient care.

##### 5.3 Emergency Irradiation

Patients are allowed to have received fractionated photon radiation therapy before enrolling on this protocol, and urgent irradiation will be permitted, but this is unlikely to be necessary, given the extensive planning related to most proton therapy cases.

##### 5.4 Equipment and Methods of Delivery and Verification

| Equipment | Protons | Photons | Electrons | α, β, γ |
|---|---|---|---|---|
| Proton beam | X | | | |
| Linear accelerator | | X | X | |
| Brachytherapy | | | | X |

Combined proton and photon treatment is permitted. Some patients may receive photon therapy in lieu of proton therapy if the proton system is unavailable.

##### 5.5 Techniques and Equipment

The principals of 3-dimensional image-based planning will be used for patients treated on this protocol, with the objective of achieving PTV coverage and sparing normal tissues from the highest doses.

##### 5.6 Treatment Planning

CT (volumetric)-based planning is required to optimize the dose to the target volumes while protecting normal tissues. Organs at risk within and external to the irradiated volume should be contoured. A dose-volume histogram (DVH) is necessary to assess target coverage and evaluate the dose to normal tissues. The robustness parameters will be set at the discretion of the treating physician. Three-dimensional imaging data will serve as the basis for treatment planning and will be used to define the target volumes and organs at risk. The size, shape, and location of the target volume and its relation to the surrounding anatomy will be incorporated into the decisions regarding the specific planning method and the goals of treatment, which include the target volume coverage, normal tissue sparing, the feasibility of delivery, and the potential for verification. Target coverage and the dose to critical structures will be evaluated by examining isodose distributions and DVHs for the target volumes and normal tissue structures. Based on these evaluations, the optimal plan that maximizes conformity of treatment and minimizes the dose to normal tissues will be selected. The homogeneity of the dose will be optimized across the targeted volumes.

##### 5.7 CT Data

At the outset of this protocol, CT data will be obtained with patients in the treatment position to serve as the fundamental data set to which additional imaging data may be registered for treatment planning. The fundamental data set will include the region of the body being scanned. These regions are specified in the Appendices. The slice thickness will be no more than 2 mm throughout the volume. Intravenous contrast will not be used unless the patient is allergic to gadolinium-based contrast and cannot receive a contrasted MRI.

#### 5.8 MR Data

When necessary, MR data will be registered to CT data for planning and evaluation purposes. MR data will be situation specific but may include imaging obtained pre- and postoperatively or pre- and post-chemotherapy. Studies performed as part of the planning process will also be co-registered. Diagnostic imaging MR (DIMR) data will include 2- and/or 3-dimensionally acquired T1-weighted pre- and post-contrast imaging, along with T2-weighted, diffusion-weighted, and FLAIR sequences. Additional investigator-specified sequences will be permitted. Radiation oncology MR (ROMR) data will include imaging studies performed for planning purposes and during treatment to evaluate for changes in tissue volumes and patient positioning.

#### 5.9 Immobilization

Patients will be immobilized for radiation therapy in accordance with their level of cooperation and clinical condition, the suitability of the available devices with respect to the tumor location, and the treatment goals. Daily general anesthesia or sedation may be required in certain patients, such as very young patients, to prevent their movement during simulation and daily treatment.

#### 5.10 Simulation

Positioning: Reproducible set-ups are critical, and the use of immobilization devices is strongly encouraged. The patient may be treated in any appropriate stable position. Consideration should be given to the implications for inter- and intrafraction motion when using nonstandard position approaches.

Immobilization devices: Standard immobilization devices will be used. The methods used for localization and immobilization of the patient and tumor are critical. The imaging studies should provide a clear assessment of the target volume with the patient in the treatment position.

Special considerations: We will use range shifters or a bolus for superficial tumors (< 4 cm from skin to target) if multi-field optimization cannot produce a solution that provides adequate target coverage.

Motion management and margins to account for target volume changes: It will be important to considering the motion of normal tissues and target volumes.

##### 5.10.1 Verification Simulation

After treatment planning, the patient may need to be repositioned at the treatment isocenter (as defined by the treatment plan) for verification.

##### 5.10.2 In-Room Verification of Spatial Positioning

In-room kV–cone beam computed tomography or orthogonal kV images will be used to verify appropriate patient orientation and the relation of normal to target volumes. These variables will be chosen at the discretion of the treating physician. Image guidance will be reviewed at least weekly by the treating physician unless significant anatomic changes necessitate more frequent review. In general, soft-tissue path-length discrepancies of less than 5 mm will be tolerated.

#### 5.11 Target Volumes

##### 5.11.1 General Comments

International Commission on Radiation Units and Measurements (ICRU) Report 78 (www.icru.org) will be used to describe the prescription methods and nomenclature used for this study. Although the CT/MR images obtained immediately before radiation therapy should be used for treatment planning, the target volumes for this study will be determined by the collective information delineating the extent of disease before and after surgical resection. Most patients will require a combination of pre- and postoperative or pre- and post-chemotherapy CT/MR sequences to delineate the initial and resultant extent of disease. The GTV, CTV, internal target volume (ITV), and PTV, together with normal tissues, must be outlined on all axial imaging slices in which the structures appear. The use of contrast with MR or CT imaging will be left to the discretion of the treating physician.

##### 5.11.2 Proton Definitions for GTV, ITV, PRV, CTV, and PTV

Gross tumor volume (GTV): Investigators should register the CT/MR imaging sequence that best demonstrates the tumor and/or operative bed to contour the region that they consider to define the region with the highest concentration of tumor cells.

Internal target volume (ITV): For thoracic and abdominal primary tumors, it may be useful to specify an ITV when substantial motion is anticipated, based on a 4D CT scan. The ITV is defined as the CTV surrounded by the internal margin (IM) component of the PTV and is meant to account for potential motion or changes in the CTV.

Planning organ at risk volume (PRV): The PRV includes the corresponding organ at risk (OAR) volume surrounded by a margin to compensate for motion or physiologic change in the OAR. A margin matching the PTV margin may be added to any OAR to form the PRV.

Clinical target volume (CTV): The CTV is defined as the GTV plus a margin that is meant to represent an area of possible subclinical microscopic disease and is anatomically confined, i.e., the CTV is limited to the anatomic confines of local spread specific to the anatomic region of interest. When the GTV approaches the boundary of an anatomic compartment, the CTV will extend up to and include the boundary. The CTV margin will be context- and disease site–specific and will be determined by concurrent nonprotocol treatment plan (NPTP) or disease site–specific protocol guidelines.

Planning target volume (PTV): The PTV includes a margin which is added to the CTV in three dimensions to create the PTV. It is defined geometrically rather than anatomically. The PTV has two components: the internal margin (IM) and the set-up margin (SM). The IM is meant to compensate for all movements and variations in the size and shape of the tissues contained within the CTV. The SM is meant to account for the set-up, mechanical, and dosimetric uncertainties related to daily patient positioning, treatment equipment, and software. For this study, the PTV margin should be 3 to 5 mm, depending on the body site. The use of a PTV margin of 3 mm requires documentation that image-guided radiation therapy (IGRT) methods are being used on a daily basis. For this study, IGRT is defined as 2- or 3-dimensional digital imaging positioning.

Given that the CTV is generally confined by anatomic boundaries (limited by the dura or body wall), the PTV may extend into or beyond the bone but is unlikely to extend beyond the surface of the patient. If the PTV does extend beyond the surface of the patient, it will be manually restricted so as to be confined within the body proper. The PTV margin chosen by the treating investigator requires treatment planning MR and/or diagnostic MR imaging data with an imaging section thickness no greater than the chosen PTV margin.

The PTV will not be used to determine the distal range for the individual proton beams but will be used to report the dose according to ICRU-78. The proton distal target margin will be determined for each beam.

#### 5.12 Target Dose

##### 5.12.1 Dose Definition

For proton therapy, the absorbed dose is specified in cobalt Gray equivalent units (CGE), which is the same as the ICRU 78 relative biological effectiveness–weighted dose (DRBE), using a standard RBE of 1.1 with respect to water.

##### 5.12.2 Prescribed Dose and Fractionation

The total dose and dose per fraction will be at the discretion of the investigator and will be subject to concomitant disease site–specific protocol constraints and limitations.

#### 5.13 Dose Uniformity

Dose uniformity parameters will be dictated per the concurrent therapeutic protocol or best clinical practice as a part of NPTP guidelines.

In the rare instance that a case has to be treated with photons, rather than protons, as a result of equipment failure, at least 95% of the protocol-specified dose should encompass 100% of the PTV, and no more than 10% of the PTV should receive more than 110% of the protocol dose as evaluated by DVH. In general, IMRT photon plans will be preferred. The 100% isodose should be equal to the protocol-specified dose. Methods of generating more uniform dose distributions will be encouraged.

#### 5.14 Tissue Heterogeneity

5.14.1 Calculations must take into account tissue heterogeneity and should be performed with CT-based treatment planning to generate dose distributions and treatment calculations from CT densities. Corrections for scatter or artifact will be encouraged.

#### 5.15 Interruptions, Delays, and Dose Modifications

5.15.1 There will be no planned rests or breaks from treatment, but once radiation therapy has been initiated, the treatment may be interrupted at the discretion of the investigator. Radiation therapy may be interrupted because of a change in medical status (e.g., VP shunt failure, pleural effusion, infection, or neurologic or general medical problems). Blood-product support should be instituted according to institutional/protocol guidelines. No minimum hemoglobin or platelet level is required. The reason for an interruption exceeding 3 treatment days should be recorded in the patient treatment chart and submitted with the QA documentation (refer to the Appendix for the form). There should be no modifications of dose fractionation based on the patient's age or the field size.

#### 5.16 Treatment Technique and Beam Arrangement

5.16.1 Pencil Beam Scanning: Pencil beam scanned protons (intensity-modulated proton radiation therapy or single-field uniform dose) will be used at the discretion of the treating radiation oncologist. Every attempt will be made to minimize the dose to organs at risk without compromising the coverage of the target volume.

5.16.2 Selection of Proton Beam Arrangements: The beam arrangement will be at the discretion of the treating physician or dictated by protocol-specific guidelines. Beam arrangements should also avoid overlap at the skin surface to minimize the skin dose.

5.16.3 Range Uncertainty and Beam Configuration: Although no specific beam orientation is recommended for each body site, the beam entrance location should be chosen to limit tissue heterogeneities relative to the planning CT. The beam entry to patient surface distances should be reproducible; therefore, anterior abdominal fields in which abdominal movement can vary depending on the presence of bowel gas, phase of respiration, etc. are also discouraged.

5.16.4 Distance from Patient: In general, an arrangement of beams should be chosen so as to obtain the shortest and the most homogeneous distance to the target in order to maximize robustness relative to range uncertainty.

5.16.5 Collision Detection: No automated means of collision detection is available. Whenever possible, a virtual simulation for confirming the couch, gantry, and patient location for each individual beam should be performed before the first treatment day.

5.16.6 Field Shaping: The field shaping for protons will be completed with orthogonal beam scanning magnets. Apertures may be used to further limit the lateral penumbra if their use is commissioned in the treatment planning software and this has passed the institutional quality assurance tests.

#### 5.17 Treatment Planning Procedures

Table 5. Treatment planning sequence.

| |
|---|
| <u>Treatment planning CT</u><br>Section thickness: < 2 mm<br>Volume: top of region to inferior extent of fixation device and region<br>↓<br><u>Register correlative CT/MR imaging</u><br>↓<br><u>Required contours</u><br>GTV, CTV, PTV<br>Normal tissue contours (specified based on body site)<br>↓<br><u>Radiation treatment planning</u><br>Target volume coverage<br>Normal tissue dose recommendations<br>↓<br>Final approved treatment plan |

#### 5.18 Registration Procedures and Imaging

Disease site–specific imaging will be registered to enable the delineation of target volumes to be considered. The choice of PET, MR sequence, or CT parameters will be at the discretion of the treating physician. If the concurrent disease site–specific treatment protocol specifies specific imaging, these guidelines should be given preference.

#### 5.19 Organs at Risk

The organs-at-risk guidelines in this section are only recommendations. If the recommended doses to the organs at risk are exceeded because of target volume coverage requirements or other conditions, an explanation should be included in the quality assurance documentation (refer to the Appendix). Normal tissue dose recommendations are measured in CGE (refer to the Appendix).

#### 5.20 Structure Definitions

Final DICOM-RT structure sets should be named according to the following convention: 3-letter initials_MODAYEAR.

#### 5.21 Target Volume Nomenclature:

| Structure name | Definition |
|---|---|
| GTV_YEARMODA | The YEARMODA refers to the date of the scan used to define the structure. |
| CTV_YEARMODA | CTV is defined by the disease site–specific NPTP guidelines or a concurrent institutional or cooperative group trial. |
| ITV_YEARMODA | The ITV will be defined for tumors with inherent motion, as shown by the relevant 4D imaging. |
| PTV_YEARMODA | A body site–specific margin addition to the CTV to allow for positional uncertainty. |

#### 5.22 Dose Calculations and Reporting

5.22.1 Prescribed Dose: Prescribed dose parameters will be dictated per the concurrent therapeutic protocol or best clinical practice as a part of NPTP guidelines.

5.22.2 Normal Tissue Dosimetry: The dose to the critical organs indicated should be calculated whenever they are directly included in a radiation field. A DVH must be submitted for a category of tissue called “unspecified tissue,” which is defined as tissue contained within the skin but not otherwise identified by containment within any other structure. A DVH for “unspecified tissue” shall be submitted to enable calculation of the required volumetric information.

5.22.3 “Unspecified tissue” is defined as the outer contour of the patient on the treatment planning CT data set minus the PTV.

5.22.4 Treated Volume (mL), and Irradiated Volume (mL):

5.22.4.1 The treated volume (TV) is the tissue volume that receives therapeutic dose. This information, along with the absolute volume of the PTV, may be used by the investigators to calculate the conformity indexes (CI) CI 100% and CI 95%, respectively.

5.22.4.2 The irradiated volume (IV) is the tissue volume that receives a dose that is considered significant in relation to normal tissue tolerance.

5.22.4.3 The descriptive statistics for these and other tissue volumes may be used for correlation with unusual side effects or to develop practical guidelines for future brain tumor protocols.

Table 6. Required volumetric information.

- TV95%
- TV100%
- PTV
- CTV
- GTV
- Unspecified tissue

#### 5.23 Quality Assurance Documentation

Definitions of deviations in protocol performance

| **DEVIATION** | |
|---|---|
| **Minor** | **Major** |
| Difference in prescribed or computed dose is 6%–10% of the protocol-specified dose | Difference in prescribed or computed dose is > 10% of the protocol- or prescription-specified dose |
| More than 10% of the PTV receives less than 110% of the protocol dose or 100% of the CTV receives 90%–95% of the protocol dose | 100% of the CTV receives < 90% of the protocol- or prescription-specified dose |
| **Volume** | |
| Margins for CTV or PTV are less than specified or excessively large | A portion of the GTV or potentially tumor-bearing area (CTV) is not included in the treated volume |
| *Organs at risk* | |
| Will be assessed at time of data review | Will be assessed at time of data review |

#### 5.24 Planning Priorities

Treatment planning will prioritize target volume coverage over minor deviations in target coverage. In the event that the target coverage results in a major deviation in target structures such as the brainstem, spinal cord, bowel, kidney, optic chiasm, or optic nerves, then a minor deviation in target coverage will be acceptable.

##### 5.25 Radiotherapeutic Variables and Analyses

***Radiation Dosimetry***: Differential DVHs will be calculated for each tumor volume and for normal tissue structures, including functional subunits, and exported from the planning system for integration over the total dose interval or summarized as a mean integral dose. The modeling of radiation therapy–related treatment effects and the contribution of the radiation dose and volume to the toxicity measures outlined in this protocol will require the inclusion of clinical and treatment-related factors deemed relevant to the analysis. Accordingly, we will develop models of treatment-related effects and descriptive statistics of fractional dose-volume data to assist in the assessment of treatment toxicity.

***Dose Volume Recording***: The PTV D100 and D95 will be used as measures of the radiation prescription coverage.

***Homogeneity Index***: The homogeneity index (HI) is equal to (D2−D98)/Dp, where D2 and D98 represent the doses received by volumes corresponding to 2% and 98% of the PTV and Dp is the prescribed dose.

***Variability in Patient Positioning***: Consistent with standard medical practice, portal imaging using electronic detection systems will be acquired daily during the entire course of radiation therapy to verify positioning. The variability in the patient’s positioning, measuring differences based on a comparison of CBCT data from the treatment planning system, baseline data, and on-treatment portal images, will be evaluated. There are a number of ways to verify positioning. The most common way is for the treatment staff and radiation oncologist to check daily anatomic images. Other possibilities include using internal fiducials or mutual information (anatomy assessed on a pixel-by-pixel basis) and image registration. These potentially automated methods enable both qualitative and quantitative assessments.

### 6.0 REQUIRED EVALUATIONS, TESTS, AND OBSERVATIONS

#### 6.1 Baseline Evaluations

| Event | Baseline[1] |
|---|---|
| Informed consent | X |
| History and physical examination | X |
| Disease assessment | X |
| Region-directed CTCAE | X |
| Skin assessment[3] | X |
| Imaging assessment[4] | X |
| CBC | X |
| Chemistry | X |
| Calculated creatinine clearance | X |
| Beta HCG ** | X |
| Systemic disease status asessment[5] | X |
| Local disease status assessment[6] | X |
| Data QA[2] | |
| ** All eligible female patients of child-bearing age will have a negative pregnancy test. [1]Within 2 weeks of enrollment. [2]Within 2 weeks of enrollment. [3]Refer to the Appendix. [4]According to therapeutic protocol or best clinical practice. [5]Refer to the Appendix. [6]Refer to the Appendix. | |

Selective pre-enrollment standard of care evaluations (MRI, bone age, blood tests, etc.) prior to enrollment is considered a part of the dataset and is critical to this research and the evaluation and treatment of each patient. Selective evaluations may be performed within 1 week prior to enrollment. Diagnostic imaging studies may be performed within 2 weeks prior to enrollment. However, some evaluations may be repeated for continuity of care.

All baseline clinical and toxicity assessments will be completed by the treating radiation oncologist and/or nurse practitioner. Corresponding basic laboratory and imaging evaluation will be completed as part of the normal workup for the disease site. All female patients of child-bearing age will be required to take a pregnancy test before undergoing irradiation according to departmental policy.

#### 6.2 Evaluations During and Immediately After Therapy

| Event | During and immediately after therapy | | | |
|---|---|---|---|---|
| | Week 1 | Week n | 4–6-week eval. | 6-month eval. |
| History and physical examination | | | | X |
| CTCAE | | X | X | X |
| Skin assessment[1] | | X | X | X |
| Imaging assessment[2] | | | X | X |
| CBC | | | | X |
| Chemistry Profile | | | | X |
| Calculated creatinine clearance | | | | X |
| Systemic disease status assessment[3] | | | | X |
| Tumor response review[4] | | | X | X |
| RT treatment plan parameter review[5] | X | | | |
| Data QA[6] | X | | | X |

In cases where the follow-up schedule on the treatment protocol is on a q4-month basis, the evaluation for the protocol may be considered sufficient if performed within 1 to 2 months of that time-point.
[1]Refer to the Appendix. [2]According to protocol or best clinical practice. [3]Refer to the Appendix. [4]Refer to the Appendix. [5]Refer to the Appendix.[6]Refer to the Appendix.

All follow-up clinical and toxicity assessments will be completed by the treating radiation oncologist and/or nurse practitioner. Laboratory and imaging evaluations will be dictated largely by any concurrent disease site–specific protocols. In those cases where patients are not treated according to a treatment protocol, laboratory and imaging evaluation will be completed according to the timing listed above. The composite of the above data will be reviewed by the clinical trial nurse/coordinator and presented to the physician for authentication before being entered into the CRIS database during week 1 of patient treatment and after the 6-month evaluation.

#### 6.3 Long-Term Follow-up Evaluations

| Event | Long-term follow-up | | | |
|---|---|---|---|---|
| | Year 1 & 3 | Year 2 & 4 | Year 5 | Year 10 |
| History and physical examination | X | X | X | X |
| Disease assessment[1] | X | X | X | X |
| CTCAE | X | X | X | X |
| Skin assessment[2] | X | X | X | |

| | | | | |
|---|---|---|---|---|
| Imaging assessment[3] | X | X | X | X |
| CBC with differential[4] | X | X | X | X |
| Chemistry[4] | X | X | X | X |
| Calculated creatinine clearance[4] | X | X | X | X |
| Lipid profile[4] | X | X | X | X |
| Local control event review[5] | X | | X | X |
| Data QA[6] | | | X | X |
| In cases where the follow-up schedule on the treatment protocol is on a q4-month basis, the evaluation for the protocol may be considered sufficient if performed within 1 to 2 months of that time point. [1]Refer to the Appendix. [2]Refer to the Appendix. [3]According to protocol or best clinical practice. [4]After patient transition to the ACT clinic—laboratory evaluation will be performed according to the provider. [5]See Section 7.3. [6]Refer to the Appendix. | | | | |

All follow-up clinical and toxicity assessments will be completed by the treating radiation oncologist and/or nurse practitioner. Laboratory and imaging evaluations will be dictated largely by any concurrent disease site–specific protocols. In cases where patients are not treated according to a treatment protocol, laboratory and imaging evaluation will be completed according to the timing listed above. The composite of the above data will be reviewed by the clinical trial nurse/coordinator and presented to the physician for authentication before being entered into the database during week 1 of patient treatment and after the 6-month evaluation.

**Instructions Regarding the Logistics of All Evaluations**

It is the intent of the investigators to perform all evaluations in all patients with a view to achieving the primary objectives and pursuing the exploratory aims. Participating patients will follow the standard schedule outlined in the above tables.

**Protocol Performance**

Despite our intent to adhere as closely as possible to the guidelines of this study, unforeseen circumstances may dictate postponements, cancellations, or minor deviations from the protocol guidelines. These circumstances may include intercurrent illness, logistical problems, the absence of care providers or investigators, time constraints, shortages of materials, and a lack of funding. Exceptions to the evaluation scheme will be made at the discretion of the Principal Investigator when clinical or logistical considerations preclude evaluation at the times specified in the protocol.

**Pre-Enrollment Data**

Information included in the medical record of study patients, including evaluations performed before study enrollment, should be considered part of the study data set. Pre-enrollment information is vital to accurately determine eligibility. Pre-enrollment information is also required to accurately assess the effect of a particular treatment. This information includes longitudinal modeling and toxicity attribution and is particularly informative regarding patients who were treated with surgery before enrollment. Examples include but are not limited to prior clinical factors, treatment factors, and CNS effects measures (section 1.3) and more specifically, records of allergies, medications, and medical problems; past medical and family history; review of systems; health status and health maintenance; personal and family social history; developmental history; physical examination and vital signs; prior laboratory and clinical data, including operative and pathology reports; and diagnostic information, including imaging and information pertaining to the patient's psychologic, neurologic, ophthalmologic, and audiologic status and functional performance. Such information is critical to determining the natural history of the enrolled patient's condition and how it relates to CNS effects observed at the time of enrollment and after treatment; the use of pre-enrollment information is implied in the design of this study.

**Research Data**

The exploratory data derived from this study will be collected, analyzed, and reported while the study is ongoing and with the approval of the Principal Investigator. The analyzed data will be reported and presented at institutional and scientific meetings and at invited presentations with the approval of the Principal Investigator. Some use of the protocol data and publication of analyzed data in the form of ongoing or preliminary reports is anticipated before the completion of the study; this will be done only with the approval of the Principal Investigator.

#### 7.0 EVALUATION CRITERIA

##### 7.1 Response Criteria

Response Evaluations

Disease Status Classification: Failure will be characterized according to the site targeted by radiation therapy.

- **Solid Tumor Disease Status Classification**
  - Lesion Classification
    - Lesions targeted by radiation therapy will be classified as measurable or nonmeasurable lesions. Measurable lesions designated as radiation therapy targets will be followed quantitatively.
    - Special circumstances: Bone and cystic lesions will be evaluated using different criteria, as detailed in Appendix V.
  - Lesions Response Evaluation
    - Lesions targeted by radiation therapy will be graded according to their response to radiation therapy at 6 months after therapy.

- Systemic Disease Evaluation
  - Baseline Evaluation: Each patient's systemic disease status will be classified according to the extent and status of their systemic disease, as described in Appendix IX.
  - Six-Month Evaluation: Each patient's systemic disease status will be classified according to the extent and status of their systemic disease, as described in Appendix IX.

- **Central Nervous System Disease Classification**
  - Lesion Classification
    - Lesions targeted by radiation therapy will be classified as measurable or nonmeasurable lesions. Measurable lesions designated as radiation therapy targets will be followed quantitatively.
  - Lesions Response Evaluation
    - Lesions targeted by radiation therapy will be graded according to their response to radiation therapy at 6 months after therapy by using the RANO criteria. RANO 2010 can be found in Appendix XII.
  - Systemic Disease Evaluation
    - Baseline Evaluation: Each patient's systemic disease status will be classified according to the extent and status of their systemic disease, as described in Appendix IX.
    - Six-Month Evaluation: Each patient's systemic disease status will be classified according to the extent and status of their systemic disease, as described in Appendix IX.

- **Hematologic Malignant Disease Status Classification**
  - Lesion Classification
    - In many cases, a discrete mass may not be present at the time of radiation therapy, as is the case with many patients preparing for transplant. In these cases, target lesions will be designated as nonmeasurable lesions. Measurable lesions designated as radiation therapy targets will be followed quantitatively.

- Lesions Response Evaluation

    - Measurable target lesions targeted by radiation therapy will be graded according to their response to radiation therapy at 6 months after therapy and will be classified according to RECIST criteria. Nonmeasurable target lesions will not have a response graded.

- Systemic Disease Evaluation

    - Baseline Evaluation: For patients with lymphoma, their systemic disease status will be classified according to their stage and risk. Patients with leukemia will not have a systemic disease evaluation noted.

    - Six-Month Evaluation: Each patient's systemic disease status (not extent) will be classified according to Appendix IX.

#### 7.2 Toxicity Evaluation Criteria

CTCAE v4.0 will be used for region-directed toxicity reporting. Copies of the CTCAE versions can be downloaded from the CTEP homepage (https://ctep.cancer.gov/) or from the St. Jude DocuShare homepage. Toxicity criteria to be used in the study will be available as an appendix to the protocol. Toxicity information should be logged as a part of the medical record in Milli/Powerchart or CRIS for later data extraction. Other items not recorded in Milli/Powerchart or CRIS will be entered at a later date.

#### 7.3 Therapy Site/Progression Criteria

Analysis of Failure: There are no accepted guidelines for classifying treatment failure with respect to the targeted volumes. Classically, the terms central, in-field, marginal, or out-of-field have been used [96]. Failures that can be unequivocally classified as "central" or "in-field" are those that occur entirely within the 95% and 80%– 95% isodose lines, respectively. Failures with 95% of the recurrent tumor within the 20%–80% isodose line are classified as marginal, whereas those with 95% of the tumor volume beyond the 20% isodose line are classified as distant.

The remaining failures may be classified according to their possible origin, their geometric centroid, or the percentage of the specific targeted volumes that they occupy. The imaging studies and sequences demonstrating tumor progression or treatment failure will be registered to the imaging studies used to plan treatment, and the DVHs for the failure volume will be computed and reported.

#### 7.4 Necrosis Evaluation Criteria

Radiation necrosis secondary to photon therapy has been secondary to dose/fractionation, volume, surgical injury, or concurrent/adjuvant therapies. Regions at risk for necrosis include both grey and white matter, although white matter appears to be more prone to necrosis, and this may lead to more long-term changes and an increased incidence of symptomatic presentation. Because the risk of radiation necrosis with proton therapy is poorly documented, we will document the cumulative incidence and grade of radiation necrosis across all patients receiving proton therapy by using CTCAE v4.0. Cases of radiation necrosis should be reviewed by two independent investigators to confirm the presence or absence of necrosis as well as its grade.

#### 7.5 Vasculopathy Grading

Vasculopathy is common among pediatric patients with brain tumors and is responsible for some of the devastating effects observed after radiation therapy. The incidence, time to onset and other factors predictive of severe and life-threatening vasculopathy have not been studied systematically [97, 98]. Perioperative vasospasm and ischemia have been attributed to surgery, whereas late events are largely attributable to the radiation dose and irradiated volume. Vasculopathy is often difficult to manage, because medical or surgical intervention is instituted or considered only after the process has become established. We seek to use existing imaging protocols and grading systems to characterize the incidence of vasculopathy in the pediatric brain tumor population treated with proton therapy. MRI performed at 1.5 T or 3 T can be used to enhance resolution for better visualization of small vessels and pathologic processes, or it can be used to improve the signal-to-noise ratio to enable more sensitive diffusion and perfusion imaging. We will evaluate the clinical and treatment factors contributing to vasculopathy.

Vasculopathy will be graded from 3 to 5 according to NCI CTCAE v4.0 and assigned to one of two categories: asymptomatic vasculopathy with minor symptoms not requiring invasive intervention (grade < 3) or symptomatic vasculopathy with disabling, life-threatening implications requiring invasive intervention that may lead to death (grade 3 or higher).

#### 7.6 Permanent Neurologic Condition or Deficits

Patients treated with conformal proton radiation therapy will be reviewed for any permanent neurologic conditions or permanent neurologic deficits. Permanent neurologic conditions or deficits of interest for categorization will include necrosis, dystonia, MCA infarction, or brainstem infarction. Historically, the incidence of permanent neurologic deficits related to photon therapy has a cumulative incidence of 7%. The assessment of neurologic conditions or deficits will rely on the CTCAE v4.0 criteria, excluding vision loss, hearing loss, psychologic and psychiatric problems, sleep disorders, seizure episodes that do not require permanent treatment with anticonvulsants, and the use of any anticoagulant in the absence of clinical symptoms. The permanent conditions or deficits must be determined by two independent practitioners.

#### 7.7 Treatment-Related Death Classification

Patients treated with conformal proton radiation therapy who succumb to their disease will be reviewed, and their mortality will be classified according to the etiology. Coincident CTCAE-graded toxicities may be useful in classifying death. Briefly, the classification criteria set out by Alexander et al. will be used to categorize terminal events [92]. The classification schema is laid out in Appendix IV.

#### 7.8 Treatment-Related Second Cancers

Dosimetry should be used to estimate the relation of radiation therapy dose and volume to the risk of second cancers. A combination of field set-up pictures, a review of 3D dosimetry performed using vendor-neutral plan review software, and a clinical examination should be used in each assessment. If the second tumor is not located accurately within an organ, the tumor site–specific dose estimates should be considered imprecise, and additional means of estimating the radiation dose (e.g., a Monte Carlo simulation of neutron dose/exposure) should be considered. If the earlier radiation treatment was administered to the patient at an age when the organ size and position in the body were changing, the original tumor location can only be inferred by extrapolating back in time, using estimates about where the progenitor cell might have been located at the time of actual treatment. Even when doses can be well characterized, the dose distributions across the study population may not be ideal for evaluating the shape of the dose response or the relation of organ-specific doses to the ultimate second tumor location. Furthermore, correlations among the type of first cancer, the organ-specific radiation dose, and the age at exposure make it difficult to distinguish the effects of each factor on subsequent cancer risk.

#### 7.9 Tissue Analysis of Treatment-Related Second Cancers

***Subject cohort:*** Twenty-three RaHGGs and 21 de novo related HGGs have been identified as mentioned above within the St. Jude cohort. The expected total cohort of RaHGG planned for analysis is 30-40, although the final number will depend on the identification of additional samples at collaborating institutions. The Childhood Cancer Survivors Study as well as the University of Alabama have expressed strong interest in collaboration. Publically available data from de novo non-brainstem HGG and DIPG will be used in some of the below analyses to expand the reference cohort in this retrospective cohort study. The exposure in this case is the treatment with radiotherapy while the disease is the characteristic molecular signature. The definition of a characteristic molecular signature is a mutation pattern which predominates in the RaHGG group relative to the de novo HGG group. While a number of analyses have previously been completed on a small subset of patients, prior analyses on existing tissue samples will not be repeated.

***Tissue specimen de-identification:*** Formalin-fixed paraffin-embedded (FFPE) material will be processed in the St. Jude Anatomic Pathology Laboratory. Patient identifiers will only be used to ensure that we are testing the appropriate material. Subsequently, de-identification of samples will be performed by labeling processed material (i.e., unstained sections and FFPE scrolls) with a research accession number devoid of patient identifiers. Tissues from outside institutions will be de-identified at that institution and only the tumor bank identifier for the study will be used in the analysis. Germline tissue is not available in many cases, and because of the ethical issues with running extensive molecular characterization assays on germline tissue that could result in patient identification, this will not be pursued despite the potential relevance to the investigation.

***DNA extraction from FFPE material:*** Tumor DNA will be extracted from FFPE material by using the Maxwell16 FFPE Plus LEV purification kit and the Maxwell 16 instrument (Promega, Madison, WI) in accordance with the manufacturer's instructions.

***Whole-Exome Sequencing and Transcriptomic Analyses:*** Whole-Exome sequencing, and RNA-seq will be performed as previously described [99], [100]. For both WES sequencing and RNA-seq, paired-end sequencing will be performed by using the Illumina Genome Analyzer IIx or HighSeq platform with 100-bp read length. Whole-Exome sequence mapping, coverage, and quality assessment, and tier annotation for sequence mutations, prediction of deleterious effects of missense mutations, and identification of loss of heterozygosity will be performed as described previously [100]. Structural variations from WGS will be analyzed using CREST, while CICERO will be used on the RNA-seq data [101]. The reference human genome assembly NCBI Build 37.5 will be used for the mapping of samples. Mutations will be filtered for depth, known variations, strong germline activity and functional impact to define a clean list of mutations which will be queried for additional analyses.

***Mutation type assignment:*** Single nucleotide variations will be classified into 3 tiers: 1) coding synonymous, non-synonymous, splice-site, and noncoding RNA variants; 2) conserved variants and 3) variants in non-repeat–masked regions.

***Analysis of the significance of mutated genes:*** assess the significance of non-silent sequence mutations, we will use the Significantly Mutated Gene test [102].

***Recurrently mutated genes:*** Genes which are commonly altered in both the RaHGG and de novo HGG will be compared and listed in a common table.

***Mutation frequency and mutational composition analysis:*** To understand the impact of radiotherapy on the number and type of mutations, the absolute number of mutations, and frequencies of base pair substitutions in SNVs will be compared across RaHGG and de novo HGG using an ANOVA and Chi-square test respectively.

***Biological Impact of SNVs:*** To understand the relevance of radiation associated mutations, the predicted impact of somatic substitutions will be characterized in the RaHGG subgroup. Mutations will be assigned low, medium and high impact status using SNPEff software based on the type of mutation identified similar to.[103]

***Mutational Signature Analysis:*** Non-negative matrix factorization will be used to extract potential stable mutational signatures by plotting the signature stability across the average Frobenius reconstruction error [104]. We expect to describe characteristic mutation types, flanking substitutions and nucleotide neighbors which distinguish between mutation signatures. Spindle plots are planned for depiction of the similarity of signatures between RaHGG and de novo HGG. If a subset of RaHGG patients have a common cancer predisposition syndrome (Li fraumeni, etc), additional subsets will be compared. Given the difficulties with performing WGS on FFPE tissues, we will be unable to evaluate for the presence of a distinctive mutational signature within non-synonymous regions in many RaHGG cases. The RaHGG patients are expected to display a higher proportion of a particular signature relative to the de novo HGG cases.

***Methylation subtyping:*** An input of 350-500 ng of DNA will be applied to the Illumina Infinium bead chip arrays according to the manufacturer’s specifications and read by using the iScan microarray scanner (Illumina, San Diego, CA). DNA methylation data will be analyzed using the open source statistical programming language R. Files with raw data generated by the iScan microarray scanner will be read and processed by using Bioconductor’s *minfi* package, following the steps described in the Illumina GenomeStudio software (Illumina, San Diego, CA) [105]. Probe filtering will be performed as previously described [106]. To determine the subgroup affiliation of our cohort, consensus clustering of methylation profiles will be performed against a reference cohort of defined pediatric brain tumors [106], in a similar manner as has been described previously [107]. Each subject from our cohort will be combined with the reference cases for unsupervised consensus clustering as previously described by Sturm (5). Then the subgroup assignment of the corresponding samples will be resolved from the consensus matrix at K=5 [106].

***Methylation type comparison:*** The proportion of patients in the Sturm defined methylation subgroups of HGG will be compared in the RaHGG and de novo HGG cases using either the Chi-square or Fishers exact test.

***Analysis of CNA:*** CNA will be extracted from methylation array data by using Bioconductor’s *conumee* package [108]. The combined intensities of all available CpG probes will be normalized against those of control samples from normal brain tissue by using a linear-regression approach. Detection of amplification and chromosomal gains and losses will be performed by manual assessment of the respective loci for each individual profile after automatic scoring [106].

***CNA pattern comparison:*** Differences the proportion of genomic gains and losses will be compared across RaHGG and de novo HGG cases. Characteristic CNA will be identified and listed for comparison across de novo and RaHGG cases.

***Pathway analysis:*** Ingenuity variant analysis software will be used to identify recurrently mutated pathways from the final filtered list of mutations from above. Molecular and cellular function according to Kyoto Encyclopedia of Genes and Genomes (KEGG) designation will be described with respect to the frequency genetic mutations involved and the significance of the association.

***Comparison of Altered Pathways:*** The number of mutations impacting KEGG defined molecular and cellular functional groups will be compared across RaHGG and de novo HGG cases.

##### 7.10 Proton Treatment Plan Parameters

Composite therapy plans will be generated by the 4 to 6–week follow-up and collated with treatment planning parameters from the treatment plan phases. Treatment plan, delivery and target/OAR parameters are detailed in the Appendices. Treatment plan parameters will be exported from Eclipse, Mosaiq, and Hitachi equipment as necessary.

#### 8.0 REMOVAL FROM PROTOCOL THERAPY AND OFF-STUDY CRITERIA

Patients will be removed from study 10 years after the end of therapy and will undergo a therapy target site control and systemic disease status evaluation if no prior event has been logged. Participants can be taken off study under the following circumstances:

##### 8.1 Off-study criteria

8.1.1 Parent/patient request
8.1.2 Death
8.1.3 Noncompliance
8.1.4 Study evaluations are complete

#### 9.0 SAFETY AND ADVERSE EVENT REPORTING REQUIREMENTS

##### 9.1 Reporting to the IRB

In addition to the continuing review reports submitted to the IRB, the Principal Investigator is responsible for reporting all serious and unexpected adverse events that affect the safety of or risk to the study participants. Unexpected deaths that occur while a patient is receiving therapy or within 30 days after protocol therapy is discontinued, or any death occurring more than 30 days after protocol treatment that is felt to be related to the protocol treatment, are to be reported by Dr. Lucas to the St. Jude IRB office within 48 hours of his learning of the event. Serious and unexpected events are to be reported to the St. Jude IRB within 10 working days. All other events will be reported in the continuing review report.

##### 9.2 Reporting Adverse Experiences and Deaths to the St. Jude IRB

9.2.1 Only “unanticipated problems involving risks to participants or others,” referred to hereafter as “unanticipated problems,” are required to be reported to the St. Jude IRB promptly, but in no event should they be reported later than 10 working days after the investigator first learns of the unanticipated problem. Regardless of whether the event is internal or external, only adverse events that constitute unanticipated problems are reportable to the St. Jude IRB. As further described in the definition of unanticipated problems, this includes any event that in the opinion of the Principle Investigator meets the following criteria:

• Unexpected (in terms of its nature, severity, or frequency), given (1) the research procedures described in the protocol-related documents, such as the IRB-approved research protocol and informed consent document, as well as other available relevant information about the research; (2) the observed rate of occurrence (as compared to a credible baseline); and (3) the characteristics of the participant population being studied; and
• Related or possibly related to participation in the research; and
• Serious; or, if not serious, suggests that the research places participants or others at a greater risk of harm (including physical, psychologic, economic, or social harm) than was previously known or recognized.

Unrelated, expected deaths do not require reporting to the IRB. Although death is “serious,” an event must meet the other two requirements, i.e., “related or possibly related” and “unexpected/unanticipated,” in order to be considered reportable.

Deaths meeting reporting requirements are to be reported immediately to the St. Jude IRB, but in no event later than 48 hours after the investigator first learns of the death.

Events that are both serious and unexpected that occur during treatment or within 30 calendar days after treatment must be reported to the St. Jude Principle Investigator within five (5) working days.

9.2.1.1 Serious adverse event.
Any adverse event temporally associated with participation in research that meets any of the following criteria:

- Results in death;
- Is life threatening (it places the participant at immediate risk of death from the event as it occurred);
- Requires inpatient hospitalization or prolongation of existing hospitalization;
- Results in a persistent or significant disability/incapacity;
- Results in a congenital anomaly/birth defect; or
- Is cancer;
- Any other adverse event that, based on appropriate medical judgment, may jeopardize the participant’s health and may require medical or surgical intervention to prevent

one of the other outcomes listed in this definition (examples of such events include any substantial disruption of the participant's ability to conduct normal life functions; allergic bronchospasm requiring intensive treatment in the emergency room or at home; blood dyscrasias or convulsions that do not result in inpatient hospitalization; or the development of drug dependency or drug abuse), a congenital anomaly/birth defect, a secondary or concurrent cancer, a medication overdose, or any medical event that requires treatment to prevent any of the medical outcomes previously listed.

9.2.1.2 Unexpected adverse event.

- Any adverse event for which the specificity or severity is not consistent with the protocol-related documents, including the applicable investigator brochure, the IRB-approved consent form, the Investigational New Drug (IND) or Investigational Device Exemption (IDE) application, or other relevant sources of information, such as product labeling and package inserts; or if it does appear in such documents, an event in which the specificity, severity, or duration is not consistent with the risk information included therein; or

- The observed rate of occurrence is a clinically significant increase in the expected rate (based on a comparison with a credible baseline rate); or

- The occurrence is not consistent with the expected natural progression of any underlying disease, disorder, or condition of the participant(s) experiencing the adverse event and the participant's predisposing risk-factor profile for the adverse event.

For the purpose of this protocol, the following will NOT be considered unexpected adverse events:

- Radiation dermatitis, dry skin
- Pain due to radiation
- Mucositis
- Photophobia
- Nausea, vomiting, loss of appetite, fatigue
- Hair loss
- New or recurrent neurologic symptoms resulting from swelling of the brain
- Cyst enlargement
- Headache with or without medication requirements
- Seizure with or without medication requirements
- Abnormal electroencephalogram
- Vasculopathy with or without medication requirements
- Stroke or necrosis occurring after the completion of radiation therapy
- Secondary tumors occurring after the completion of radiation therapy
- Hormone deficiencies

- Events relating to hypothalamic obesity, precocious puberty, or diabetes insipidus
- Loss of hearing, vision, or neurologic function
- Permanent neurologic conditions or impairment, including dysautonomia or dystonia
- Psychiatric or psychologic disorders
- Wound infection or dehiscence from surgery
- Complications associated with surgery performed during or after proton therapy
- Death unequivocally related to disease progression; hospitalization for treatment of expected signs or symptoms of disease complications (shunt placement and revisions, cyst decompression) or progression of disease

9.2.2 Internal Events
Events experienced by a research participant enrolled at a site under the jurisdiction of the St. Jude IRB for either multicenter or single-center research projects.

9.2.3 External Events
Events experienced by participants enrolled at a site external to the jurisdiction of the St. Jude Institutional Review Board (IRB) or in a study for which St. Jude is not the coordinating center or the IRB of record.

9.2.4 Unanticipated Problems Involving Risks to Participants or Others
An unanticipated problem involving risks to participants or others is an event that was not expected to occur and that increases the degree of risk posed to research participants. Such an event, in general, meets all of the following criteria:

- Unexpected;
- Related or possibly related to participation in the research; and
- Suggests that the research places participants or others at a greater risk of harm (including physical, psychologic, economic, or social harm) than was previously known or recognized. An unanticipated problem involving risk to participants or others may exist even when actual harm does not occur to any participant.

9.2.5 Consistent with FDA and OHRP guidance on reporting unanticipated problems and adverse events to IRBs, the St. Jude IRB does not require the submission of external events, for example, IND safety reports, nor is a summary of such events/reports required; however, if an event giving rise to an IND safety report or other external event report constitutes an “unanticipated problem involving risks to participants or others” it must be reported in accordance with this policy. Correspondingly, given that this is a noninterventional trial and the study is being conducted on a FDA approved device, reporting of clinical trial results to the FDA is not required per FDAAA 801 requirements. In general, to be reportable, external events must have implications for the conduct of the study (for example, requiring a significant and usually safety-related change in the protocol and/or the informed consent form).

9.2.6 Although some adverse events will qualify as unanticipated problems involving risks to participants or others, some will not qualify as such, and there may be other unanticipated problems that go beyond the definitions of serious and/or unexpected adverse events. Examples of unanticipated problems involving risks to participants or others include the following:

- Improperly staging a participant's tumor, resulting in the participant being assigned to an incorrect arm of the research study;
- The theft of a research computer containing confidential participant information (breach of confidentiality); and
- The contamination of a study drug. Unanticipated problems will generally warrant consideration of substantive changes in the research protocol or informed consent process/document or other corrective actions in order to protect the safety, welfare, or rights of participants or others.

9.2.7 Reporting Adverse Events and Serious Adverse Events

Toxicity will be evaluated and graded using the scoring system according to the NCI Common Toxicity Criteria v4.0. Only the radiation therapy–related adverse events will be collected and scored. Adverse events will be reported in accordance with institutional policy. Both serious and unexpected adverse events (Grade 3–5) will be reported through the St. Jude TRACKS (Total Research and Knowledge System). Toxicity grading will be determined by the Principle Investigator, radiation oncologist, or neurosurgeon by referring to the pertinent patient history, physical examinations, and imaging studies. A persistent adverse event that has not resolved to baseline is kept open in the CRIS database until it resolves completely or returns to baseline, and it is closed out at the completion of the trial if unresolved. A persistent adverse event is one that extends continuously without resolution between treatment courses. A persistent adverse event is reported only once unless the grade becomes more severe in a subsequent treatment course, in which case the adverse event must be reported again with the new grade.

#### 9.3 Radiation Safety Committee

Initial approval is not required.

### 10.0 DATA COLLECTION, STUDY MONITORING, AND CONFIDENTIALITY

Case report forms will be replaced by electronic data entry into the CRIS database. All protocol-specific data and all radiation therapy–related Grade 3–5 adverse events will be entered into a secure CRIS database developed and maintained by St. Jude Clinical Research Informatics. Toxicity, treatment or management information not be logged as a part of the medical record in Milli/Powerchart or CRIS will be entered at a later date.

#### 10.1 Study Monitoring

Low risk: non-therapeutic prospective studies with a low-risk intervention or noninterventional trials with a greater than minimum risk. As this is a non-therapeutic protocol, a formal monitoring rule for toxicity is not required. Concurrent treatment for disease site specific studies will specify their own monitoring rules. Non protocol treatment patients will be treated per standard of care COG or institutional guidelines. Given the use of protons with COG or institutional therapy is not experimental, no formal monitoring rule is required.

The Principal Investigator and study team are responsible for ensuring protocol compliance. The study team will hold quarterly team meetings and review case histories or quality summaries of participants.

Source document review of eligibility and the informed consent process will be performed on 100% of St. Jude participants by the Protocol Eligibility Coordinators in the Central Protocol and Data Monitoring Office (CPDMO).

The Clinical Research Monitors will review up to 10% of the study participants annually for appropriateness of the informed consent process and eligibility. Additionally, serious adverse event (SAE) reporting in TRACKS and status on study will be reviewed for all participants. Other information may be monitored at the request of the Internal Monitoring Committee (IMC), the IRB, or another institutional administration. The monitor will generate a formal report to be shared with the Principal Investigator, the study team, and the IMC.

Protocol continuing reviews by the IRB and CT-SRC will occur at least annually. In addition, SAE reports in TRACKS will be reviewed by the IRB/ OHSP.

#### 10.2 Confidentiality

Source documents from the study that identify the study participant will be kept confidential in a secured area and in a password-protected database.

### 11.0 STATISTICAL CONSIDERATIONS

The data derived from this study will be collected, analyzed and reported while the study is ongoing with the approval of the principal investigators of respective therapeutic protocols such as SJMB12 and RT3CR etc. and DSMB.

The analyzed data will be reported and presented at institutional and scientific meetings and at invited presentations with the approval of the principal investigators of respective therapeutic protocols such as SJMB12 and RT3CR etc. and DSMB. Publication of analyzed data is anticipated prior to completion of the study in the form of ongoing or preliminary reports and will be done only with approval of the principal investigator of respective therapeutic protocols such as SJMB12 and RT3CR etc. and DSMB.

#### 11.1 Design and Analysis Plan for the Primary Objective

The expected number of patients, who will enroll at St. Jude Children's research hospital treated with proton therapy, is at least 100 per year. We plan to do a series of preliminary analyses after 300 patients enrolled and the last patient of those 300 patients has been followed up for at least 1 year, 3 years, 5 years and 10 years, respectively. Reporting of trial toxicity results will be contingent on the timeline of the enrolling disease site specific studies. Toxicity data from concurrent disease site specific studies will not be reported prior to the primary investigator presenting their findings at other meeting venues, in the form of a manuscript, or to clinicaltrials.gov. Final analysis will be done after 1000 patients have been enrolled and the last patient has been followed up for at least 10 years.

The primary study objective is to estimate the incidence of grade 3 and grade 4 non-hematologic toxicities at 1, 3, 5 and 10 years after the initiation of proton therapy. Serial assessments will document toxicity according to the current version of the CTCAE. Cumulative incidence of grade ≥3 non-hematologic toxicity by CTCAE will be estimated for the first occurrence of grade ≥3 non-hematologic toxicity, where failure will be considered as 'competing events'.

#### 11.2 Secondary and Exploratory Objectives

##### 11.2.1 (Secondary Objectives)

**Secondary Objective 11.2.1.1:** To estimate the incidence of necrosis, vasculopathy, and permanent neurologic deficits at 1, 3, 5, and 10 years after the initiation of proton therapy in children treated for brain tumor.

A series of preliminary analyses will be done after 300 patients enrolled and the last patient of those 300 patients has been followed up for at least 1 year, 3 years, 5 years and 10 years, respectively. Final results of the secondary objectives are expected to be available 10 years after the last patient of 1000 patients enrolled.

Cumulative incidence of vasculopathy with at least major symptoms (i.e., Vasculopathy grade > 2) will be estimated for the first occurrence of vasculopathy, where failure will be considered as 'competing events'. The associations between cumulative incidence of vasculopathy of major severity and imaging, demographic and clinical factors of interest such as radiation dose to the brain, vascular supply, age of diagnosis, host factors including genetics, and the grade of pre-existing vasculopathy will be investigated using Fine and Gray [109] regression models. Imaging variables may also be included in these models as time-dependent covariates. Patients who have pre-proton therapy vasculopathy with at least major severity will be excluded from the above analysis.

Similarly, cumulative incidence of necrosis will be estimated for the first occurrence of necrosis, where death, disease progression, and development of a new or secondary tumor will be considered as 'completing events'. Cumulative incidence of permanent neurologic deficits will be estimated for the first occurrence of permanent neurologic deficits, where failure will be considered as a ‘competing event’.

**Secondary Objective 11.2.1.2 & Secondary Objective 11.2.1.3:**

For secondary objective 11.2.1.2 and 11.2.1.3: Preliminary analyses will be done after 300 patients enrolled and the last patient of those 300 patients has been followed up for at least 5 years and 10 years, respectively. Final results of this secondary objectives are expected to be available 5 and 10 years after the last patient of 1000 patients enrolled.

**Secondary Objective 11.2.1.2:** To estimate the incidence of treatment-related mortality at 5 and 10 years after the initiation of proton therapy.

The cumulative incidence (with standard error) of treatment-related death will be calculated for all patients. Competing events are treatment not related death. Survival times will be calculated from the proton radiation date to the date of death. Patients who are alive without experiencing an event were censored on their last follow-up date.

**Secondary Objective 11.2.1.3:** To estimate the incidence of secondary cancers at 5 and 10 years after the initiation of proton therapy.

To estimate the probability of secondary cancer, the cumulative incidence (with standard error) will be calculated for all patients. Failure will be considered a competing event. Survival times were calculated from the proton radiation date to the date of first event. Patients who were still alive without experiencing an event were censored on their last follow-up date.

**Secondary Objective 11.2.1.4:** To estimate the incidence of fracture, and osteonecrosis at 1, 3, 5, and 10 years after the initiation of proton therapy in children treated for musculoskeletal tumors in an irradiated region directed manner.

The cumulative incidence (with standard error) of incidence of fracture and osteonecrosis will be calculated for children treated for musculoskeletal tumors. Competing events are treatment failure. Survival times will be calculated from the proton radiation date to the date of first event. Patients who are still alive without experiencing an event will be censored on their last follow-up date.

#### 11.3 Exploratory Objectives

11.3.1 To evaluate the contribution of treatment delivery and planning parameters to local control and toxicity.

Local failure is defined as progression of residual primary site tumor or the reappearance of tumor at the primary site. Cumulative incidence of local failure would be estimated with distant failure and death as competing risk events. Fine Gray regression method will be used to explore the relationship of cumulative incidence of local failure and the effect of the contribution of treatment delivery and planning parameters. Similarly, Fine Gray regression method will be used to explore the relationship of cumulative incidence of toxicity and treatment delivery and planning parameters.

11.3.2 To evaluate the mutational signature in patients with radiation associated high grade gliomas.

A detailed analysis plan is in section 7.9.

11.3.3 To evaluate the contribution of systemic therapies in perpetuating radiation associated toxicities.

The cumulative incidence of radiation associated toxicities in patients with systemic therapies will be compared with patients without systemic therapies through Gray's test.

11.3.4 To compare specific proton and photon radiotherapy associated toxicities specified in 1.2.1 and 1.2.4.

Historical photon radiotherapy associated toxicities were documented well for brain tumor patients (RT1 protocol) and sarcoma patients (RTSARC protocol). We will compare the toxicity data from proton radiotherapy with those protocol in disease specific manner. If the toxicity grading schema differs between SJPROTON1 and the prior, conversion between toxicity grading schemes will be performed. The toxicity data will described in listing and tables descriptively first and then the cumulative incidence of the toxicity data will be compared by using Gray's test between photon and proton patients. The tests will be implemented in exploratory manner and only in disease population with relatively big number of patients.

#### 12.0 OBTAINING INFORMED CONSENT

The process of obtaining informed consent will follow institutional guidelines. Informed consent will be obtained by the attending physician or their designee. After the diagnosis of cancer is confirmed and the research participant is deemed eligible, we will seek consent from the parents or guardians if the research participant is less than 18 years of age and from the research participant if he/she is 18 years of age or older. Verbal assent will be obtained from research participants aged 7 to 14 years and written assent from research participants aged at least 14 years but less than 18 years. A copy of the signed consent document will be given to the person who has signed the form. When a participant who was enrolled in the study with parental or guardian permission subsequently reaches the age of majority (18 years of age), the participant must undergo an informed consent process in order to continue participating in the study.

#### 12.1 Consent When English is Not the Primary Language

When English is not the primary language of the patient, parent, or legally authorized representative, the Social Work department will determine the need for an interpreter. This information will be documented in the participant's medical record. Either a certified interpreter or the telephone interpreter service will be used to translate the consent information. The process for obtaining an interpreter and for the appropriate use of an interpreter is outlined on the Interpreter Services, OHSP, and CPDMO websites.

#### 13.0 REFERENCES

[1] Boone ML, Lawrence JH, Connor WG, Morgado R, Hicks JA, Brown RC. Introduction to the use of protons and heavy ions in radiation therapy: historical perspective. Int J Radiat Oncol Biol Phys. 1977;3:65-9.

[2] Merchant TE. Clinical controversies: proton therapy for pediatric tumors. Semin Radiat Oncol. 2013;23:97-108.

[3] SJMB96 – Chemotherapy, Radiation Therapy, and Peripheral Stem Cell Transplantation in Treating Children with Newly Diagnosed Medulloblastoma or Supratentorial Primitive Neuroectodermal Tumor. ClinicalTrialsgov Identifier: NCT00003211.

[4] SJMB03 – Treatment of Patients with Newly Diagnosed Medulloblastoma, Supratentorial Primitive Neuroectodermal Tumor, or Atypical Teratoid Rhabdoid Tumor. ClinicalTrialsgov Identifier: NCT00085202.

[5] SJMB12 – A Clinical and Molecular Risk-Directed Therapy for Newly Diagnosed Medulloblastoma. ClinicalTrialsgov Identifier: NCT01878617.

[6] RT1 – A Study for Image-Guided Radiation Therapy in Pediatric Brain Tumors and Side Effects. ClinicalTrialsgov Identifier: NCT00187226.

[7] SJYC07 – Risk-Adapted Therapy for Young Children with Embryonal Brain Tumors, Choroid Plexus Carcinoma, High Grade Glioma or Ependymoma. ClinicalTrialsgov Identifier: NCT00602667.

[8] RTSARC – Radiation Therapy to Treat Musculoskeletal Tumors. ClinicalTrialsgov Identifier: NCT00186992.

[9] RMS13 – Treatment of Rhabdomyosarcoma with Chemotherapy, Radiation Therapy (Proton Beam), and Surgery. ClinicalTrialsgov Identifier: NCT01871766.

[10] ESFT13 – Therapeutic Trial for Patients With Ewing Sarcoma Family of Tumor and Desmoplastic Small Round Cell Tumors. ClinicalTrialsgov Identifier: NCT01946529.

[11] Merchant TE, Hua CH, Shukla H, Ying X, Nill S, Oelfke U. Proton versus photon radiotherapy for common pediatric brain tumors: comparison of models of dose characteristics and their relationship to cognitive function. Pediatr Blood Cancer. 2008;51:110-7.

[12] Krasin MJ, Constine LS, Friedman DL, Marks LB. Radiation-related treatment effects across the age spectrum: differences and similarities or what the old and young can learn from each other. Semin Radiat Oncol. 2010;20:21-9.

[13] Krasin MJ, Xiong X, Wu S, Merchant TE. The effects of external beam irradiation on the growth of flat bones in children: modeling a dose-volume effect. Int J Radiat Oncol Biol Phys. 2005;62:1458-63.

[14] Merchant TE, Kun LE, Wu S, Xiong X, Sanford RA, Boop FA. Phase II trial of conformal radiation therapy for pediatric low-grade glioma. J Clin Oncol. 2009;27:3598-604.

[15] Merchant TE, Sharma S, Xiong X, Wu S, Conklin H. Effect of cerebellum radiation dosimetry on cognitive outcomes in children with infratentorial ependymoma. Int J Radiat Oncol Biol Phys. 2014;90:547-53.

[16] Merchant TE, Conklin HM, Wu S, Lustig RH, Xiong X. Late effects of conformal radiation therapy for pediatric patients with low-grade glioma: prospective evaluation of cognitive, endocrine, and hearing deficits. J Clin Oncol. 2009;27:3691-7.

[17] Merchant TE, Schreiber JE, Wu S, Lukose R, Xiong X, Gajjar A. Critical combinations of radiation dose and volume predict intelligence quotient and academic achievement scores after craniospinal irradiation in children with medulloblastoma. Int J Radiat Oncol Biol Phys. 2014;90:554-61.

[18] Hua C, Bass JK, Khan R, Kun LE, Merchant TE. Hearing loss after radiotherapy for pediatric brain tumors: effect of cochlear dose. Int J Radiat Oncol Biol Phys. 2008;72:892-9.

[19] Merchant TE, Rose SR, Bosley C, Wu S, Xiong X, Lustig RH. Growth hormone secretion after conformal radiation therapy in pediatric patients with localized brain tumors. J Clin Oncol. 2011;29:4776-80.

[20] Krasin MJ, Wiese KM, Spunt SL, Hua CH, Daw N, Navid F, et al. Jaw dysfunction related to pterygoid and masseter muscle dosimetry after radiation therapy in children and young adults with head-and-neck sarcomas. Int J Radiat Oncol Biol Phys. 2012;82:355-60.

[21] Hua C, Hoth KA, Wu S, Kun LE, Metzger ML, Spunt SL, et al. Incidence and correlates of radiation pneumonitis in pediatric patients with partial lung irradiation. Int J Radiat Oncol Biol Phys. 2010;78:143-9.

[22] Krasin MJ, Hoth KA, Hua C, Gray JM, Wu S, Xiong X. Incidence and correlates of radiation dermatitis in children and adolescents receiving radiation therapy for the treatment of paediatric sarcomas. Clin Oncol (R Coll Radiol). 2009;21:781-5.

[23] Tinganelli W, Durante M, Hirayama R, Kramer M, Maier A, Kraft-Weyrather W, et al. Kill-painting of hypoxic tumours in charged particle therapy. Sci Rep. 2015;5:17016.

[24] Giantsoudi D, Sethi RV, Yeap BY, Eaton BR, Ebb DH, Caruso PA, et al. Incidence of CNS Injury for a Cohort of 111 Patients Treated With Proton Therapy for Medulloblastoma: LET and RBE Associations for Areas of Injury. Int J Radiat Oncol Biol Phys. 2016;95:287-96.

[25] Grassberger C, Trofimov A, Lomax A, Paganetti H. Variations in linear energy transfer within clinical proton therapy fields and the potential for biological treatment planning. Int J Radiat Oncol Biol Phys. 2011;80:1559-66.

[26] Kanai T, Kawachi K, Kumamoto Y, Ogawa H, Yamada T, Matsuzawa H, et al. Spot scanning system for proton radiotherapy. Med Phys. 1980;7:365-9.

[27] Kasper HB, Raeke L, Indelicato DJ, Symecko H, Hartsell W, Mahajan A, et al. The Pediatric Proton Consortium Registry: A Multi-institutional Collaboration in U.S. Proton Centers. Int J of Particle Therapy. 2014;1:323-33.

[28] Sethi RV, Giantsoudi D, Raiford M, Malhi I, Niemierko A, Rapalino O, et al. Patterns of failure after proton therapy in medulloblastoma; linear energy transfer distributions and relative biological effectiveness associations for relapses. Int J Radiat Oncol Biol Phys. 2014;88:655-63.

[29] Eaton BR, MacDonald SM, Yock TI, Tarbell NJ. Secondary Malignancy Risk Following Proton Radiation Therapy. Front Oncol. 2015;5:261.

[30] Gunther JR, Sato M, Chintagumpala M, Ketonen L, Jones JY, Allen PK, et al. Imaging Changes in Pediatric Intracranial Ependymoma Patients Treated With Proton Beam Radiation Therapy Compared to Intensity Modulated Radiation Therapy. Int J Radiat Oncol Biol Phys. 2015;93:54-63.

[31] Mahajan A. Normal tissue complications from low-dose proton therapy. Health Phys. 2012;103:586-9.

[32] Guan F, Bronk L, Titt U, Lin SH, Mirkovic D, Kerr MD, et al. Spatial mapping of the biologic effectiveness of scanned particle beams: towards biologically optimized particle therapy. Sci Rep. 2015;5:9850.

[33] Particle Therapy Co-Operative Group, www.ptcog.ch, accessed 10/26/2015.

[34] The National Association for Proton Therapy, www.proton-therapy.org, accessed 10/26/2015.

[35] Arndt C. Combination Chemotherapy in Treating Patients With Previously Untreated Rhabdomyosarcoma, ClinicalTrials.gov Identifier: NCT00003958. 1999.

[36] Matsuura T, Maeda K, Sutherland K, Takayanagi T, Shimizu S, Takao S, et al. Biological effect of dose distortion by fiducial markers in spot-scanning proton therapy with a limited number of fields: a simulation study. Med Phys. 2012;39:5584-91.

[37] Jensen CA, Chan MD, McCoy TP, Bourland JD, deGuzman AF, Ellis TL, et al. Cavity-directed radiosurgery as adjuvant therapy after resection of a brain metastasis. J Neurosurg. 2011;114:1585-91.

[38] Alexander S, Pole JD, Gibson P, Lee M, Hesser T, Chi SN, et al. Classification of treatment-related mortality in children with cancer: a systematic assessment. The Lancet Oncology. 2015;16:e604-e10.

[39] Kralik SF, Ho CY, Finke W, Buchsbaum JC, Haskins CP, Shih CS. Radiation Necrosis in Pediatric Patients with Brain Tumors Treated with Proton Radiotherapy. AJNR Am J Neuroradiol. 2015;36:1572-8.

[40] Sabin ND, Merchant TE, Harreld JH, Patay Z, Klimo P, Jr., Qaddoumi I, et al. Imaging changes in very young children with brain tumors treated with proton therapy and chemotherapy. AJNR Am J Neuroradiol. 2013;34:446-50.

[41] Watterson J, Simonton SC, Rorke LB, Packer RJ, Kim TH, Spiegel RH, et al. Fatal brain stem necrosis after standard posterior fossa radiation and aggressive chemotherapy for metastatic medulloblastoma. Cancer. 1993;71:4111-7.

[42] Chung CS, Yock TI, Nelson K, Xu Y, Keating NL, Tarbell NJ. Incidence of second malignancies among patients treated with proton versus photon radiation. Int J Radiat Oncol Biol Phys. 2013;87:46-52.

[43] Trotti A, Colevas AD, Setser A, Rusch V, Jaques D, Budach V, et al. CTCAE v3.0: development of a comprehensive grading system for the adverse effects of cancer treatment. Semin Radiat Oncol. 2003;13:176-81.

[44] Denis F, Garaud P, Bardet E, Alfonsi M, Sire C, Germain T, et al. Late toxicity results of the GORTEC 94-01 randomized trial comparing radiotherapy with concomitant radiochemotherapy for advanced-stage oropharynx carcinoma: comparison of LENT/SOMA, RTOG/EORTC, and NCI-CTC scoring systems. Int J Radiat Oncol Biol Phys. 2003;55:93-8.

[45] Rubin P, Constine LS, Fajardo LF, Phillips TL, Wasserman TH. Overview: Late efectives of normal tissues (LENT) scoring system. Int J Radiat Oncol Biol Phys. 1995;31:1041-2.

[46] LENT IV Late Effects Workshop. April 13-16, 2002. St. Petersburg, FL.

[47] Dische S, Warburton MF, Jones D, Lartigau E. The recording of morbidity related to radiotherapy. Radiother Oncol. 1989;16:103-8.

[48] Annibali R, Pisacreta M. Prevention of ischemic stroke after TIA: medical or surgical treatment? A review of the main trials. Panminerva Med. 1989;31:57-70.

[49] Murphy ES, Merchant TE, Wu S, Xiong X, Lukose R, Wright KD, et al. Necrosis after craniospinal irradiation: results from a prospective series of children with central nervous system embryonal tumors. Int J Radiat Oncol Biol Phys. 2012;83:e655-60.

[50] Ness KK, Mertens AC, Hudson MM, Wall MM, Leisenring WM, Oeffinger KC, et al. Limitations on physical performance and daily activities among long-term survivors of childhood cancer. Ann Intern Med. 2005;143:639-47.

[51] Neglia JP, Friedman DL, Yasui Y, Mertens AC, Hammond S, Stovall M, et al. Second malignant neoplasms in five-year survivors of childhood cancer: childhood cancer survivor study. J Natl Cancer Inst. 2001;93:618-29.

[52] Bowers DC, Liu Y, Leisenring W, McNeil E, Stovall M, Gurney JG, et al. Late-occurring stroke among long-term survivors of childhood leukemia and brain tumors: a report from the Childhood Cancer Survivor Study. J Clin Oncol. 2006;24:5277-82.

[53] Bowers DC, Mulne AF, Reisch JS, Elterman RD, Munoz L, Booth T, et al. Nonperioperative strokes in children with central nervous system tumors. Cancer. 2002;94:1094-101.

[54] Reulen RC, Winter DL, Frobisher C, Lancashire ER, Stiller CA, Jenney ME, et al. Long-term cause-specific mortality among survivors of childhood cancer. Jama. 2010;304:172-9.

[55] Fager M, Toma-Dasu I, Kirk M, Dolney D, Diffenderfer ES, Vapiwala N, et al. Linear energy transfer painting with proton therapy: a means of reducing radiation doses with equivalent clinical effectiveness. Int J Radiat Oncol Biol Phys. 2015;91:1057-64.

[56] Harrod-Kim P, Kadkhodayan Y, Derdeyn CP, Cross DT, 3rd, Moran CJ. Outcomes of carotid angioplasty and stenting for radiation-associated stenosis. AJNR Am J Neuroradiol. 2005;26:1781-8.

[57] Price RA, Birdwell DA. The central nervous system in childhood leukemia. III. Mineralizing microangiopathy and dystrophic calcification. Cancer. 1978;42:717-28.

[58] Yu SC, Zou WX, Soo YO, Wang L, Hui JW, Chan AY, et al. Evaluation of carotid angioplasty and stenting for radiation-induced carotid stenosis. Stroke; a journal of cerebral circulation. 2014;45:1402-7.

[59] Indelicato DJ, Flampouri S, Rotondo RL, Bradley JA, Morris CG, Aldana PR, et al. Incidence and dosimetric parameters of pediatric brainstem toxicity following proton therapy. Acta oncologica. 2014;53:1298-304.

[60] Carangelo B, Cerillo A, Mariottini A, Peri G, Rubino G, Mourmouras V, et al. Therapeutic strategy of late cerebral radionecrosis. A retrospective study of 21 cases. Journal of neurosurgical sciences. 2010;54:21-8.

[61] Drezner N, Hardy KK, Wells E, Vezina G, Ho CY, Packer RJ, et al. Treatment of pediatric cerebral radiation necrosis: a systematic review. J Neurooncol. 2016;130:141-8.

[62] Chen JR, Xu HZ, Ding JB, Qin ZY. Radiotherapy after hyperbaric oxygenation in malignant gliomas. Current medical research and opinion. 2015;31:1977-84.

[63] Glover M, Smerdon GR, Andreyev HJ, Benton BE, Bothma P, Firth O, et al. Hyperbaric oxygen for patients with chronic bowel dysfunction after pelvic radiotherapy (HOT2): a randomised, double-blind, sham-controlled phase 3 trial. The Lancet Oncology. 2016;17:224-33.

[64] Gajjar A, Pfister SM, Taylor MD, Gilbertson RJ. Molecular insights into pediatric brain tumors have the potential to transform therapy. Clin Cancer Res. 2014;20:5630-40.

[65] Armstrong GT, Liu Q, Yasui Y, Huang S, Ness KK, Leisenring W, et al. Long-term outcomes among adult survivors of childhood central nervous system malignancies in the Childhood Cancer Survivor Study. J Natl Cancer Inst. 2009;101:946-58.

[66] Armstrong GT, Liu Q, Yasui Y, Neglia JP, Leisenring W, Robison LL, et al. Late mortality among 5-year survivors of childhood cancer: a summary from the Childhood Cancer Survivor Study. J Clin Oncol. 2009;27:2328-38.

[67] Ladra MM, Wang KK, Terezakis SA. Pencil-beam scanning for pediatric rhabdomyosarcoma: Promise and precautions. Pediatr Blood Cancer. 2016;63:1698-9.

[68] Ladra MM, Edgington SK, Mahajan A, Grosshans D, Szymonifka J, Khan F, et al. A dosimetric comparison of proton and intensity modulated radiation therapy in pediatric rhabdomyosarcoma patients enrolled on a prospective phase II proton study. Radiother Oncol. 2014;113:77-83.

[69] Moteabbed M, Yock TI, Depauw N, Madden TM, Kooy HM, Paganetti H. Impact of Spot Size and Beam-Shaping Devices on the Treatment Plan Quality for Pencil Beam Scanning Proton Therapy. Int J Radiat Oncol Biol Phys. 2016;95:190-8.

[70] Kamran SC, Berrington de Gonzalez A, Ng A, Haas-Kogan D, Viswanathan AN. Therapeutic radiation and the potential risk of second malignancies. Cancer. 2016;122:1809-21.

[71] Setlow RB, Swenson PA, Carrier WL. Thymine Dimers and Inhibition of DNA Synthesis by Ultraviolet Irradiation of Cells. Science. 1963;142:1464-6.

[72] Sjoblom T, Jones S, Wood LD, Parsons DW, Lin J, Barber TD, et al. The consensus coding sequences of human breast and colorectal cancers. Science. 2006;314:268-74.

[73] Sherborne AL, Davidson PR, Yu K, Nakamura AO, Rashid M, Nakamura JL. Mutational Analysis of Ionizing Radiation Induced Neoplasms. Cell Rep. 2015;12:1915-26.

[74] Galaburda AM, Holinger DP, Bellugi U, Sherman GF. Williams syndrome: neuronal size and neuronal-packing density in primary visual cortex. Archives of neurology. 2002;59:1461-7.

[75] Tarkkanen M, Wiklund TA, Virolainen MJ, Larramendy ML, Mandahl N, Mertens F, et al. Comparative genomic hybridization of postirradiation sarcomas. Cancer. 2001;92:1992-8.

[76] Nakashima M, Takamura N, Namba H, Saenko V, Meirmanov S, Matsumoto N, et al. RET oncogene amplification in thyroid cancer: correlations with radiation-associated and high-grade malignancy. Human pathology. 2007;38:621-8.

[77] Paugh BS, Qu C, Jones C, Liu Z, Adamowicz-Brice M, Zhang J, et al. Integrated molecular genetic profiling of pediatric high-grade gliomas reveals key differences with the adult disease. J Clin Oncol. 2010;28:3061-8.

[78] Liu X, Yan Y, Li F, Zhang D. Fruit and vegetable consumption and the risk of depression: A meta-analysis. Nutrition. 2016;32:296-302.

[79] Craft AW. Childhood cancer--mainly curable so where next? Acta paediatrica. 2000;89:386-92.

[80] Armenian SH, Hudson MM, Mulder RL, Chen MH, Constine LS, Dwyer M, et al. Recommendations for cardiomyopathy surveillance for survivors of childhood cancer: a report from the International Late Effects of Childhood Cancer Guideline Harmonization Group. The Lancet Oncology. 2015;16:e123-36.

[81] Braam KI, van der Torre P, Takken T, Veening MA, van Dulmen-den Broeder E, Kaspers GJ. Physical exercise training interventions for children and young adults during and after treatment for childhood cancer. Cochrane Database Syst Rev. 2016;3:CD008796.

[82] Cox CL, Zhu L, Ojha RP, Li C, Srivastava DK, Riley BB, et al. The unmet emotional, care/support, and informational needs of adult survivors of pediatric malignancies. Journal of cancer survivorship : research and practice. 2016;10:743-58.

[83] Wang KW, Valencia M, Banfield L, Chau R, Fleming A, Singh SK, et al. The effectiveness of interventions to treat obesity in survivors of childhood brain tumors: a systematic review protocol. Systematic reviews. 2016;5:101.

[84] Armstrong GT, Chen Y, Yasui Y, Leisenring W, Gibson TM, Mertens AC, et al. Reduction in Late Mortality among 5-Year Survivors of Childhood Cancer. N Engl J Med. 2016;374:833-42.

[85] Berg CJ, Stratton E, Esiashvili N, Mertens A. Young Adult Cancer Survivors' Experience with Cancer Treatment and Follow-Up Care and Perceptions of Barriers to Engaging in Recommended Care. Journal of cancer education : the official journal of the American Association for Cancer Education. 2016;31:430-42.

[86] Green DM, Kun LE, Matthay KK, Meadows AT, Meyer WH, Meyers PA, et al. Relevance of historical therapeutic approaches to the contemporary treatment of pediatric solid tumors. Pediatr Blood Cancer. 2013;60:1083-94.

[87] Hudson MM, Neglia JP, Woods WG, Sandlund JT, Pui CH, Kun LE, et al. Lessons from the past: opportunities to improve childhood cancer survivor care through outcomes investigations of historical therapeutic approaches for pediatric hematological malignancies. Pediatr Blood Cancer. 2012;58:334-43.

[88] Mauz-Korholz C, Metzger ML, Kelly KM, Schwartz CL, Castellanos ME, Dieckmann K, et al. Pediatric Hodgkin Lymphoma. J Clin Oncol. 2015;33:2975-85.

[89] Bartels U. Chemotherapy Followed by Radiation Therapy in Treating Younger Patients With Newly Diagnosed Localized Central Nervous System Germ Cell Tumors, ClinicalTrials.gov Identifier: NCT01602666. 2012.

[90] Packer RJ, Gajjar A, Vezina G, Rorke-Adams L, Burger PC, Robertson PL, et al. Phase III study of craniospinal radiation therapy followed by adjuvant chemotherapy for newly diagnosed average-risk medulloblastoma. J Clin Oncol. 2006;24:4202-8.

[91] Merchant TE, Hodgson D, Laack NN, Wolden S, Indelicato DJ, Kalapurakal JA, et al. Children's Oncology Group's 2013 blueprint for research: radiation oncology. Pediatr Blood Cancer. 2013;60:1037-43.

[92] Alexander S, Pole JD, Gibson P, Lee M, Hesser T, Chi SN, et al. Classification of treatment-related mortality in children with cancer: a systematic assessment. The Lancet Oncology. 2015;16:e604-10.

[93] Hwang UJ, Shin DH, Kim TH, Moon SH, Lim YK, Jeong H, et al. The effect of a contrast agent on proton beam range in radiotherapy planning using computed tomography for patients with locoregionally advanced lung cancer. Int J Radiat Oncol Biol Phys. 2011;81:e317-24.

[94] Hallemeier C. Pencil Beam Scanning Proton Radiotherapy for Esophageal Cancer, ClinicalTrials.gov Identifier: NCT02452021. https://clinicaltrials.gov/ct2/show/NCT02452021?term=NCT02452021&rank=1. 2015.

[95] Prescribing, Recording, and Reporting Proton Beam Therapy. ICRU Report 62 (Supplement to ICRU Report 50). International Commission on Radiation units and measurements. Washington, D.C.: ICRU Report; 1999.

[96] McDonald MW, Shu HK, Curran WJ, Jr., Crocker IR. Pattern of failure after limited margin radiotherapy and temozolomide for glioblastoma. Int J Radiat Oncol Biol Phys. 2011;79:130-6.

[97] Murphy ES, Xie H, Merchant TE, Yu JS, Chao ST, Suh JH. Review of cranial radiotherapy-induced vasculopathy. J Neurooncol. 2015;122:421-9.

[98] Sutton LN. Vascular complications of surgery for craniopharyngioma and hypothalamic glioma. Pediatr Neurosurg. 1994;21 Suppl 1:124-8.

[99] Zhang J, Benavente CA, McEvoy J, Flores-Otero J, Ding L, Chen X, et al. A novel retinoblastoma therapy from genomic and epigenetic analyses. Nature. 2012;481:329-34.

[100] Zhang J, Ding L, Holmfeldt L, Wu G, Heatley SL, Payne-Turner D, et al. The genetic basis of early T-cell precursor acute lymphoblastic leukaemia. Nature. 2012;481:157-63.

[101] Wang J, Mullighan CG, Easton J, Roberts S, Heatley SL, Ma J, et al. CREST maps somatic structural variation in cancer genomes with base-pair resolution. Nat Methods. 2011;8:652-4.

[102] Dees ND, Zhang Q, Kandoth C, Wendl MC, Schierding W, Koboldt DC, et al. MuSiC: identifying mutational significance in cancer genomes. Genome Res. 2012;22:1589-98.

[103] Cingolani P, Patel VM, Coon M, Nguyen T, Land SJ, Ruden DM, et al. Using Drosophila melanogaster as a Model for Genotoxic Chemical Mutational Studies with a New Program, SnpSift. Front Genet. 2012;3:35.

[104] Gao Y, Church G. Improving molecular cancer class discovery through sparse non-negative matrix factorization. Bioinformatics. 2005;21:3970-5.

[105] Aryee MJ, Jaffe AE, Corrada-Bravo H, Ladd-Acosta C, Feinberg AP, Hansen KD, et al. Minfi: a flexible and comprehensive Bioconductor package for the analysis of Infinium DNA methylation microarrays. Bioinformatics. 2014;30:1363-9.

[106] Sturm D, Witt H, Hovestadt V, Khuong-Quang DA, Jones DT, Konermann C, et al. Hotspot mutations in H3F3A and IDH1 define distinct epigenetic and biological subgroups of glioblastoma. Cancer Cell. 2012;22:425-37.

[107] Broniscer A, Chamdine O, Hwang S, Lin T, Pounds S, Onar-Thomas A, et al. Gliomatosis cerebri in children shares molecular characteristics with other pediatric gliomas. Acta Neuropathol. 2016;131:299-307.

[108] Hovestadt V, Zapatka M. Enhanced copy-number variation analysis using Illumina 450k methylation arrays. In: Bioconductor: open source software for bioinformatics. R package version 0.99.4. *conumee*. 2015.

[109] Fine JP, Gray RJ. A proportional hazards model for the subdistribution of a competing risk. J Am Stat Assoc. 1999:496-509.

### APPENDICES

#### APPENDIX I: SCHEDULE OF EVALUATIONS

| Event | Baseline[1] | Early phase | | | | Late phase | | | |
|---|---|---|---|---|---|---|---|---|---|
| | | **Week 1** | **Week n** | **4–6-week eval.** | **6-month eval.** | **Year 1, 3 eval.** | **Year 2, 4 eval.** | **Year 5 eval.** | **Year 10 eval.** |
| Informed consent | X | | | | | | | | |
| History and physical examination[3] | X | | | | X | X | X | X | X |
| Disease assessment[5] | X | | | | X | X | X | X | X |
| CTCAE | X | | X | X | X | X | X | X | X |
| Skin assessment[3] | X | | X | X | X | X | | X | |
| Imaging assessment[4] | X | | | X | X | X | X | X | X |
| Hematology[6] | X | | | | X | X | X | X | X |
| Chemistries[6] | X | | | | X | X | X | X | X |
| Calculated creatinine clearance[6] | X | | | | X | X | X | X | X |
| Lipid profile[6] | X | | | | X | X | X | X | X |
| Urinalysis[6] | X | | | | X | X | X | X | X |
| Beta HCG[6] | X | | | | | | | | |
| Clinical event review[5] | | | | | X | X | | X | X |
| Systematic Disease Status Assessment[5] | | | | | | | | | |
| Local Disease Status Assessment[5] | | | | | | | | | |
| Tumor Response Review[5] | | | | | | | | | |
| RT treatment plan parameter review[2] | | | | | | | | | |
| Data QA | | X | | | X | | | X | X |

** All eligible female patients of child-bearing age will have a negative pregnancy test. [1]Within 2 weeks of enrollment. [2]Refer to the Appendix. [3]Refer to the Appendix. [4]According to therapeutic protocol or best clinical practice. [5]Refer to the Appendix. [6]Refer to the Appendix.

#### APPENDIX II: Early Phase QA

#### CHECKLIST

| **Clinical information** | | **Toxicity evaluation** |
|---|---|---|
| Baseline & 6-month history & physical examination<br><br>Disease assessment<br><br>Treatment information | | CTCAE & RTOG: baseline, week n, 4–6-week evaluation, 6-month evaluation<br><br>PRO-CTCAE evaluation: baseline, 4–6-week evaluation, 6-month evaluation<br><br>Skin evaluation: baseline, week n, 4–6-week evaluation, 6-month evaluation |
| **Laboratory & pathology assessment** | | **Imaging assessment** |
| Hematology, chemistries, creatinine clearance, lipid profile, urinalysis<br><br>G4K: germline & tumor samples | | Region-directed MRI: baseline, 4–6-week evaluation, 6-month evaluation<br><br>Tumor-directed MRI: baseline, 4–6-week evaluation, 6-month evaluation |
| **Radiation therapy treatment plan** | | **Radiation therapy treatment data** |
| CT planning scan<br><br>RT dose file<br><br>LET dose file<br><br>RT structure file: region-directed OARs, target | | Serial CBCT<br><br>Mosaiq beam data<br><br>Hitachi treatment data<br><br>Physics QA assessment (OSL) |

| |
|---|
| RT plan file<br><br>Disease-directed MRI |
| **Clinical event** |
| Treatment-related mortality<br><br>Alive/dead<br><br>Local control/failure |

**PROCESSING**

RT structure file: Scrubbed and standardized structures
RT dose and LET dose file: registered to CT planning scan
Imaging database: region-directed MRI aligned and registered to CT planning scan
IGRT database: CBCT aligned and registered to CT planning scan

#### APPENDIX III: Late-Phase QA

##### CHECKLIST

| Clinical information | | Toxicity evaluation |
|---|---|---|
| History & physical examination: Year 1, 2, 3, 4, 5<br><br>Disease assessment: Year 1, 2, 3, 4, 5<br><br>Subsequent treatment information: Year 1, 2, 3, 4, 5 | | CTCAE & RTOG: Year 1, 2, 3, 4, 5<br><br>PRO-CTCAE evaluation: Year 1, 2, 3, 4, 5<br><br>Skin evaluation: Year 1, 2, 3, 4, 5 |
| **Laboratory & pathology assessment** | | **Imaging assessment** |
| Hematology, chemistries, creatinine clearance, lipid profile, urinalysis: Year 1, 2, 3, 4, 5 | | Region-directed MRI: Year 1, 2, 3, 4, 5<br><br>Tumor-directed MRI: Year 1, 2, 3, 4, 5 |
| **Clinical event** | | |
| Treatment-related mortality<br><br>Alive/dead<br><br>Local control/failure | | |

##### PROCESSING

Imaging database: region-directed MRI aligned and registered to CT planning scan

#### APPENDIX IV: Treatment-Related Mortality

***Panel*: Criteria used in previous treatment-related mortality definitions in paediatric oncology**

- Absence of progressive disease at death[4,5,12–15]
- In remission at death[4,15–18]
- No history of haemopoietic stem-cell transplantation[19–21]
- No history of relapse[4,5,12–18]
- No history of induction failure[12–14]
- Death during induction therapy[16–18]
- Death before a set number of days of therapy[4,5,15]

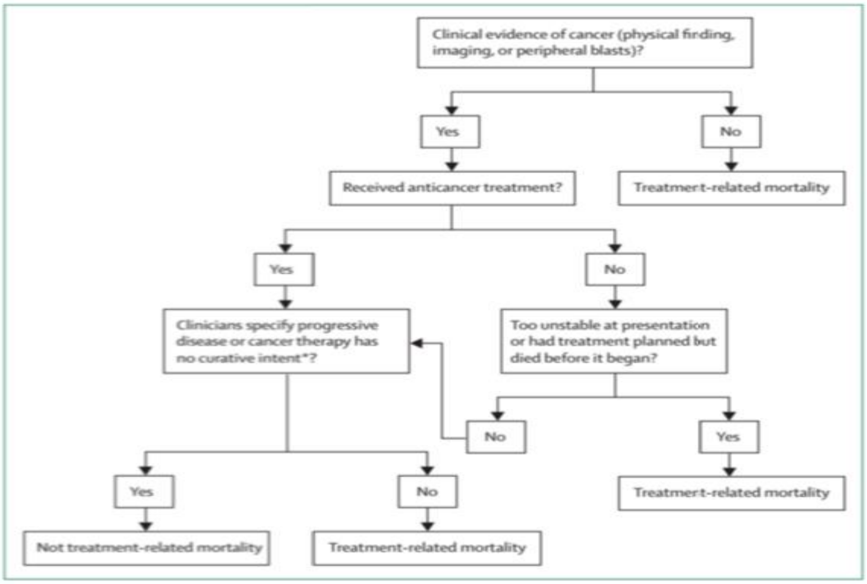

***Figure*: Classification of treatment-related mortality in children with cancer**
*Associated with key terms including refractory disease, resistant disease, non-responsive disease, palliative care, intervention not possible, poor prognosis, end-of-life care.

[92]

**APPENDIX V: Evaluation criteria for Target Lesions**

Non-measurable lesions will not be followed quantitatively for response and thus will be simply followed qualitatively. Non-measurable lesions include ascites, lymphangitic spread, masses smaller than 10 mm, lymph nodes 10 to 14 mm on the short axis, or benign findings.

Measurable lesions will be followed quantitatively and evaluated at 6 months for response.

Definition:

- Tumor ≥ 10 mm in longest diameter (LD) on an axial image on CT or MRI with ≤ 5 mm reconstruction interval
  - ―If slice thickness exceeds 5 mm, LD must be at least twice the thickness
- Tumor ≥ 20 mm LD by chest x-ray (if clearly defined and surrounded by aerated lung); CT is preferred (even without contrast)
- Tumor ≥ 10 mm LD on clinical evaluation (photo) with electronic calipers; skin photos should include a ruler
  - ―Lesions that cannot be accurately measured with calipers should be recorded as non-measurable
- Lymph nodes ≥ 15 mm on short axis on CT (CT slice thickness no more than 5 mm)
- Ultrasound cannot be used to measure lesions

Measurable lesions designated and treated as radiation therapy targets will be followed quantitatively according to RECIST.

Baseline evaluation:

Measurable target lesions will be measured at baseline before therapy based on corollary imaging from the time of simulation. Measurements should be documented in three dimensions (length, width, and height). Measurements may be gleaned from diagnostic radiology reports immediately before radiation therapy.

Six-month post-treatment evaluation:

Measurable target lesions treated with radiation therapy will be measured in three dimensions based on multimodality imaging and classified according to the following response definitions.

##### Target Lesion Evaluation

| Response | Definition |
|---|---|
| Complete Response (CR) | Disappearance of all extranodal target lesions. All pathological lymph nodes must have decreased to <10 mm in short axis. |
| Partial Response (PR) | At least a 30% decrease in the SLD of target lesions, taking as reference the baseline sum diameters |
| Progressive Disease (PD) | SLD increased by at least 20% from the smallest value on study (including baseline, if that is the smallest)<br>The SLD must also demonstrate an absolute increase of at least 5 mm.<br>(Two lesions increasing from 2 mm to 3 mm, for example, does not qualify) |
| Stable Disease (SD) | Neither sufficient shrinkage to qualify for PR nor sufficient increase to qualify for PD |

#### APPENDIX VI: CTCAE V4.0 Grading

http://evs.nci.nih.gov/ftp1/CTCAE/CTCAE_4.03_2010-06-14_QuickReference_8.5x11.pdf
http://ctep.cancer.gov/protocolDevelopment/electronic_applications/ctc.htm

**APPENDIX VII: Unplanned Treatment Changes**

Any unplanned modification or deviation from the original treatment plan stated in the physician's treatment planning note, initial consultation, or concurrent disease site–specific protocol should specify the reasons for discontinuation of treatment and provide supporting documentation. These patients will remain on study.

Acceptable reasons for discontinuation include the following:

- Disease progression. Progression leading to discontinuation could take the form of progression necessitating more aggressive management (e.g., switching from definitive chemoradiotherapy to a surgical approach), progression leading to worsening symptoms necessitating ICU admission, or other forms of intensive care, etc. Supporting evidence from imaging, pathology, or a clinical exam illustrating the means of detecting progression should be provided. If multiple supporting pieces of evidence are available, then all should be documented. For example, radiographic demonstration of DIPG progression leading to new cranial nerve deficits, etc.

- Unexpected toxicity. This is defined as any clinically significant toxicity that the patient finds intolerable. The toxicity may have a laboratory, imaging, or symptom-only component. All evidentiary components should be documented. The unexpected toxicity grade and CTCAE designation should be documented. The duration of the toxicity, as well as any modifying features or contributors (either patient or MD specified) should also be noted.

- Patient preference. A patient may simply chose to not receive further therapy.
    - Supporting documentation should specify the nature of the patient's preference. This may overlap with other reasons for discontinuation or may concern more practical matters, such as being homesick or a major family event having occurred (e.g., the death of a family member death).

- Death. The type of death should be noted and described according to the treatment related mortality section of Appendix IV.

- Change in the intent of the intervention.
    - The intent of the intervention may change due to patient preference, progression, or the appearance of toxicities coincident with therapy.
    - The reasons for a change in intent should be documented, as well as the initial and subsequent intent of the intervention.
    - Multiple specified reasons are allowed when the motivations/reasons overlap. For example, when disease progression leads to a change in the intent of the intervention from curative to palliative intent.
    - A patient may choose to forgo further therapy because of toxicity; thus, this could be recorded as both preference and toxicity. Additionally, disease progression and death could also overlap.

#### APPENDIX VIII: Systemic Disease Evaluation

##### Baseline Disease Evaluation

Extent of disease at initial diagnosis: the disease extent at the time of presentation before any and all therapy:

Localized: no evidence of spread beyond the primary tumor.

Locoregional: presence of the primary tumor and lymph nodes.

Oligo-metastatic: less than 5 lesions present, as evaluated by PET, CT C/A/P, or bone scan.

Metastatic: more than 5 lesions present or evidence of bone marrow involvement.

Extent of disease at radiation therapy: The disease extent at the time of radiotherapy will be classified according to the following criteria:

Localized: no evidence of spread beyond the primary tumor.

Locoregional: presence of the primary tumor and lymph nodes.

Oligo-metastatic: Less than 5 lesions present, as evaluated by PET, CT C/A/P, or bone scan.

Metastatic: more than 5 lesions present or evidence of bone marrow involvement.

Status at time of radiation therapy:

No evidence of disease: the absence of any radiographic disease.

Stable disease: the presence of radiographic disease with no progression over the last 3 months.

Local progression: see definition in Appendix V.

Systemic progression: local progression as defined above coincident with progression in the last 3 months as more than 1 metastatic lesion, the development of new areas of radiographic disease, or the cytopathologic demonstration of disease within a body compartment (CSF, pleural, ascitic fluid) not previously documented.

**Six-Month Disease Evaluation**

Extent of disease at 6 months: The disease extent at 6 months will be classified according to the following criteria:

Localized: no evidence of spread beyond the primary tumor.

Locoregional: the presence of the primary tumor and lymph nodes.

Oligo-metastatic: more than 5 lesions present, as evaluated by PET, CT C/A/P, or bone scan.

Metastatic: more than 5 lesions present or evidence of bone marrow involvement.

Status at 6 months:

No evidence of disease: the absence of any radiographic disease.

Stable disease: the presence of radiographic disease with no progression over the last 6 months.

Local progression: see definition in Appendix V.

Systemic progression: progression in the last 6 months defined as 1 or more new metastatic lesions, or the cytopathologic demonstration of disease within a body compartment (CSF, pleural, ascitic fluid) not previously documented.

Local and systemic progression: both local and systemic progression as defined above.

#### APPENDIX IV: CNS Systemic Disease Evaluation

##### Baseline Disease Evaluation

Extent of disease at initial diagnosis: the disease extent at the time of presentation before any and all therapy:

Intracranial localized: no evidence of noncontiguous spread beyond the primary tumor.

Spine localized: no evidence of noncontiguous spread beyond the primary tumor.

Intracranial metastatic: primary tumor of intracranial origin with leptomeningeal disease or noncontiguous spread in the brain parenchyma.

Spinal metastatic: primary tumor of intraspinal origin with leptomeningeal disease, positive cytology, or noncontiguous spread along the spinal cord.

Intracranial and spinal metastatic disease: the presence of intracranial and spinal metastatic spread.

Extra-CNS disease: positive bone marrow or non-contiguous extra-CNS disease.

Extent of disease at radiation therapy: The disease extent at the time of radiation therapy will be classified according to the following criteria:

Intracranial localized: no evidence of noncontiguous spread beyond the primary tumor.

Spine localized: no evidence of noncontiguous spread beyond the primary tumor.

Intracranial metastatic: primary tumor of intracranial origin with leptomeningeal disease or noncontiguous spread in the brain parenchyma.

Spinal metastatic: primary tumor of intraspinal origin with leptomeningeal disease, positive cytology, or noncontiguous spread along the spinal cord.

Intracranial and spinal metastatic disease: the presence of intracranial and spinal metastatic spread.

Extra-CNS disease: positive bone marrow or non-contiguous extra-CNS disease.

Status at time of radiation therapy:

No evidence of disease: the absence of any radiographic disease.

Stable disease: the presence of radiographic disease without progression over the last 3 months.

Local progression: See definition in Appendix V.

Systemic progression: local progression as defined above coincident with progression in the last 3 months as more than 1 metastatic lesion, the development of new areas of radiographic disease, or the cytopathologic demonstration of disease within a body compartment (CSF, pleural, ascitic fluid) not previously documented.

**Six-Month Disease Evaluation**

Extent of disease at 6 months: The disease extent at 6 months will be classified according to the following criteria:

Intracranial localized: no evidence of noncontiguous spread beyond the primary tumor.

Spine localized: no evidence of noncontiguous spread beyond the primary tumor.

Intracranial metastatic: primary tumor of intracranial origin with leptomeningeal disease or noncontiguous spread in the brain parenchyma.

Spinal metastatic: Primary tumor of intraspinal origin with leptomeningeal disease, positive cytology, or noncontiguous spread along the spinal cord.

Intracranial and spinal metastatic disease: the presence of intracranial and spinal metastatic spread.

Extra-CNS disease: positive bone marrow or non-contiguous extra-CNS disease

Status at 6 months:

No evidence of disease: the absence of any radiographic disease.

Stable disease: The presence of radiographic disease with no progression over the last 6 months.

Local progression: See definition in Appendix V.

Systemic progression: local progression as defined above coincident with progression in the last 6 months as more than 1 metastatic lesion, the development of new areas of radiographic disease, or the cytopathologic demonstration of disease within a body compartment (CSF, pleural, ascitic fluid) not previously documented.

**APPENDIX X: Body Regions**

Brain/head & neck patients: from the top of the indexing device (i.e., the indexing bar, wing board, zak 2 board [proton fixation device]) through the sternal notch/top of the shoulders.

Chest: from the bottom of patients chin through the top of the iliac crests.

Abdomen/pelvis: from the top of diaphragm through the mid-femur.

Lower extremity: the entire immobilization device through the mid-femur.

Upper Extremity: the entire extremity, as well as the entire fixation device. Include any anatomy to be avoided, such as the patient's head.

CT parameters:

- Scans should be completed with a slice thickness of 1.0 mm to 1.5 mm for most simulations.

- In cases where the entire body is scanned, slices less than or equal to 2 mm should be used.

- When contrast is used, one scan with contrast and an identical scan without contrast should be acquired.

**APPENDIX XI: RANO Criteria**

https://www.iconplc.com/icon-files/docs/IMI%20Brochures/IMI_RANO_Critieria_Booklet.pdf

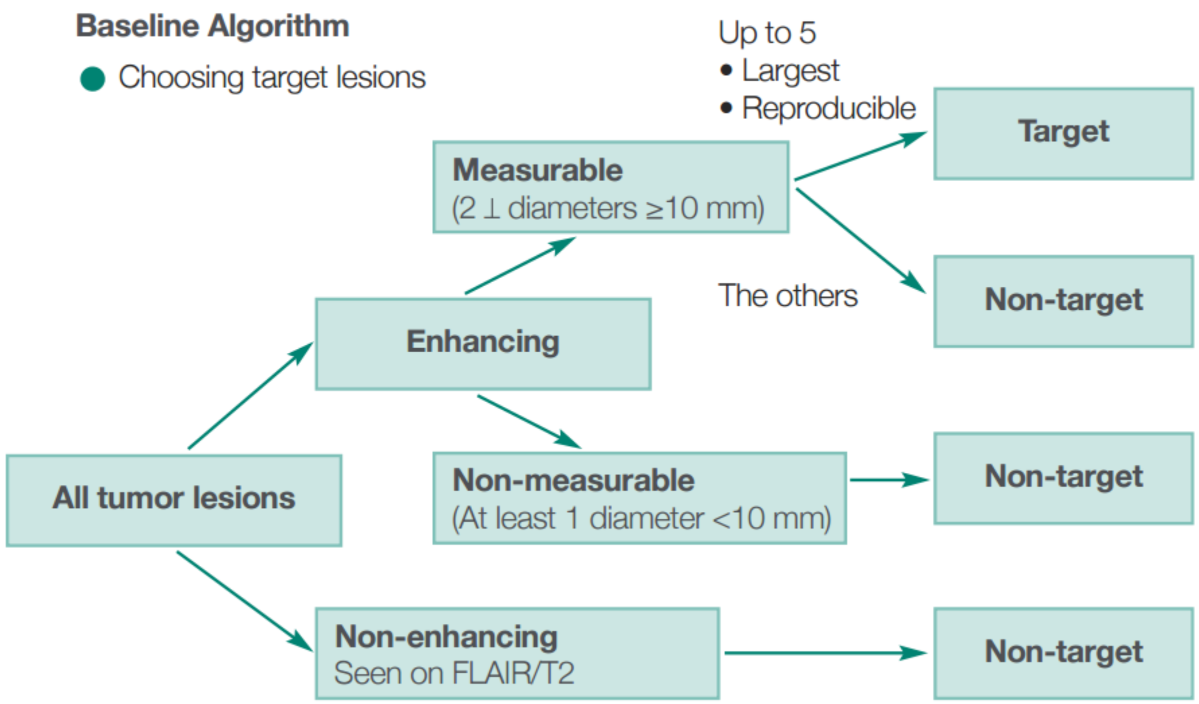

**Target Lesions**

- Calculate products of maximal diameters and add them together to yield the sum of products of diameters (SPD)

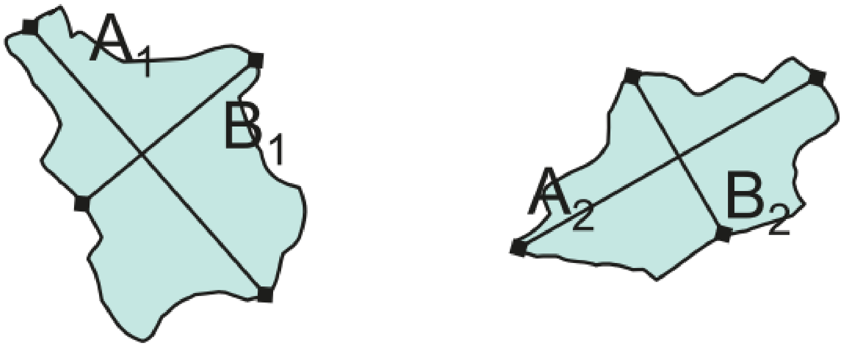

$$A_1 \times B_1 + A_2 \times B_2 + \ldots = SPD$$

**Target Lesion Response Definitions**

| Response | Definition |
|---|---|
| Complete Response (CR) | All target lesions have disappeared (look out for pseudoresponse†) |
| Partial Response (PR) | SPD decreased by ≥50% from baseline value (look out for pseudoresponse†) |
| Stable Disease (SD) | SPD <50% decrease to <25% increase |
| Progressive Disease (PD) | SPD increased by ≥25% from nadir value (look out for pseudoprogression‡) |
| Unable to Assess (UA) | Some target lesions cannot be evaluated because of technical factors |

† - CR and PR have to be confirmed ≥4 wks later. If not confirmed, response is SD.

‡ - Apparent PD within 12 weeks of radiation

Response imaging criteria:

**From Lesions to Timepoint**

| | CR | PR | SD | PD |
|---|---|---|---|---|
| T1-Gd+ | None | ≥50% | <50%↓-<25%↑ | ≥25%↑* |
| T2/FLAIR | Stable or ↓ | Stable or ↓ | Stable or ↓ | ↑* |
| New Lesion | None | None | None | Present* |
| Corticosteroids | None | Stable or ↓ | Stable or ↓ | NA |
| Clinical Status | Stable or ↑ | Stable or ↑ | Stable or ↑ | ↓* |
| Requirement for Response | All | All | All | Any* |

**CR = Complete Response, PR = Partial Response, SD = Stable Disease, PD = Progressive Disease**

* Progression occurs when any of these criteria are met present.

**NA:** An increase in steroid dose alone will not cause a determination of progression in the absence of clinical deterioration or radiographically documented lesion growth.

#### Pseudoprogression

- Pseudoprogression
  - Enhancement that simulates tumor growth, most often caused by radiation (whole brain or focal).
  - Growth of existing lesions or appearance of new lesions within 12 weeks of completion of radiation therapy may be the result of treatment effects rather than growth of tumor.
  - Continued follow-up imaging can determine whether initial lesion growth was true progression or pseudoprogression.
    - If lesion continues to enlarge, the initial growth is called true progression
    - If lesion stabilizes or shrinks, the initial growth is confirmed as pseudoprogression
      - In such cases, the baseline SPD is no longer included when choosing the nadir value for the purposes of determining when progression occurs
  - Diffusion weighted imaging can help distinguish pseudoprogression from true tumor growth, but its use is still experimental. The use of MR perfusion and spectroscopy is also being explored.

**APPENDIX XII: Treatment plan, Delivery, and Target/OAR parameters**

| Treatment technique | Machine parameters | Delivery parameters | Target parameters | OAR parameters |
|---|---|---|---|---|
| Beam # | Table range shifter (Yes/No) | IGRT type | Tabular dose EQD2 data for target | Tabular dose EQD2 data for region-specific OARs |
| SFUD/IMPT | Nozzle range shifter (Yes/No) | Daily shifts | Tabular acute BED data for target | Tabular late BED data for region-specific OARs |
| Beam angles | Aperture (Yes/No) | Anesthesia / sedation use | Conformity metrics | Constraint achieved for each OAR |
| Couch positions | # Repaint | Gantry # | Target coverage metrics | Integral dose |
| # Replans / phase | Spot size | Treatment break (Yes/No) | Homogeneity indices | OAR doses across robustness scenarios |
| # Treatment phases | # Painted layers | Break length (days) | Target coverage across robustness scenarios | dLET map for OARs |
| | | Treatment time | dLET map for target | |

**APPENDIX XIII: Skin Assessment and Toxicity**

- Toxicity will be graded according to CTCAE.
- Assessment should note the status of any scar or incision within the radiation therapy beam path.
- Scar/incision status shall be categorized as follows:
    - Healed: Sutures/staples have been removed with tissue surfaces approximated with no signs of infection and only minimal erythema.
    - Healed by primary intention: Incisional surfaces are well approximated, but sutures or staples are still in place.
    - Healed by secondary intention: Incisional surfaces are not yet approximated and there are signs of residual tissue loss.
    - Healed by tertiary intention: Closure or suture is delayed because of poor circulation or drainage.

**APPENDIX XIV: Systemic Disease Status Assessment**

- Systemic disease status may be classified as follows:
  - Treated systemic disease: prior chemotherapy for current extent of disease.
  - Untreated systemic disease: no prior chemotherapy for current extent of disease.

**APPENDIX XV: LOCAL Disease Status Assessment**

- Local disease status may be classified as follows:
  - Treated local disease:
    - Prior surgery
    - Prior chemotherapy
    - Prior chemotherapy and surgery
    - Prior other local therapy
    - Prior other local therapy and chemotherapy
  - Untreated local disease: no prior therapy for current extent of localized disease

- Additionally, local disease may be classified as follows according to amount of the local disease:
  - No gross or microscopic residual disease
  - Residual microscopic residual disease
  - Gross residual disease representing < 10% of the original tumor volume
  - Gross residual disease representing 10% to 60% of the original tumor volume
  - Gross residual disease representing > 60% of the original tumor volume
